# Population level impact of increasing tuberculosis treatment coverage and addressing determinants of risk in men: a modelling study in Kenya, Malawi, Nigeria, and Uganda

**DOI:** 10.1101/2025.03.31.25324963

**Authors:** Alexandra S Richards, Mphatso D Phiri, Jasper Nidoi, Jeremiah Chakaya, Peter MacPherson, Bruce J Kirenga, John S Bimba, Chukwuebuka Ugwu, Rhoda Pola, S Bertel Squire, Katherine C Horton

## Abstract

**Background:** Globally, the burden of tuberculosis (TB) falls more on men than women and children, and there are large gaps between men and women at all stages of exposure, disease incidence, and treatment. We examined the impact of addressing determinants of these gender gaps in Kenya, Malawi, Nigeria, and Uganda.

**Methods and Findings:** We created a deterministic transmission model of TB, with strata for sex, age, and HIV co-infection and antiretroviral therapy, and calibrated to country-specific data on prevalence, incidence, mortality, and notifications between 2010 and 2022. We then examined the potential epidemiological impact of strategies to increase treatment coverage among men, and decrease the effects of social and structural determinants that increase men’s risk of developing TB. We investigated impact (overall and by age and sex) on projected incidence and mortality in 2035, and notification rates between 2025 and 2030.

Relative reductions in overall TB incidence from increased treatment coverage among men ranged from 4.8% [95% uncertainty interval (UI)0.7-10.6%] in Malawi to 24.8% [UI 14.3-37.0%] in Uganda. Reducing men’s excess risk of TB led to a similar range in relative reductions from 8.4% [UI 6.6-10.4%] in Nigeria to 23.3% [UI 18.0-30.0%] in Uganda. Combining strategies increased the relative reduction beyond that seen with one strategy alone, but as would be anticipated by greater prevention of incident TB, less than the sum of both strategies. Impacts extended across the population with median estimates of country-level declines in incidence of between 1.6—10.6% and 2.8—19.5% in women and children respectively, across the four countries. Increasing treatment coverage increased the median country-level estimates of notifications between 3.1% and 12.4% in the first three years, however combining increased treatment coverage with a reduction in risk reduced the median annual country-level notifications by 6.7% to 18.1% by 2035.

**Conclusion:** Strategies that prioritise increasing TB treatment coverage among men and mitigating men’s higher susceptibility to TB could reduce disease burden for men, women, and children. Such gender-responsive strategies are essential to ensure a person-centred TB response and accelerate global progress towards the End TB targets.

## Introduction

Tuberculosis (TB) is the leading infectious cause of death globally, with 10.8 million people developing TB disease and 1.25 million people dying from TB in 2023.[1] The greatest burden of TB falls on men (aged ≥15 years), who account for an estimated 55% of all cases globally, compared to 33% in women and 12% in children.[1] These sex differences exist through the course of the disease, with men having a greater exposure to *Mycobacterium tuberculosis (M*.*tb)*, a higher risk of infection, and a higher incidence of disease than women.[2,3] Men also experience longer durations of untreated disease due to social and structural barriers to healthcare, leading to higher prevalence to notification (P:N) ratios for men than women.[1,4,5] As such, men are estimated to make up two-thirds of the adults who developed TB but were unable to access diagnosis and treatment in 2023.[1]

People unable to access diagnosis and treatment represent a key failure in the TB response, as they remain outside the reach of lifesaving care and contribute to ongoing community transmission. Concerted efforts to reach these people are essential to end TB. Between 2015 and 2023, there were reductions in global incidence of 8.3% and mortality of 23%, but these gains fall well below the EndTB 2025 milestones of 50% and 75% reductions respectively.[1] The EndTB targets for 2035 go further, with an aim of a 90% reduction in incidence and 95% reduction in mortality, relative to levels in 2015; current trends are well below what is needed to achieve these targets.[1]

Tremendous progress in reducing the TB burden has been made in the World Health Organization (WHO) African region, with a 24% reduction in incidence since 2015.[1] However, this progress is driven primarily by a reduction in HIV-associated TB burden through the rapid expansion of HIV prevention and treatment during this time.[1] Whilst TB incidence in HIV-positive people has almost halved in in the last eight years, the same is not true among HIV-negative people, where TB incidence has been increasing steadily since 2015 and is now 10% higher than it was in 2015.[1] As such, half of the WHO’s high TB burden countries are in the African region.[1]

Our work considers four countries with a high burden of TB and HIV from the African region: Kenya, Malawi, Nigeria, and Uganda.[1,6] Spread across east, south, and west Africa, each country presents a unique profile of the challenges faced to reduce the TB burden. The WHO considers Kenya, Nigeria, and Uganda to have a high TB burden; all four countries to have a high TB-HIV burden; and Nigeria to have a high drug-resistant TB burden.[1] Between 2012 and 2016, these countries each conducted a national TB prevalence survey that estimated the true burden of undiagnosed TB within the country.[1,7] These surveys also highlighted the gender disparities, with higher estimates for TB prevalence for men than women in each country.[1,7]

As in the rest of the world, these disparities in burden can partially be explained by gaps in access to treatment and diagnosis, but may also be the result of other social and structural determinants of health that impact men more than women. At an individual level, there are likely biological differences that make men more susceptible to TB than women.[8] Interacting with these baseline differences are the social aspects such as higher levels of alcohol consumption and tobacco smoking among men than women, and institutional aspects such as occupational risks, limited social protection, higher rates of incarceration, and primary healthcare systems oriented towards maternal and child health.[1,9–11] Men also mix socially more with other men than with women, meaning that alongside being more at risk of TB, they also have greature exposure to TB.[12–15]

Reducing the burden of disease in men by addressing these disparities could accelerate progress towards the EndTB targets. To investigate the potential epidemiological impact of strategies to improve men’s access to TB care and to reduce men’s excess risk of TB, we have used mathematical modelling to examine the impact of each of these strategies on incidence and mortality at both a population level and among sub-populations, specifically men, women, and children.

## Methods

### Model

We developed a compartmental model of TB disease and *M*.*tb* transmission, building on previously published models (**Section 1 in S1 Appendix**).[16,17] As shown in Figure 1, the model comprises four TB states: susceptible (uninfected), infection, asymptomatic TB, and symptomatic TB.[18] The states of infection and disease are stratified by TB treatment history, and all compartments are further stratified by sex (male/female), age group (0-14, 15-29, …, 60-74, 75+), and HIV and antiretroviral treatment (ART) status (HIV negative, HIV positive no treatment, HIV positive 0-6 months treatment, HIV positive 7-12 months treatment, HIV positive >12 months treatment).

**Figure 1:**
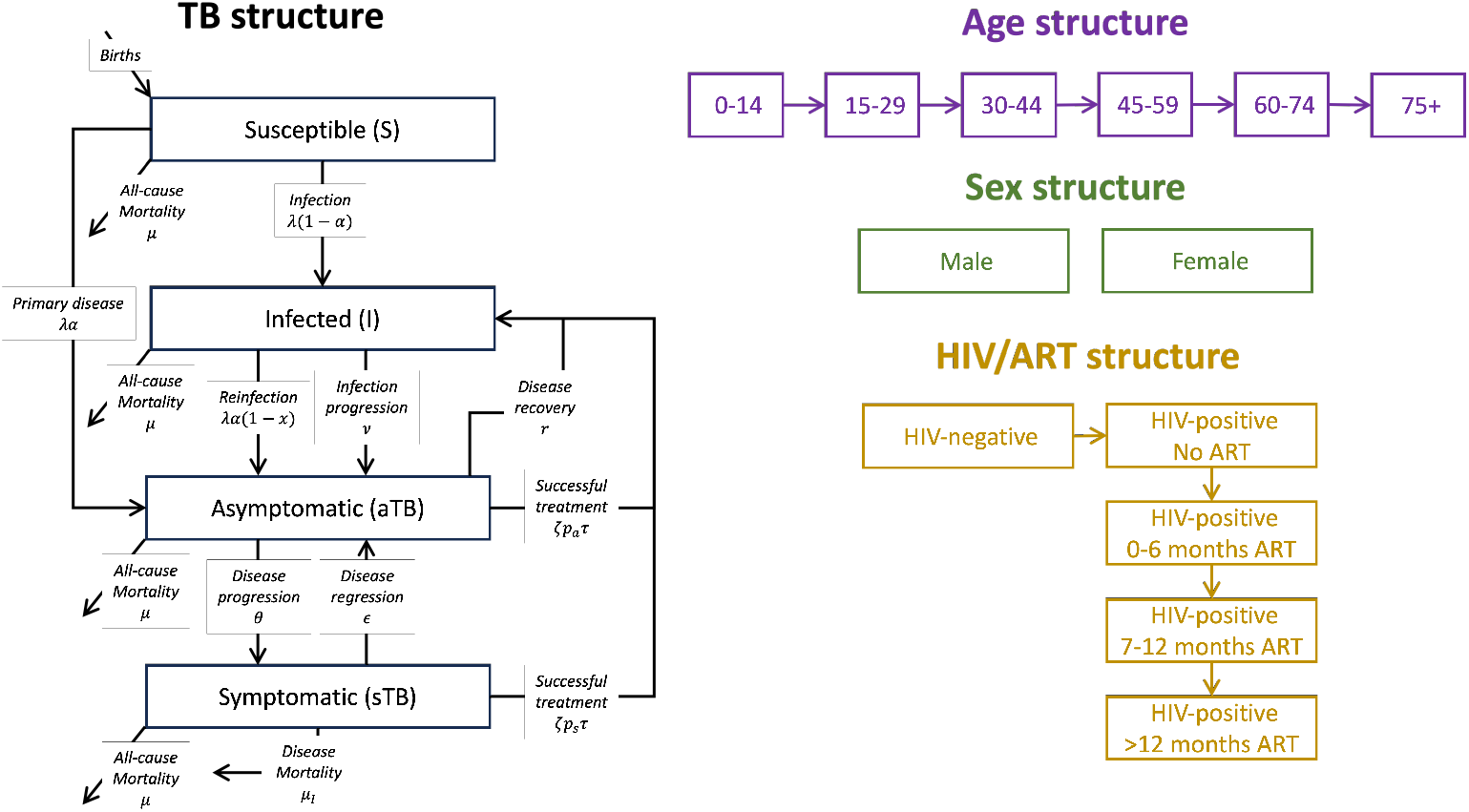
Model structure showing the TB transitions (left), the demographic transitions (top right), the gender strata (middle right) and the HIV/ART transitions (bottom right)

### Data

For each country, the population in each age group, HIV incidence and prevalence, and ART uptake incidence and prevalence were informed by time-series estimates, reported annually by sex in five-year age groups, from the DemProj and AIM models of the Spectrum suite.[19–21] At each time point, values from our model were cross-checked with AIM outputs and the incidence of HIV and ART initiation required to match the estimate was calculated and applied.[16] The model was also informed by annual country-level data on vaccination coverage (with the Bacillus Calmette-Guérin [BCG] vaccine), TB treatment coverage (defined as the proportion of estimated incidence that is notified each year), and TB treatment success (the proportion of people who successfully complete a course of TB treatment) for both HIV-negative and HIV-positive populations, as reported by the WHO.[22–25]

Sex- and age-assortative mixing patterns are reflected in the model with a social contact matrix between men (males ≥ 15 years), women (females ≥15 years) and children (both sexes age < 15 years). Country-specific sex- and age-assortative matrices were only available for Kenya and Malawi; we used a summary contact matrix from all available surveys conducted within Africa for Nigeria and Uganda.[12–15]

Prior ranges for all parameters were informed by literature, where available. TB natural history parameters were informed by studies that have collated historical data to best represent the progression of *Mtb* infection and TB disease.[26–29] The impact of age, HIV and ART on TB natural history parameters were based on previous estimates.[16,17] We assumed there were differential effects on treatment coverage from the population average depending on age (children less likely to access care for TB), sex (men less likely, and women more likely to access care for TB), symptom status (asymptomatic disease less likely to prompt care-seeking), and ART status (more likely to receive TB testing as part of ART care). To account for the increased frequency of TB disease among men compared to women, we incorporated a parameter to represent the quantifiable, but not specifically assigned, excess risk. A full list of parameters and prior ranges is available in the supplementary materials (**Section 3 in S1 Appendix**).

### Calibration

For each country, we calibrated the model to country-specific epidemiological targets of TB prevalence, incidence, mortality, and case notificationsfor the overall population and, where possible, for age, sex, and HIV strata.[6,22,30–32] TB prevalence targets, both overall and by sex, were informed by the national TB prevalence surveys.[30,33– 36] Additional targets included: estimated TB incidence, overall and for men, women and TB-HIV status; TB mortality, overall and by TB-HIV status; notification rates, overall and by sex, age, and HIV status. Each of these data points were sourced from the WHO TB data files on incidence, burden, and notifications respectively.[22,31,32]

The model was calibrated using a history matching and emulation algorithm (*hmer*) in R.[37–39] This algorithm is an iterative process of testing the outputs of a selection of parameter sets that span the available space against the calibration targets, calculating the implausibility of those parameter sets, and discarding the most implausible.[37] This results in a set of parameters that fit the calibration targets and span the available parameter space.[37] For each country, we ran the algorithm through up to 10 rounds of the iterative process (stopped earlier if there was no improvement between already successful rounds), and then used the emulators from the best performing round to generate at least 1000 parameter sets for each country.[37] The resulting parameter sets were randomly sampled to select 1,000 parameter sets to be used for further analysis. Figures showing the progress of each round of calibration are included in the supplementary materials for each country (**Figures 6, 9, 12, and 15 in S1 Appendix**).

Results for the calibration are presented with plots of the distributions of the 1,000 parameter sets compared to the priors and median values of those parameters with 95% credible intervals (CrIs), along with plots illustrating how the calibrated model fitted the epidemiological targets over time (**Figures 7, 8, 10, 11, 13, 14, 16, and 17 and Tables 23-26 in S1 Appendix**). Results are presented with the median and 95% uncertainty intervals (UIs) of the outputs from these 1,000 parameter sets.

### Gender-responsive strategies

We considered three main strategies: improving TB treatment coverage in men to match the levels achieved in women by 2030, reducing men’s excess risk of TB, and a combination of the two. Each strategy was implemented over a period of five years from 2025 to 2030 with an S-shaped uptake curve (further details can be found in the supplementary materials - **Section 6 in S1 Appendix**). Improving treatment coverage in men was modelled as increasing the calibrated parameter for treatment coverage specific to men to match the calibrated parameter for women’s treatment coverage.

The reduction of excess risk was modelled as reducing the calibrated parameter representing men’s risk by 50% of the excess above 1.[8] The third strategy coombined these two strategies over the same five-year period. Supplementary analyses of excess risk reductions by 25% and 75% have been included in the supplementary materials (**Section 7.3 in S1 Appendix**).

### Analysis

We evaluated each strategy by comparing the projected TB incidence, prevalence, mortality, and notifications (**Section 7.2 in S1 Appendix**) in 2035 and 2050 to a business-as-usual (BAU) scenario. The BAU scenario assumed that the TB treatment coverage in each country would continue to improve, and other time dependent parameters such as treatment success and BCG coverage would continue at similar levels to 2023, but that no new strategies for TB or HIV would be introduced. Unless otherwise specified, results are presented for the entire population.

All model development, calibration, and analyses were performed using R 4.3.3.[39] The code used can be found in the GitHub repository https://github.com/alexandrasrichards/LIGHT.

## Results

### Calibration

Each country was successfully calibrated to its respective set of epidemiological target ranges, with posterior parameter estimates and model estimates compared to calibration targets available in the supplementary materials (**Section 5.2.1 in S1 Appendix**). All four countries showed decreasing prevalence, incidence, and mortality over the period 2010-2022. In both Kenya and Malawi, the P:N ratio decreased slightly over time for both men and women. Nigeria and Uganda had relatively stable P:N ratios until each country began increasing active case finding initiatives in 2020 and 2022 respectively, when the P:N ratio decreased for men and women.[40,41] Despite these improvements in treatment coverage, in all four countries there was a small, but continued, increase in the male-to-female prevalence ratio from 2010 until 2022. Plots showing the calibrated model trends are shown in the supplementary materials (**Figures 8, 11, 14, 17 in S1 Appendix**).

### Impact on burden

Increasing men’s treatment coverage was projected to have the greatest impact on disease burden in Nigeria and Uganda with a 16.6% [UI 8.6—25.3%] and 24.8% [UI 14.3—37.0%] reduction, respectively, in 2035 incidence relative to BAU. This strategy was expected to avert 34,634 [UI 18,714—55,087] TB deaths in Nigeria and 6,532 [UI 3,595—11,447] TB deaths in Uganda between 2025 and 2035. The relative reduction in TB incidence was lower for this scenario in Kenya, with an 8.0% [UI 1.9—15.5%] reduction in 2035 incidence relative to BAU, and Malawi, where 2035 incidence was 4.8% [UI 0.7—10.6%] lower than BAU (Figure 2). In Kenya, increasing men’s treatment coverage was estimated to prevent 6,990 [UI 1,385—14,857] TB deaths, while in Malawi, 1,065 [UI 151—2,395] TB deaths were averted between 2025 and 2035.

**Figure 2:**
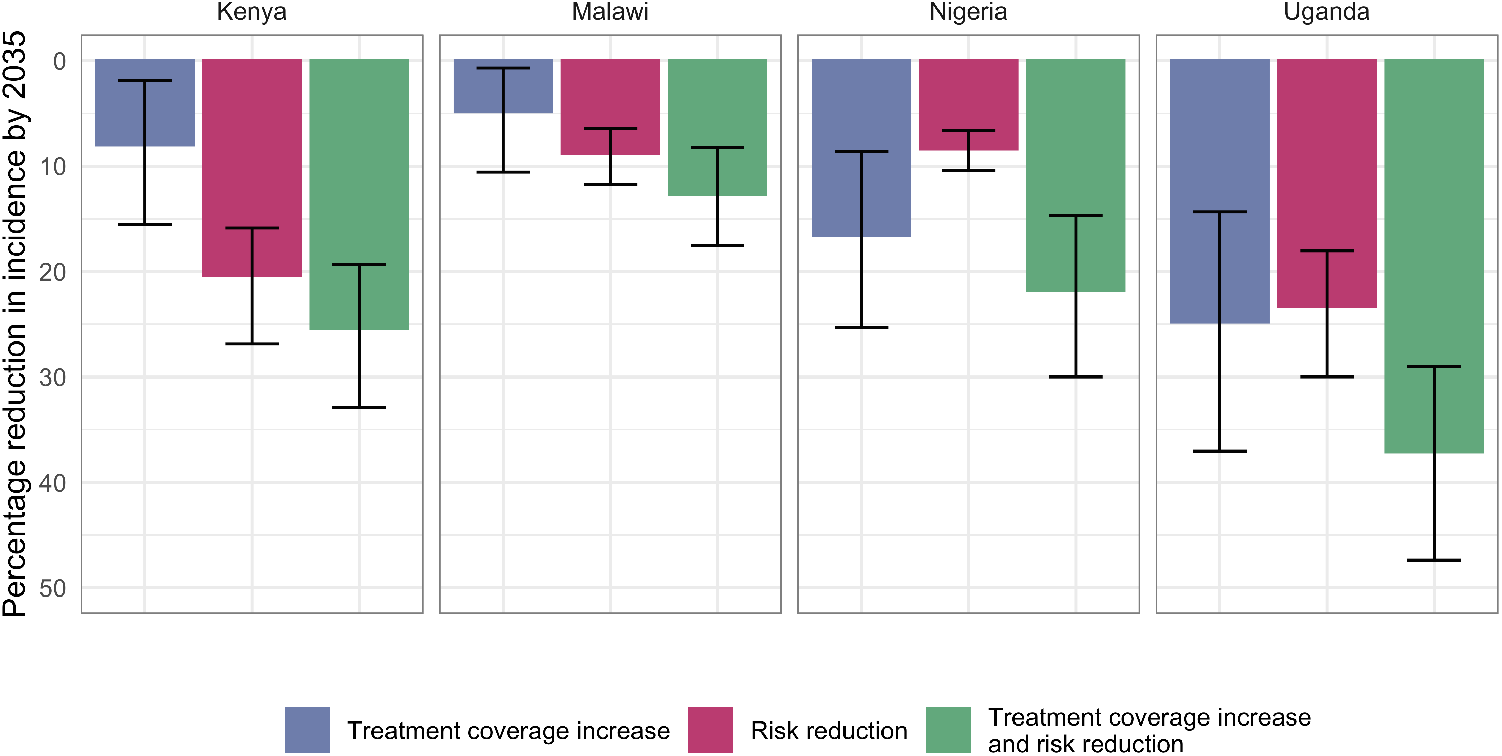
Percentage reductions in 2035 TB incidence, relative to a business-as-usual scenario, across the three strategies to address gender disparities in TB – increasing treatment coverage in men, reducing men’s excess risk of TB by 50%, and the combination of these two. Colloured bars show the median estimate and error bars show the 95% uncertainty interval.

Reducing men’s excess risk of TB lead to similar relative reductions in 2035 incidence relative to BAU, at 20.4% [UI 15.9—26.9%] and 23.3% [UI 18.0—30.0%] in Kenya and Uganda respectively, with 13,208 [UI 8,993—20,070] and 3,986 [UI 2,427—6,224] TB deaths averted respectively. The impact of this strategy was projected to be more limited in Nigeria and Malawi, with 8.4% [UI 6.6—10.4%] and 8.8% [UI 6.4—11.7%] respective reductions in 2035 incidence relative to BAU. Between 2025 and 2035, reducing men’s excess TB risk was projected to avert 1,234 [UI 751—1,992] TB deaths in Malawi and 11,222 [UI 8,104—15,914] TB deaths in Nigeria.

Combining the two strategies was projected to lead to greater reductions in TB burden. As with the individual strategies, the greatest relative reduction from the combined strategy was in Uganda where we projected a 37.1% [UI 29.0—47.4%] reduction in 2035 incidence relative to BAU and 8,683 [UI 5,391—13,799] TB deaths averted between 2025 and 2035. Reductions in 2035 incidence were similar in Nigeria and Kenya, 21.8% [UI 14.7—30.0%] and 25.4% [UI 19.3—32.9%] respectively. Due t differences in population size, the number of TB deaths averted from this strategy was much larger in Nigeria (41,539 [UI 25,937—63,160]) than in Kenya (18,561 [UI 11,784—28,873]). With a 12.7% [UI 8.2—17.5%] reduction in 2035 incidence relative to BAU, impact was more limited in Malawi, but we project 2,157 [UI 1,159—3,706] TB deaths would be averted here between 2025 and 2035.

### Impact on men, women, and children

The impact of these strategies varied among men, women, and children (Figure 3). As the strategies targeted men, as exppected, the greatest impact was seen among men. The median estimate for the percentage reduction in incidence for men ranged between 20.1% (Malawi) and 47.4% (Uganda) in 2035 compared to BAU, translating to a percentage increase from the population level of between 28% (Malawi) and 58% (Uganda). In women, the combined strategy reduced incidence by between 3.7% (Malawi) and 10.6% (Uganda), equivalent to a percentage decrease from the population level of between 59% (Kenya) and 74% (Nigeria). Children were estimated to have a median incidence reduction of between 6.7% (Malawi) and 19.5% (Uganda), equating to between 36% (Nigeria) and 57% (Kenya). Despite the lower impact in women and children, all strategies provide a significant reduction in incidence

**Figure 3:**
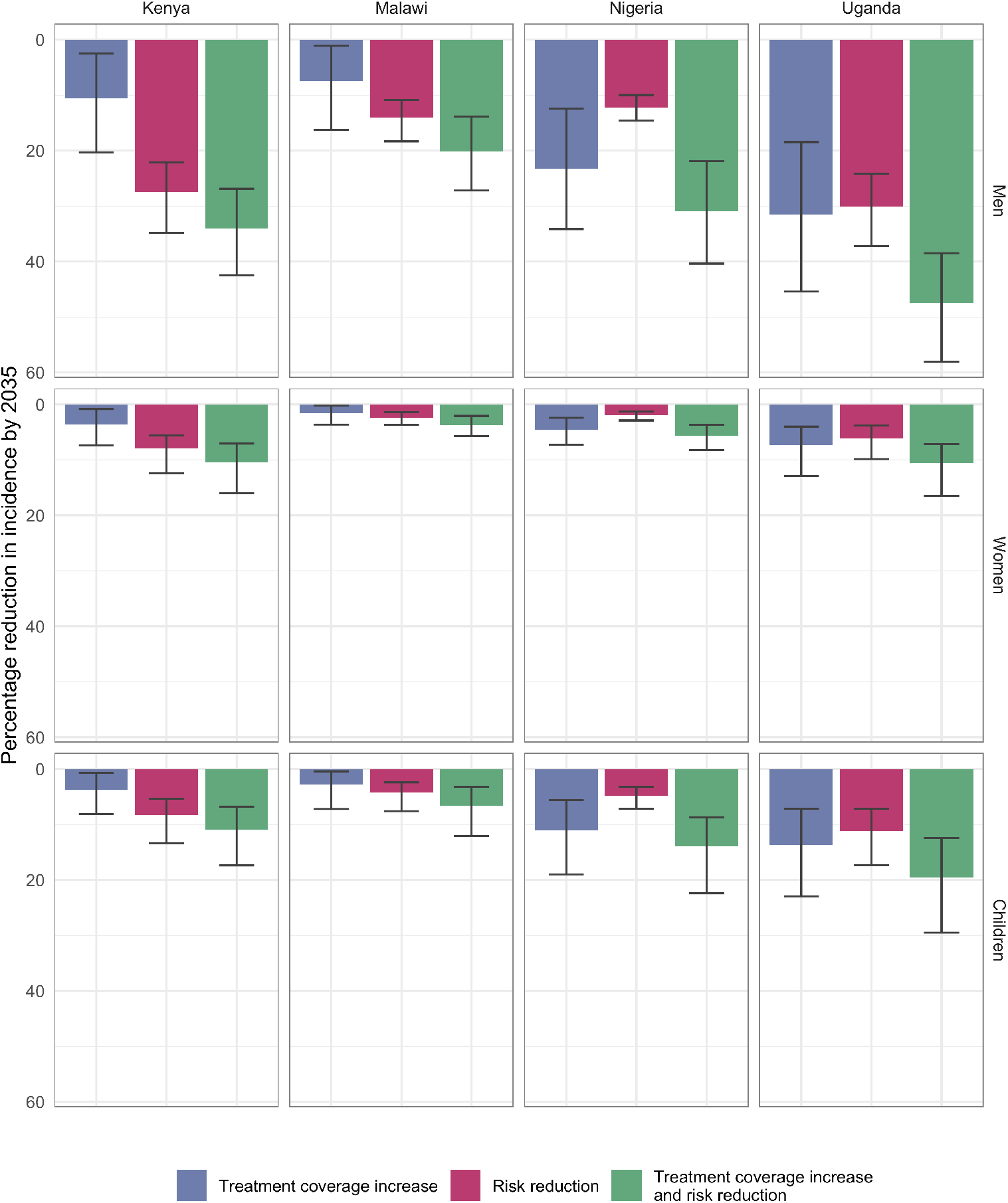
Percentage reductions in 2035 TB incidence in men, women, and children, relative to a business-as-usual scenario, across the three strategies to address gender disparities in TB - increasing treatment coverage in men, reducing men’s excess risk of TB by 50%, and the combination of these two. Coloured bars show the median estimate and error bars show the 95% uncertainty interval.

### Impact on notifications

As expected, increasing treatment coverage in men initially increased notifications across all countries with peak increases of 4.1% [UI 0.8-8.4%] and 3.1% [UI 0.5-6.8%] in Kenya and Malawi respectively in 2029. These percentage increases translated to an additional 2,264 [UI 442-4,927] notifications in Kenya and an additional 419 [UI 58-923] notifications in Malawi in 2029 compared to BAU. The peak increase in notifications in Nigeria and Uganda occurred a year earlier, but resulted in much greater increases at 10.5% [UI 5.2-17.1%] and 12.4% [UI 6.9-20.7%] respectively, equating to an additional 16,699 [UI 8,545-27,802] notifications in Nigeria and an additional 4,158 [UI 2,278-7,249] notifications in Uganda in 2028 compared to BAU.

Conversely, reducing men’s risk of TB did not increase notifications at any point, with reductions observed from 2027 onwards. Combining these two interventions resulted in marginal increases in notifications in Kenya and Malawi (0.8% [UI −2.1-5.1%] and 1.3% [UI −1.1-4.8%] respectively) a year earlier than with increasing treatment coverage alone. Nigeria and Uganda still expperienced a substantial increase in notifications (8.8% [UI 3.6-15.0%] and 8.3% [UI 2.9-15.9%] respectively), with an additional 13,987 [UI 5,870-24,608] notifications in Nigeria and an extra 2,744 [UI 980-5,688] notifications in Uganda, in 2028.

While short-term impacts over the first two years of implementation showed increased notifications, longer term impacts showed reduction in notifications across strategies by 2035. Figure 4 shows that reductions in overall notifications by increasing treatment coverage among men had substantial impact in Uganda (7.0% [UI 3.4-12.5%] reduction) but resulted in marginal changes in Kenya, Malawi, and Nigeria. Changes in notifications from booth the risk reduction strategy and the combined strategy were similar, with greater reductions in Kenya and Uganda (17.6% [UI 13.7-23.2%] and 18.1% [UI 14.0-23.8%] respectively) than in Malawi and Nigeria (6.9% [UI 5.1-9.1%] and 6.7% [UI 5.3-8.2%] respectively).

**Figure 4:**
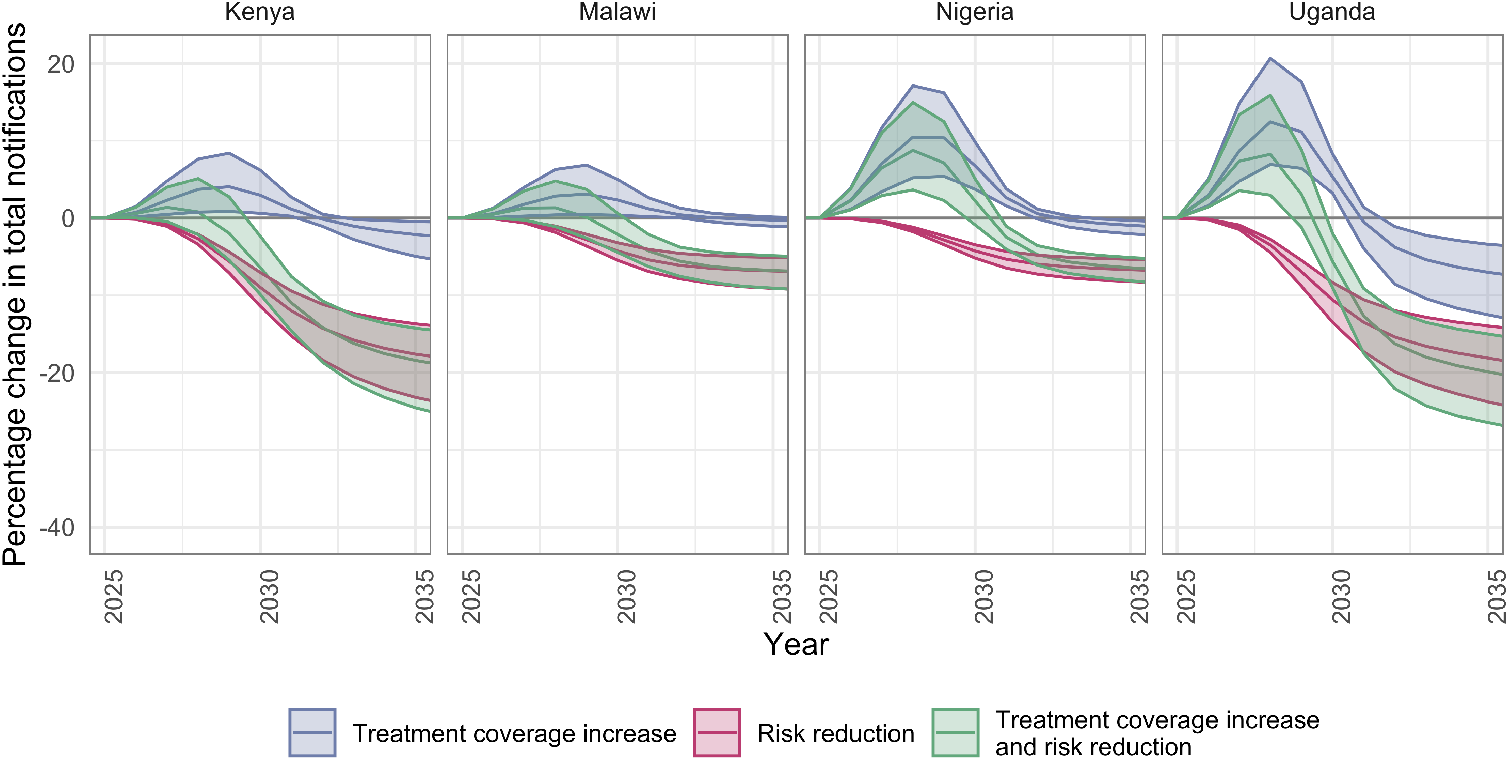
The percentage change in notifications from a baseline of no intervention for the five years before, the five years during, and the five years after the implementation of each strategy (increasing treatment coverage in men, reducing men’s excess risk of TB by 50%, and the combination of these two). The central lines show the median estimate for the relative percentage reduction and the shaded regions show the 95% uncertainty interval.

### Sensitivity analyses

Sensitivity analyses were conducted for a 25% and 75% reduction in excess risk. As expected, reducing excess risk by only 25% reduced the impact of risk reduction and the combined strategy, while reducing excess risk by 75% increased effects (**Figures 23 and 24 in S1 Appendix**). Notable differences were that with a 25% reduction in excess risk, the risk reduction strategy alone became substantially less effective than the treatment coverage strategy in Nigeria, and with a 75% reduction in excess risk, the risk reduction strategy alone became substantially more effective than the treatment coverage strategy in Kenya.

Percentage reductions in each measure were similar across all four countries.

While main results focus on the period through 2035, we also modelled predictions of impact through 2050 (**Figures 25 and 26 in S1 Appendix**) and, with time, each intervention had a greater impact on incidence, likely reflecting non-linear effects. Reduction in excess risk in men yielded slightly higher improvements over time than improving treatment coverage in men and there was more of an improvement of impact for women and children than there was for men. For each intervention, the number of notifications also continued to reduce over time, reflecting progressive reductions in diseasae burden, with the greatest improvement in the combined strategy in all four countries.

## Discussion

Our results show that addressing gender disparities in access to TB care and determinants of risk may result in declines in TB incidence of between 12.7% [UI 8.2— 17.5%] in Malawi and 37.1% [UI 29.0—47.4%] in Uganda. These reductions are greater than the reductions that have been achieved globally in the last 10 years and so have important implications for TB care, prevention, and programming.[1] In settings with large gaps in treatment coverage between men and women, such as Nigeria and Uganda, closing this gap may be particularly effective at reducing TB burden in men and subsequently throughout whole population. Where gendered gaps persist beyond treatment coverage, such as in Kenya, strategies that focus on reducing men’s risk of TB may be more effective at reducing TB burden. Whilst these strategies focus solely on men, the impact extends across the population, with reductions in incidence and mortality in women and children.

Whilst the estimated reductions in incidence could seem large for only 10 years, there have been trials conducted recently, on varying scales, that show similar percentage reductions, highlighting the feasibility of our estimates. Community-wide screening in Viet Nam resulted in a 56% reduction in prevalence over 4 years.[42] Whilst this is a larger impact than seen in any of our modelled results, the strategy involved annual rapid nucleic acid amplification testing for all adults, making this a costly and logistically challenging strategy.[42] Conditional cash transfers in Brazil have also seen a reduction in TB incidence of 41% which is a similar impact as the expected population-level impact of the combined strategy in Uganda.[43] As the conditions of these cash transfers include healthcare and education, and other studies of these transfers have shown they have reduces the burden of many chronic diseases, it is likely that it has both increased access to healthcare and reduced some of the excess risks in men, providing a real world example and confirmation of the effects modelled.[43] The RATIONS trial in India, where TB patients and their household contacts received nutritional support also saw a similar reduction in TB incidence of between 39 and 48%.[44]

Closing gaps in treatment coverage requires an understanding of the cultural, gender, and healthcare norms in each country, and so are likely to be different even across the four countries included here and may change with time. A common theme across study countries is that there are societal expectations for men and women that may impact uptake and effectiveness of strategies, and understanding these will play a key role in informing strategies intended to increase access to care for men.[5,45] However, there has also been research conducted in each of the four countries to understand strategies that might help. In Uganda, recent work as part of the IGNITE study found that stigma and societal expectations of masculinity, paired with clinic hours being only available during work hours prevented men from accessing care.[46] The proposed interventions included strategic clinic opening times, either outside of normal working hours, or on market days, and education to improve TB awareness among men.[46] In Nigeria in the DESTINE study a Delphi process found that targeted awareness campaigns and screening in men’s congregate settings was the preferred strategy to increase men’s access to care.[47] Both the proposals of clinics on market days and outside of working hours were found to be less preferred in this Nigerian setting.[47] Occupational screening has yielded promising results in Kenya, with twice as many men as women initiated on treatment, however community involvement and adaptability of processes are required for success across workplaces.[48] Targeting male dominated industries, such as the fishing industry, and prisons, where 95% of incarcerated individuals are male, was trialled in Uganda, and resulted in increased notifications across all groups.[49–51] Other studies have looked at opening male-specific clinics in rural Kenya, and suggested that improved labour regulations and payment during sick leave in Malawi could help increase men’s participation in healthcare.[52,53]

The distribution of TB determinants varies across different countries, and each country’s capacity to implement strategies that reduce these risks will differ.[54] In order to achieve the results presented here, determinants that impact men more than women will need to be addressed. Many social and structural determinants which can increase the risk of infection and progression to disease are more prevalent in men than women, such as tobacco and alcohol use.[1,9] Men are also more likely to work in occupations with low ventilation or exposure to lung irritants like silica dust and more likely to experience incarceration.[10,11] It is likely that biological sex characteristics also contribute, therefore a complete reduction of excess risk was not considered in this work.[8] Some of these determinants will be harder to eliminate, and the fact that addressing their root causes llies outside the scope of a national TB programme.[55] Any strategy to effectively reduce men’s excess risk will require a multi-sectoral approach.

Our analysis omitted some impacts of these interventions that can’t fully be modelled. Firstly, there is potential that any strategy that increases men’s access to care, or decreases their excess risk may also impact women and children. This could come in the form of increased awareness and reduced stigma improvind utilisation of TB services or through, for example, decreased exposure to secondhand tobacco smoke that would reduce risk. While we cannot quantify this additional effect, it would likely increase reductions in burden estimated here, both overall and among women and children. Secondly increasing men’s access to TB care may offer opportunities for earlier diagnosis and treatment of other conditions, such as hypertension and diabetes, and addressing social and structural determinants may also reduce risk for other infectious and chronic conditions.[56,57] These additional impacts are beyond the scope of this work, but may be useful to consider.

We did not explicitly model the impact of the COVID-19 pandemic and rellated non-pharmaceutical interventions that may have affected risks of transmission and reduced treatment coverage; rather we have calibrated our model to estimates before and after the pandemic.[58] In our BAU scenario, we recognise other ongoing work to improve treatment coverage and have assumed that treatment coverage will ccontinue to increase for all four countries included in this work.[59] We do not expect the proportional reductions shown in our modelling to change without gender-responsive approaches in each country. Our results do not incorporate within-country heterogeneity in disease burden, so any strategies introduced to higher burden areas, or areas with greater sex disparities, using local knowledge could result in greater impacts on burden than have been presented here.[60] Country-specific social contact matrices were only available for Kenya and Malawi, so the combined contact matrix used for Nigeria and Uganda may not reflect the true distributions of contacts by age and sex. Despite these limitations, the model calibration reflects trajectories of disease that have been observed over a 15-year period, and therefore the results llikely provide a reasonable estimate of the impact of the strategies that have been implemented.

Our modelling work highlights the population-level benefits of reaching men, a high burden group under-represented in TB prevention and care in Kenya, Malawi, Nigeria, and Uganda. These findings likely have implications for other settings given the widespread nature of gender disparities in TB burden and care. Our findings suggest that implementation of strategies in line with those described here could reduce TB morbidity and mortality across the population, accelerating proogress towards the EndTB milestones.

## Data Availability

All data produced in the present work are contained in the manuscript

## Acknowledgements

Authors appreciate support from members of the national tuberculosis programmes in each of the countries in this analysis, including Wendy Nkirote, Aiban Ronoh, and Immaculate Kathure (National Tuberculosis, Leprosy, and Lung Disease Program, Kenya), Obioma Chijioke-Akaniro (National Tuberculosis, Leprosy and Buruli Ulcer Control Programme, Nigeria), Kuzani Mbendera, and Tisungane Mwenyenkulu (National Tuberculosis Control Programe, Malawi), and Dr Stavia Turyahabwe (Ministry of Health, Uganda), Dr Geofrey Amanya and Mr Muzamiru Bamuloba (National Tuberculosis and Leprosy Program, Uganda) who shared their insights and understanding of TB epidemiology with the research team. Authors also appreciate support from Andy Iskauskas (University of Durham) and Nicky McCreesh (London School of Hygiene and Tropical Medicine) for their assistance in the calibration methodology. Authors also acknowledge support from management, research, research uptake, and programme management teams within The LIGHT Consortium

## Author contributions

**Conceptualization:** ASR, KCH, PM, SBS

**Formal Analysis:** ASR

**Funding Acquisition:** KCH, JC, PM, JSB, BJK, SBS

**Investigation:** ASR

**Methodology:** ASR, KCH

**Resources:** KCH, SBS

**Software:** ASR, KCH

**Supervision:** KCH, SBS

**Validation:** MDP, JN, CU, RP, JC, PM, JSB, BJK, KCH

**Visualization:** ASR

**Writing – Original Draft:** ASR

**Writing – Review & Editing:** ASR, KCH, MDP, JN, CU, RP, JC, PM, JSB, BJK, SBS

## Notes

### Competing Interest Statement

The authors have declared no competing interest.

### Funding Statement

ASR, MDP, JN, JC, PM, BJK, JSB, CU, RP, SBS, and KCH are supported by the UK Foreign, Commonwealth and Development Office (Leaving No-one Behind Transforming Gendered Pathways to Health for Tuberculosis). KCH is also supported by the US National Institutes of Health (R-202309-71190). SBS is also supported by the Start4All programme, funded by Unitaid. The views expressed do not necessarily reflect the UK Government's official policies. The funders had no role in study design, data collection and analysis, decision to publish, or preparation of the manuscript.

## References

1. World Health Organization. Global tuberculosis report 2024. World Health Organization; 2024. Available: https://www.who.int/teams/global-tuberculosis-programme/tb-reports/global-tuberculosis-report-2024

2. Horton KC, Sumner T, Houben RMGJ, Corbett EL, White RG. A Bayesian approach to understanding gender differences in tuberculosis disease burden. American Journal of Epidemiology. 2018;187: 2431–2438. doi:10.1093/aje/kwy131

3. Rickman HM, Phiri MD, Feasey HR, Krutikov M, Horton KC, Dowdy D, et al. Sex Differences in the Risk of Mycobacterium Tuberculosis Infection: A Systematic Review and Meta-Analysis of Population-Based Immunoreactivity Surveys. SSRN [Preprint]; 2025 Jan. doi:10.2139/ssrn.5080783

4. Horton KC, MacPherson P, Houben RMGJ, White RG, Corbett EL. Sex differences in tuberculosis burden and notifications in low- and middle-income countries: A systematic review and meta-analysis. PLOS Medicine. 2016;13: e1002119. doi:10.1371/journal.pmed.1002119

5. Chikovore J, Hart G, Kumwenda M, Chipungu GA, Corbett L. ‘For a mere cough, men must just chew conjex, gain strength, and continue working’: The provider construction and tuberculosis care-seeking implications in blantyre, malawi. Global Health Action. 2015;8: 26292. doi:10.3402/gha.v8.26292

6. WHO Global Tuberculosis programme. WHO TB profiles. https://worldhealthorg.shinyapps.io/tb_profiles/; 2024.

7. Organization WH. National tuberculosis prevalence surveys 2007-2016. World Health Organization; 2021. Available: https://iris.who.int/handle/10665/341072

8. Gupta M, Srikrishna G, Klein S, Bishai WR. Genetic and hormonal mechanisms underlying sex-specific immune responses in tuberculosis. Trends in immunology. 2022;43: 640–656. doi:10.1016/j.it.2022.06.004

9. Wessels J, Walsh CM, Nel M. Smoking habits and alcohol use of patients with tuberculosis at standerton tuberculosis specialised hospital, mpumalanga, south africa. Health SA Gesondheid. 2019;24: 6. doi:10.4102/hsag.v24i0.1146

10. Davies LRL, Smith MT, Cizmeci D, Fischinger S, Lee JS-L, Lu LL, et al. IFN-γ independent markers of mycobacterium tuberculosis exposure among male south african gold miners. eBioMedicine. 2023;93. doi:10.1016/j.ebiom.2023.104678

11. Sequera VG, Aguirre S, Estigarribia G, Cellamare M, Croda J, Andrews JR, et al. Increased incarceration rates drive growing tuberculosis burden in prisons and jeopardize overall tuberculosis control in paraguay. Scientific Reports. 2020;10: 21247. doi:10.1038/s41598-020-77504-1

12. Kleynhans J, Tempia S, McMorrow ML, Gottberg A von, Martinson NA, Kahn K, et al. A cross-sectional study measuring contact patterns using diaries in an urban and a rural community in South Africa, 2018. BMC public health. 2021;21: 1055. doi:10.1186/s12889-021-11136-6

13. Thindwa D, Jambo KC, Ojal J, MacPherson P, Dennis Phiri M, Pinsent A, et al. Social mixing patterns relevant to infectious diseases spread by close contact in urban Blantyre, Malawi. Epidemics. 2022;40: 100590. doi:10.1016/j.epidem.2022.100590

14. Del Fava E, Adema I, Kiti MC, Poletti Pp, Merler S, Nokes DJ, et al. Individual’s daily behaviour and intergenerational mixing in different social contexts of kenya | scientific reports. Scientific Reports. 2021;11: 21589. doi:10.1038/s41598-021-00799-1

15. Miller PB, Zalwango S, Galiwango R, Kakaire R, Sekandi J, Steinbaum L, et al. Association between tuberculosis in men and social network structure in Kampala, Uganda. BMC infectious diseases. 2021;21: 1023. doi:10.1186/s12879-021-06475-z

16. Houben RMGJ, Lalli M, Sumner T, Hamilton M, Pedrazzoli D, Bonsu F, et al. TIME impact a new user-friendly tuberculosis (TB) model to inform TB policy decisions. BMC Medicine. 2016;14: 56. doi:10.1186/s12916-016-0608-4

17. Horton KC, White RG, Hoa NB, Nguyen HV, Bakker R, Sumner T, et al. Population benefits of addressing programmatic and social determinants of gender disparities in tuberculosis in viet nam: A modelling study. PLOS Global Public Health. 2022;2: e0000784. doi:10.1371/journal.pgph.0000784

18. Coussens AK, Zaidi SMA, Allwood BW, Dewan PK, Gray G, Kohli M, et al. Classification of early tuberculosis states to guide research for improved care and prevention: An international delphi consensus exercise. The Lancet Respiratory Medicine. 2024;12: 484–498. doi:10.1016/S2213-2600(24)00028-6

19. Stover J, McKinnon R, Winfrey B. Spectrum: A model platform for linking maternal and child survival interventions with AIDS, family planning and demographic projections. International Journal of Epidemiology. 2010;39: i7–i10. doi:10.1093/ije/dyq016

20. UN World Population Prospects. Population by select age groups - female. https://population.un.org/wpp/Download/Standard/Population/; 2024.

21. UN World Population Prospects. Population by select age groups - male. https://population.un.org/wpp/Download/Standard/Population/; 2024.

22. World Health Organization Global Tuberculosis programme. WHO TB burden estimates. https://www.who.int/teams/global-tuberculosis-programme/data; 2024.

23. World Health Orgaanization. Bacillus calmette-guérin (BCG) vaccination coverage. https://immunizationdata.who.int/pages/coverage/BCG.html?CODE=Global+KEN+NGA+UGA+MWI&YEAR=; 2024.

24. World Health Organization Global Tuberculosis programme. Treatment success rate: HIV-positive TB cases. https://www.who.int/data/gho/data/indicators/indicator-details/GHO/treatment-success-rate-hiv-positive-tb-cases; 2023.

25. WHO Global Tuberculosis programme. TB: Treatment success. https://www.who.int/data/gho/data/themes/topics/indicator-groups/indicator-group-details/GHO/tb-treatment-success; 2024.

26. Richards AS, Sossen B, Emery JC, Horton KC, Heinsohn T, Frascella B, et al. Quantifying progression and regression across the spectrum of pulmonary tuberculosis: A data synthesis study. The Lancet Global Health. 2023;11: e684–e692. doi:10.1016/S2214-109X(23)00082-7

27. Horton KC, Richards AS, Emery JC, Esmail H, Houben RMGJ. Reevaluating progression and pathways following mycobacterium tuberculosis infection within the spectrum of tuberculosis. Proceedings of the National Academy of Sciences. 2023;120: e2221186120. doi:10.1073/pnas.2221186120

28. Ragonnet R, Flegg JA, Brilleman SL, Tiemersma EW, Melsew YA, McBryde ES, et al. Revisiting the natural history of pulmonary tuberculosis: A bayesian estimation of natural recovery and mortality rates. Clinical Infectious Diseases. 2021;73: e88–e96. doi:10.1093/cid/ciaa602

29. Emery JC, Dodd PJ, Banu S, Frascella B, Garden FL, Horton KC, et al. Estimating the contribution of subclinical tuberculosis disease to transmission: An individual patient data analysis from prevalence surveys. Kana BD, editor. eLife. 2023;12: e82469. doi:10.7554/eLife.82469

30. Law I, Floyd K, Group the ATPS. National tuberculosis prevalence surveys in africa, 20082016: An overview of results and lessons learned. Tropical Medicine & International Health. 2020;25: 1308–1327. doi:10.1111/tmi.13485

31. WHO Global Tuberculosis programme. WHO TB incidence estimates disaggregated by age group, sex, and risk factor. https://www.who.int/teams/global-tuberculosis-programme/data; 2024.

32. WHO Global Tuberculosis programme. Case notifications. https://www.who.int/teams/global-tuberculosis-programme/data; 2024.

33. National Tuberculosis L, Lung Disease Program R of K Ministry of Health. Kenya tuberculosis prevalence survey 2016. Nairobi, Kenya; 2018. Available: https://www.nltp.co.ke/survey-reports-2/

34. Ministry of Health NTCP. Technical report: Malawi tuberculosis prevalence survey (2013–2014). 2016.

35. Department of Public Health FR of N. Report of the first national TB prevalence survey 2012, nigeria. 2014.

36. Public Health MUS of. Report on the population-based survey of prevalence of tuberculosis disease in uganda 2014–15. Kampala, Uganda; 2016. Available: https://library.health.go.ug/index.php/communicable-disease/tuberculosis/uganda-national-tuberculosis-prevalence-survey-2014-2015-survey

37. Iskauskas A, Vernon I, Goldstein M, Scarponi D, McCreesh N, McKinley TJ, et al. Emulation and history matching using the hmer package. Journal of Statistical Software. 2024;109: 1–48. doi:10.18637/jss.v109.i10

38. RStudio Team. RStudio: Integrated development environment for r. Boston, MA: RStudio, PBC.; 2020. Available: http://www.rstudio.com/

39. R Core Team. R: A language and environment for statistical computing. Vienna, Austria: R Foundation for Statistical Computing; 2024. Available: https://www.R-project.org/

40. Turyahabwe S, Bamuloba M, Mugenyi L, Amanya G, Byaruhanga R, Fry Imoko J, et al. Community tuberculosis screening, testing and care, uganda. Bulletin of the World Health Organization. 2024;102: 400–409. doi:10.2471/BLT.23.290641

41. Ogoamaka C, Bethrand O, Lotanna U, Chidubem O, Sani U, Nkiru N, et al. The TB surge intervention: An optimized approach to TB case-finding in nigeria. Public Health Action. 2023;13: 136–141. doi:10.5588/pha.23.0039

42. Marks GB, Nguyen NV, Nguyen PTB, Nguyen T-A, Nguyen HB, Tran KH, et al. Community-wide Screening for Tuberculosis in a High-Prevalence Setting. The New England Journal of Medicine. 2019;381: 1347–1357. doi:10.1056/NEJMoa1902129

43. Jesus GS, Gestal PFPS, Silva AF, Cavalcanti DM, Lua I, Ichihara MY, et al. Effects of conditional cash transfers on tuberculosis incidence and mortality according to race, ethnicity and socioeconomic factors in the 100 million brazilian cohort. Nature Medicine. 2025;31: 653–662. doi:10.1038/s41591-024-03381-0

44. Bhargava A, Bhargava M, Meher A, Benedetti A, Velayutham B, Teja GS, et al. Nutritional supplementation to prevent tuberculosis incidence in household contacts of patients with pulmonary tuberculosis in india (RATIONS): A field-based, open-label, cluster-randomised, controlled trial. The Lancet. 2023;402: 627–640. doi:10.1016/S0140-6736(23)01231-X

45. Chikovore J, Hart G, Kumwenda M, Chipungu G, Desmond N, Corbett EL. TB and HIV stigma compounded by threatened masculinity: Implications for TB health-care seeking in malawi. The International Journal of Tuberculosis and Lung Disease. 2017;21: S26–S33. doi:10.5588/ijtld.16.0925

46. Nidoi J, Pulford J, Wingfield T, Rachael T, Ringwald B, Katagira W, et al. Finding the missing men with tuberculosis: A participatory approach to identify priority interventions in uganda. Health Policy and Planning. 2025;40: 1–12. doi:10.1093/heapol/czae087

47. Ugwu C, Adekeye O, Ringwald B, Thomson R, Chijioke-Akaniro O, Anyaike C, et al. Co-creation of a gender responsive TB intervention in nigeria: A researcher-led collaborative study. BMC Health Services Research. 2025;25: 63. doi:10.1186/s12913-025-12241-7

48. Mwirigi N, Wandia R, Oira D, Kathure I, Nyaboga LM. Breaking barriers: Targeting TB in the workplace for men. 55th union world conference on lung health, bali, 2024. 2024. pp. S583–S584.

49. Karamagi E, Sensalire S, Muhire M, Kisamba H, Byabagambi J, Rahimzai M, et al. Improving TB case notification in northern uganda: Evidence of a quality improvement-guided active case finding intervention. BMC Health Services Research. 2018;18: 954. doi:10.1186/s12913-018-3786-2

50. UN Women Africa. Women breaking barriers in the fish farming industry in uganda. 2023. Available: https://africa.unwomen.org/en/stories/feature-story/2023/08/women-breaking-barriers-in-the-fish-farming-industry-in-uganda

51. World Prison Brief. Uganda | world prison brief. 2024. Available: https://www.prisonstudies.org/country/uganda

52. Dowden J, Mushamiri I, McFeely E, Apat D, Sacks J, Amor YB. The impact of “male clinics” on health-seeking behaviors of adult men in rural kenya. PLOS ONE. 2019;14: e0224749. doi:10.1371/journal.pone.0224749

53. Phiri MM, Makepeace E, Nyali M, Kumwenda M, Corbett E, Fielding K, et al. Improving pathways to care through interventions cocreated with communities: A qualitative investigation of men’s barriers to tuberculosis care-seeking in an informal settlement in blantyre, malawi. BMJ Open. 2021;11: e044944. doi:10.1136/bmjopen-2020-044944

54. Költringer FA, Annerstedt KS, Boccia D, Carter DJ, Rudgard WE. The social determinants of national tuberculosis incidence rates in 116 countries: A longitudinal ecological study between 20052015. BMC Public Health. 2023;23: 337. doi:10.1186/s12889-023-15213-w

55. Gilmore AB, Fabbri A, Baum F, Bertscher A, Bondy K, Chang H-J, et al. Defining and conceptualising the commercial determinants of health. The Lancet. 2023;401: 1194–1213. doi:10.1016/S0140-6736(23)00013-2

56. Calderwood CJ, Marambire ET, Larsson L, Banze D, Mfinanga A, Nhamuave C, et al. HIV, malnutrition, and noncommunicable disease epidemics among tuberculosis-affected households in east and southern africa: A cross-sectional analysis of the ERASE-TB cohort. PLOS Medicine. 2024;21: e1004452. doi:10.1371/journal.pmed.1004452

57. Wong EB, Olivier S, Gunda R, Koole O, Surujdeen A, Gareta D, et al. Convergence of infectious and non-communicable disease epidemics in rural south africa: A cross-sectional, population-based multimorbidity study. The Lancet Global Health. 2021;9: e967–e976. doi:10.1016/S2214-109X(21)00176-5

58. Falzon D, Zignol M, Bastard M, Floyd K, Kasaeva T. The impact of the COVID-19 pandemic on the global tuberculosis epidemic. Frontiers in Immunology. 2023;14: 1234785. doi:10.3389/fimmu.2023.1234785

59. Kenya Ministry of Health, National Tuberculosis, Leprosy and Lung Disease Programme. Kenya national strategic plan for 2023/24 to 2027/28. 2023. Available: https://nltp.co.ke/national-strategic-plan-2019-2023-3/

60. Smith JP, Oeltmann JE, Hill AN, Tobias JL, Boyd R, Click ES, et al. Characterizing tuberculosis transmission dynamics in high-burden urban and rural settings. Scientific Reports. 2022;12: 6780. doi:10.1038/s41598-022-10488-2

