## Supplementary material for "Population level impact of increasing tuberculosis treatment coverage and addressing determinants of risk in men: a modelling study in Kenya, Malawi, Nigeria, and Uganda": S1 Appendix

### Supplementary Materials

### 1 Model Structure

#### 1.1 Tuberculosis model

We developed a sex-stratified dynamic compartmental model of *M.tb* transmission and tuberculosis (TB) disease, adapted from previously published models.[1–3] The core TB model has four states: susceptible, infection, asymptomatic bacteriologically confirmed disease, and symptomatic bacteriologically confirmed disease, as shown in Figure 1. Unlike in other variants of the model, disease states are split by symptoms rather than smear status. [2–4]

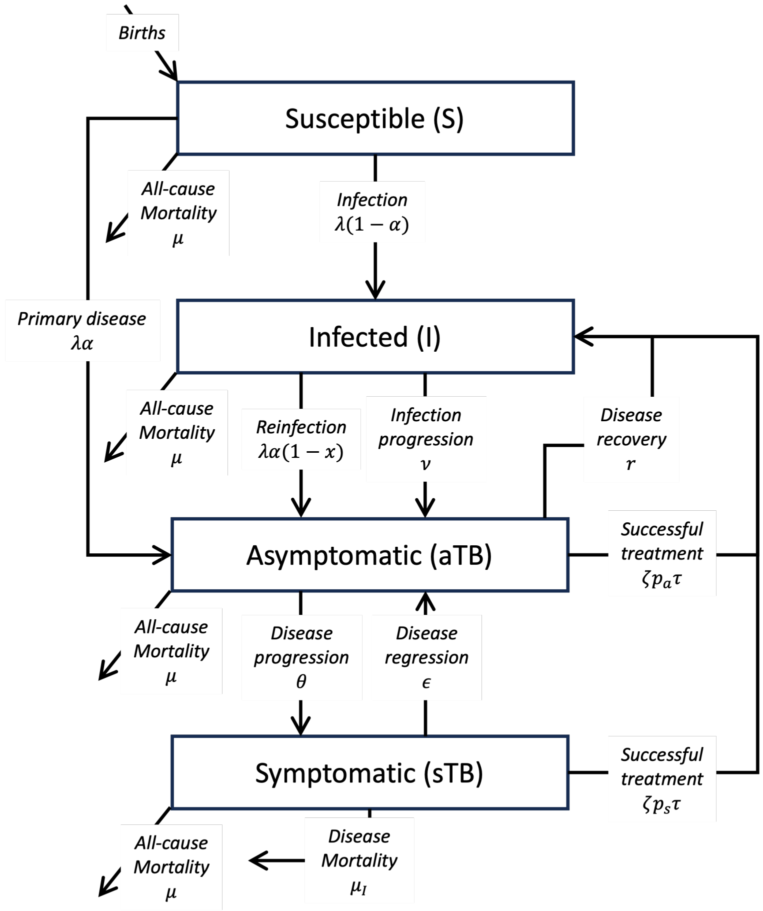

*Figure 1: The underlying TB model structure*

Individuals enter the susceptible compartment (S) at birth and are at risk of all-cause mortality throughout the model.

Susceptible individuals (S) have a force of infection, $\lambda$. A proportion, $\alpha$, of individuals progress directly to asymptomatic disease, and the rest, $(1-\alpha)$, remain infected with *M.tb*. Infected individuals haven an annual rate of progression, $\nu$, along with a rate of progression through reinfection, at a rate $\lambda\alpha(1-x)$, where $x$ is the protection against progression to disease upon reinfection, provided by previous infections.

Individuals with asymptomatic disease progress to symptomatic disease at a rate $\theta$, and they naturally clear disease at a rate $r$. Individuals with symptomatic disease regress to asymptomatic disease at a rate $\epsilon$ and die from disease at a rate $\mu_I$.

Individuals who recover from TB disease return to the infected compartment, where they are at risk of reinfection and natural progression as described above. Natural recovery is only possible from asymptomatic disease, as described above, but recovery of disease through successful treatment is possible from both disease stages.

The infection and disease compartments (I, aTB, and sTB) are stratified by the treatment history, with treatment naive indicated by a subscript $N$ and previously treated indicated by a subscript $P$. Individuals move from the treatment naive stratum to the previously treated stratum after any treatment, whether successful or unsuccessful, as shown in Figure 2 and detailed in previous work. [1,5]

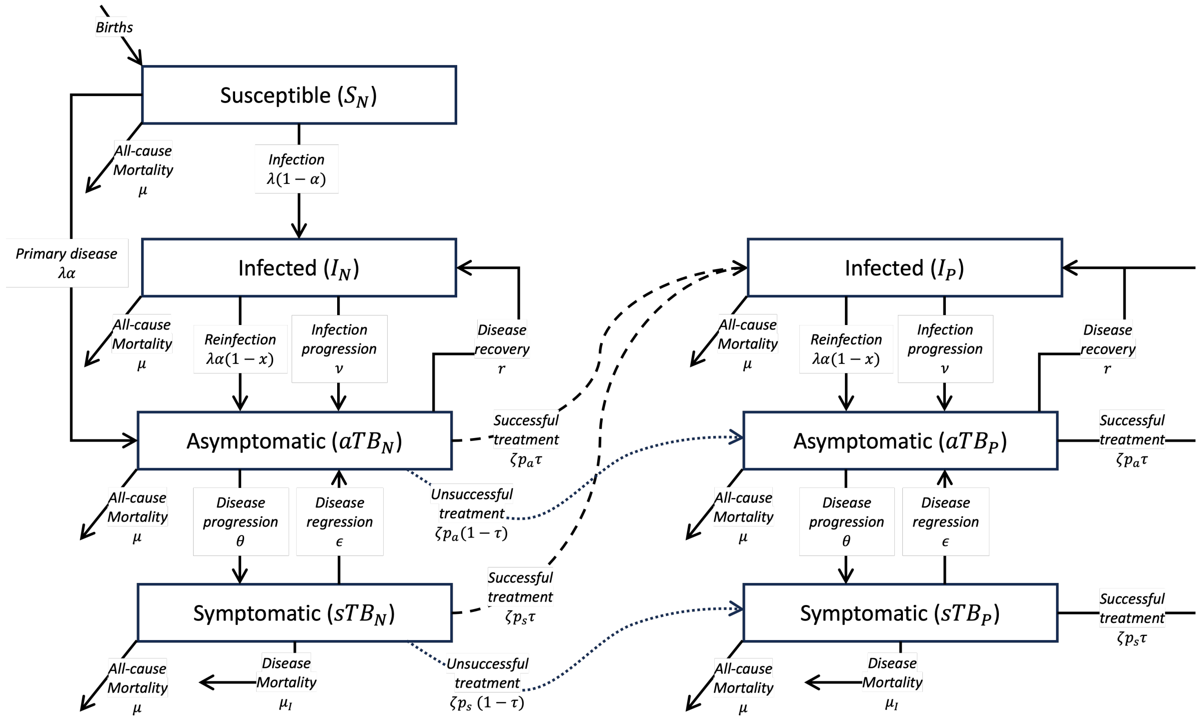

*Figure 2: The TB treatment model structure*

The full model is stratified by sex (male and female), age in fifteen-year age groups (0 - 14, 15 - 29, ... 60 - 74, ≥ 75), and HIV status (positive and negative), with the HIV-positive stratum further stratified by duration of anti-retroviral therapy (ART) (none, 0 - 6 months, 7 - 12 months, >12 months)

#### 1.2 Demographic model

The demographic model is designed to reflect the demographic projection model (DemProj) of the Spectrum software suite.[6] The demographic model accounts for births, migration, and all-cause mortality. The population is split across the two sex strata (male and female) and 17 age strata split into five-year age groups (0 - 4, 5 - 9, ..., 75 - 79, ≥ 80). Sex is assumed constant throughout, but individuals age up between the different age groups. For this model we have combined the age groups to form 6 strata in fifteen-year age groups (0 - 14, 15 - 29, ... 60 - 74, ≥ 75).

Births are modelled using the crude birth rate from the United Nations (UN) Population Division estimates, whilst also acknowledging the sex distribution of new births, both sourced from UN World Population Prospects estimates.[7] New births are added to the susceptible compartment of the TB model and aging is modelled such that one-fifteenth of the population of each fifteen-year age group moves to the subsequent age group each year.

Migration is calculated by age and sex for each year according to adjusted DemProj estimates.[6] Spectrum provides the annual net migration by gender and the percentage distribution of migration in five-year age groups by gender.[6] The percentages are then summed over three five-year age groups to give the percentage for one fifteen-year age group, and the raw migration numbers are split across the ages by the appropriate percentages. Migration is considered independent of TB or HIV disease status and migrants are distributed across disease states based on the relative size of each compartment.

All-cause mortality is modelled using rates derived from UN Population Division life tables.[7] This is calculated using the proportion of cohort survival over 15 years (i.e., the number of survivors at year 15 compared to the number of survivors at year 0). As TB and HIV deaths are included in background mortality rates, duplicate TB- and HIV-associated deaths are removed at each time point.

#### 1.3 HIV model

The HIV model has been simplified from previous work. [1,5] Individuals are either categorised as HIV-negative or HIV-positive. Those who are HIV-positive are further stratified into four stages of ART duration: none, 0 - 6 months, 7 - 12 months, >12 months). HIV incidence, ART initiation and duration, and HIV-associated mortality are unchanged from previous works, with the exception of the progression through CD4-count categories, as described elsewhere.[1,5]

### 2 Model equations

**Susceptible**

$$\frac{d\mathbf{S}_{g,a,h,k,t}}{dt} =$$

$$-\lambda_{g,a,t} \times \mathbf{S}_{g,a,h,k,t}$$

**Infection (never previously treated)**

$$\frac{d\mathbf{I}_{N_{g,a,h,k,t}}}{dt} =$$

$$\begin{aligned}
\left(\lambda_{g,a,t}\left(1-\alpha_{a,h,k,t}\right)\right) \times &\quad \mathbf{S}_{g,a,h,k,t}\\
-\left(\lambda_{g,a,t}\alpha_{a,h,j,t}\left(1-x_{a,h,k}\right)+\nu_{a,h,k,t}\right) \times &\quad \mathbf{I}_{N_{g,a,h,k,t}}\\
+r_h \times &\quad \mathbf{aTB}_{N_{g,a,h,k,t}}
\end{aligned}$$

**Asymptomatic (never previously treated)**

$$\frac{d\mathbf{aTB}_{N_{g,a,h,k,t}}}{dt} =$$

$$\begin{aligned}
\lambda_{g,a,t}\alpha_{a,h,k,t} \times &\quad \mathbf{S}_{g,a,h,k,t}\\
+\left(\lambda_{g,a,t}\alpha_{a,h,k,t}\left(1-x_{a,h,k}\right)+\nu_{a,h,k,t}\right) \times &\quad \mathbf{I}_{N_{g,a,h,k,t}}\\
-\left(\theta_h + r_h + \zeta p_{a,d=\mathbf{aTB},g,k}\tau_{h,k,t}\right) \times &\quad \mathbf{aTB}_{N_{g,a,h,k,t}}\\
+\epsilon_h \times &\quad \mathbf{sTB}_{N_{g,a,h,k,t}}
\end{aligned}$$

**Symptomatic (never previously treated)**

$$\frac{d\mathbf{sTB}_{N_{g,a,h,k,t}}}{dt} =$$

$$\begin{aligned}
\left(\theta_h\right) \times &\quad \mathbf{aTB}_{N_{g,a,h,k,t}}\\
-\left(\epsilon_h + \mu_{I_{a,h,k}} + \zeta p_{a,d=\mathbf{sTB},g,k}\tau_{h,k,t}\right) \times &\quad \mathbf{sTB}_{N_{g,a,h,k,t}}
\end{aligned}$$

**Infected (previously treated)**

$$\frac{d\mathbf{I}_{P_{g,a,h,k,t}}}{dt} =$$

$$\begin{aligned}
-\left(\lambda_{g,a,t}\alpha_{a,h,j,t}\left(1-x_{a,h,k}\right)+\nu_{a,h,k,t}\right) \times &\quad \mathbf{I}_{P_{g,a,h,k,t}}\\
\left(\zeta p_{a,d=\mathbf{aTB},g,k}\tau_{h,k,t}\right) \times &\quad \mathbf{aTB}_{N_{g,a,h,k,t}}\\
+\left(\zeta p_{a,d=\mathbf{aTB},g,k}\tau_{h,k,t} + r_h\right) \times &\quad \mathbf{aTB}_{P_{g,a,h,k,t}}\\
+\left(\zeta p_{a,d=\mathbf{sTB},g,k}\tau_{h,k,t}\right) \times &\quad \mathbf{sTB}_{N_{g,a,h,k,t}}\\
+\left(\zeta p_{a,d=\mathbf{sTB},g,k}\tau_{h,k,t}\right) \times &\quad \mathbf{sTB}_{P_{g,a,h,k,t}}
\end{aligned}$$

**Asymptomatic (previously treated)**

$$\frac{d\mathbf{aTB}_{P_{g,a,h,k,t}}}{dt} =$$

$$+\left(\lambda_{g,a,t}\alpha_{a,h,k,t}\left(1 - x_{a,h,k}\right) + \nu_{a,h,k,t}\right) \times \quad \mathbf{I}_{P_{g,a,h,k,t}}$$

$$-\left(\theta_h + r_h + \zeta p_{a,d=\mathbf{aTB},g,k}\tau_{h,k,t}\right) \times \quad \mathbf{aTB}_{P_{g,a,h,k,t}}$$

$$+\zeta p_{a,d=\mathbf{aTB},g,k}\left(1 - \tau_{h,k,t}\right)) \times \quad \mathbf{aTB}_{N_{g,a,h,k,t}}$$

$$+\epsilon_h \times \quad \mathbf{sTB}_{P_{g,a,h,k,t}}$$

**Symptomatic (previously treated)**

$$\frac{d\mathbf{sTB}_{P_{g,a,h,k,t}}}{dt} =$$

$$(\theta_h) \times \quad \mathbf{aTB}_{P_{g,a,h,k,t}}$$

$$-\left(\epsilon_h + \mu_{I_{a,h,k}} + \zeta p_{a,d=\mathbf{sTB},g,k}\tau_{h,k,t}\right) \times \quad \mathbf{sTB}_{P_{g,a,h,k,t}}$$

$$+\zeta p_{a,d=\mathbf{sTB},g,k}\left(1 - \tau_{h,k,t}\right)) \times \quad \mathbf{sTB}_{N_{g,a,h,k,t}}$$

### 3 Model parameters

#### 3.1 Force of infection

The force of infection ($\lambda_{g,a,t}$) is a time-dependent parameter, specific to sex ($g$), age ($a$), and time ($t$). It acknowledges the heterogeneous patterns of social contacts between men, women, and children, as well as a constant sex-specific relative risk of infection attributable to biological and/or social factors. The force of infection is as follows:

$$\lambda_{g,a,t} = (z)(cscal)\left(RR_{sex_{g,a}}\right) \times \left(c_{M_{g,a}} \frac{\mathbf{sTB}_{g=M,a\geq15,t} + q\mathbf{aTB}_{g=M,a\geq15,t}}{T_t} + c_{F_{g,a}} \frac{\mathbf{sTB}_{g=F,a\geq15,t} + q\mathbf{aTB}_{g=F,a\geq15,t}}{T_t} + c_{C_{g,a}} \frac{\mathbf{sTB}_{a=0-14,t} + q\mathbf{aTB}_{a=0-14,t}}{T_t}\right)$$

for gender $g$, age $a$, and time step $t$. There are three sex-specific contact parameters: $c_{M_{g,a}}$, $c_{F_{g,a}}$, and $c_{C_{g,a}}$ and these are the average number of contacts with adult men, adult women, and children respectively for the given age group and sex. These are each multiplied by the sum of the number of people of that respective age and sex with symptomatic disease ($I_{M,t}$, $I_{F,t}$, $I_{C,t}$) and the product of the relative infectiousness for asymptomatic disease, $q$, and the number of people of that respective age and sex with asymptomatic disease ($N_{M,t}$, $N_{F,t}$, $N_{C,t}$) and divided through by the total population $T_t$. The sum of the sex-specific component is then further multiplied by the probability of *Mtb* transmission per respiratory contact between an infectious and an uninfected individual ($z$) , a scaling factor to account for uncertainty in the total number of contacts ($cscal$) and the relative risk of infection attributable to sex- or gender-based risks ($RR_{sex_{g,a}}$). Prior ranges and data sources for parameters are described below in Table 1 and Table 5.

*Table 1: Prior ranges and data sources for parameters related to the force of infection*

| Parameter | Description | Prior range or set value | References |
|---|---|---|---|
| q | Relative infectiousness of asymptomatic disease compared to symptomatic disease | 0.62 - 1 | [8] |
| z | Probability of transmission per respiratory contact between infectious and uninfected individuals | 0.1 | [1,9–11] |
| cscal | Scaling factor to adjust the total number of contacts | 6 - 25 | Assumption |

###### 3.1.1.1 *Adjustments for heterogeneous mixing*

The force of infection acknowledges heterogeneous mixing by sex and age by incorporating the average number of contacts between men (male, aged $\geq$ 15 years), women (female, $\geq$ 15 years), and children (both sexes, age $<$ 15 years). Estimates for the average number of contacts were, where possible, taken from social contact studies. The number of close contacts within a 24-hour period between each participant and each contact group (men, women, boys, and girls) and then we smoothed the matrix to ensure symmetry.[12] Estimates for the average number of contacts in Kenya are based on a social contact survey conducted in urban and rural areas in Mombassa and Kilifi counties in 2015 and the final smoothed contact matrix is shown in Table 2. [13]

*Table 2: The contact matrix used for the Kenya model*

| Participants | Average number of close contacts | | |
|---|---|---|---|
| | Men | Women | Children |
| Men | 4.48 | 3.12 | 1.97 |
| Women | 1.70 | 3.57 | 2.17 |
| Children | 1.06 | 2.15 | 5.39 |

A similar survey was conducted in Blantyre, Malawi, and the resulting contact matrix is presented in Table 3. [14]

*Table 3: The contact matrix used for the Malawi model*

| Participants | Average number of close contacts | | |
|---|---|---|---|
| | Men | Women | Children |
| Men | 3.36 | 1.78 | 3.81 |
| Women | 1.74 | 3.21 | 3.90 |
| Children | 1.65 | 1.72 | 8.40 |

We found no single social contact survey that covered both sexes and adults and children for either Nigeria or Uganda, so we amalgamated all social contact surveys from countries in Africa to create a more generic matrix for the two countries, as shown in Table 4. [13–18]

*Table 4: The contact matrix used for both the Nigeria and the Uganda models*

| Participants | Average number of close contacts | | |
|---|---|---|---|
| | Men | Women | Children |
| Men | 4.03 | 3.02 | 1.40 |
| Women | 2.57 | 4.17 | 1.90 |
| Children | 2.39 | 3.80 | 8.70 |

###### *3.1.1.2 Adjustments for sex- or gender-based risks*

The relative risk of infection attributable to sex- or gender-based risks $\left(RR_{sex_{g,a}}\right)$ reflects the accumulation of risks through biological, social, economic, and health determinants that may contribute to men's increased risk of *M.tb* infection relative to women and children.[19] In this model, the factors that contribute to these differences are considered constant over time, and the prior for the relative risk of infection in men, relative to women and children is a range of between 1 and 2.5 (or 3 in Uganda), as shown in Table 5.

*Table 5: Prior ranges and data sources for relative risks of M.tb infection attributable to sex- or gender-based risks*

| Parameter | Description | Prior range or set value | References |
|---|---|---|---|
| $RR_{sex_{g=M,a\geq15}}$ | Relative risk of infection attributable to additional sex- or gender-based risks in men | 1 - 2.5<br>(10 in Uganda) | Assumption |
| $RR_{sex_{g=F,a\geq15}}$ | Relative risk of infection attributable to additional sex- or gender-based risks in women | 1 | Assumption |
| $RR_{sex_{g,a=0-14}}$ | Relative risk of infection attributable to additional sex- or gender-based risks in children | 1 | Assumption |

#### 3.2 Progression to disease

##### 3.2.1 Rapid progression to disease

A proportion of all new infections progress direct to asymptomatic disease ($\alpha_{a,h,k,t}$) and this is a time-dependent parameter specific to age, HIV status, and ART duration.

The proportion of new infections progressing directly to asymptomatic disease in HIV-negative individuals is defined as:

$$\alpha_{a,t} = \alpha_a RR_{BCG_{a,t}}$$

for age $a$, and time-step $t$, where $\alpha_a$ is the base proportion of new infections progressing directly to asymptomatic disease and $RR_{BCG_a}$ is the relative risk of progression from *M.tb* infection to disease attributable to BCG vaccination. The baseline values for $\alpha_a$ are contained in Table 6.

*Table 6: Prior ranges and data sources for the proportion of infections that progress rapidly to asymptomatic disease*

| Parameter | Description | Prior range or set value | References |
|---|---|---|---|
| $\alpha_{a\geq15}$ | Base proportion of rapid progression to asymptomatic disease in adults | 0.08 - 0.15+ | [1, 5, 20–22] |
| $\alpha_{a=0-14}$ | Base proportion of rapid progression to asymptomatic disease in adults | 0.08 - 0.15 | [1, 5, 21–23] |

$^{+}\alpha_{a\geq15}$ must be greater than $\alpha_{a=0-14}$

##### *3.2.1.1 Adjustments for BCG vaccination*

The relative risk of progression from *M.tb* infection to disease attributable to protection from BCG vaccination is defined as follows:

$$RR_{BCG_{a,t}} = \left(1 - BCG_{cov_t} \times BCG_{eff_a}\right)$$

for age $a$ and time $t$, where $BCG_{cov_t}$ is the coverage of BCG vaccination in infants over time, and $BCG_{eff_a}$ is the efficacy of BCG vaccination.

Coverage of BCG vaccination in infants for each country is shown in Figure 3. For each country, vaccine coverage is assumed to increase linearly from 1970 until the data becomes available - 1983 for Kenya, 1981 for the other three countries. The data used are the WHO/UNICEF Estimates of National Immunization Coverage which provides the percentage of live births who have received one dose of the BCG vaccine in a given year. [24] The assumed coverage from 2023 to 2050 represents the likely achievable coverage based om the coverage for the years prior to 2023. BCG coverage is assumed to be equal by sex as a 2009 WHO Expanded Programme on Immunizations survey found similar coverage for boys and girls. [25]

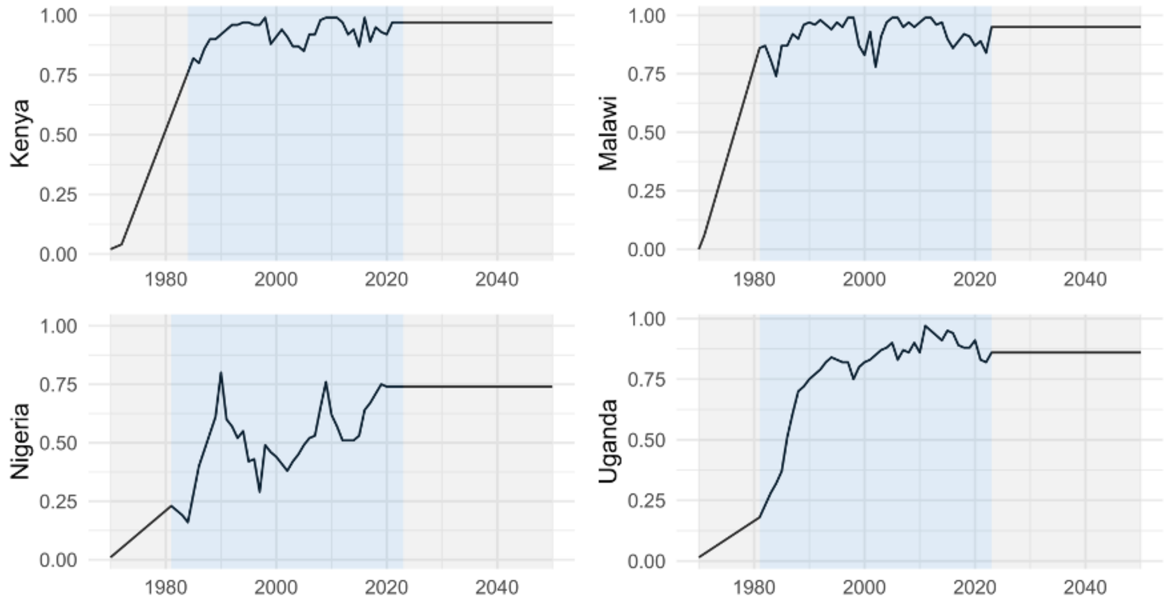

*Figure 3: The BCG vaccination coverage in infants over time from 1970 to 2050. Data is available for the years shaded in blue, and the years shaded in grey are assumed coverage.*

The efficacy of BCG vaccination is assumed to vary by age, as shown in Table 7

*Table 7: Prior ranges and data sources for the efficacy of BCG vaccination*

| Parameter | Description | Prior range or set value | References |
|---|---|---|---|
| $BCG_{eff_{a\geq15}}$ | Efficacy of BCG vaccination in ages ≥ 15 | 0 | Assumption |
| $BCG_{eff_{a=0-14}}$ | Efficacy of BCG vaccination in ages 0 - 14 years | 0.39 - 0.72 | [1, 5, 26] |

##### *3.2.1.2 HIV-positive and ART strata*

The proportion of new infections developing primary disease is adjusted for individuals with HIV as follows:

$$\alpha_h = \alpha \times RR_{1_\alpha}$$

where $\alpha$ is the baseline parameter for rapid progression, and $RR_{1_\alpha}$ is the risk ratio of progression for individuals with HIV. The value for the risk ratio is in Table 8.

ART reduces the effect of HIV infection on rapid progression to disease, bringing the value closer to that for an individual without HIV:

$$\alpha_{h,k}^A = \max(\alpha_h^H \times (1 - ART_k), \alpha)$$

where $\alpha$ is the baseline parameter for rapid progression and $RR_{1_\alpha}$ is the relative risk of progression for individuals with HIV. Subscripts $h$ and $k$ are the HIV status and ART duration respectively, and superscripts $A$ and $H$ refer to HIV-positive individuals on ART and not on ART respectively. The values of these HIV- and ART-specific parameters are in Table 8.

*Table 8: Prior ranges and data sources for the risk ratio associated with being HIV-positive and the reduction of risk from ART*

| Parameter | Description | Prior range or set value | References |
|---|---|---|---|
| $RR_{1_\alpha}$ | The risk ratio for new infections rapidly progressing to asymptomatic disease due to HIV co-infection | 2.11 - 20 | [1,5,22,27] |
| $ART_{TB_{t<6}}$ | Protective effect of ART treatment against developing disease for treatment duration < 6 months | 0.16 - 0.27 | [5,28] |
| $ART_{TB_{t=6-12}}$ | Protective effect of ART treatment against developing disease for treatment duration 6 - 12 months | 0.30 - 0.73 | [5,28] |
| $ART_{TB_{t\geq 12}}$ | Protective effect of ART treatment against developing disease for treatment duration $\geq$ 12 months | 0.54 - 0.92$^+$ | [5,28] |

$^+ART_{TB_{t\geq 12}}$ must be greater than $ART_{TB_{t=6-12}}$

#### 3.2.2 Progression from infection

There is a fixed rate of progression from infection to disease, independent of time since infection, $\nu_{a,h,k}$, that is dependent on age, HIV status, and ART duration. The progression rate in HIV-negative individuals is $\nu_a$, dependent only on age. The relevant values for progression from infection are included in Table 9

##### *3.2.2.1 HIV-positive and ART strata*

The rate of progression to asymptomatic disease is adjusted for HIV in the same way as rapid progression in Section 3.2.1.2, using the parameter $RR_{1_\nu}$. The prior range and references for this parameter are included in Table 9.

The rate of progression to asymptomatic disease is adjusted for ART duration in the same way as rapid progression in Section 3.2.1.2, using the values described in Table 8.

References and

*Table 9: Prior ranges and data sources for the annual rate of progression to asymptomatic disease in HIV-negative individuals and the risk ratio associated with being HIV-positive*

| Parameter | Description | Prior range or set value | References |
|---|---|---|---|
| $\nu_{a\geq 15}$ | The annual rate of progression from infection to asymptomatic disease in ages $\geq 15$ | 0.0001 - 0.0025 | [1, 5, 20–22, 29] |
| $\nu_{a=0-14}$ | The annual rate of progression from infection to asymptomatic disease in ages 0 - 14 | 0.0001 - 0.0025 | [1, 5, 20–22, 29] |
| $RR_{1_\nu}$ | The risk ratio for infection progression or reactivation in HIV-positive individuals | 2.11 - 20 | [1, 5, 20, 22, 27, 30] |

##### 3.2.3 Reinfection

Previous infection confers some protection against reinfection, $x_{a,h,k}$, a constant specific to age, HIV status, and ART duration. The progression rate in HIV-negative individuals is $x_a$, dependent only on age. The relevant values for progression from infection are included in Table 10

###### *3.2.3.1 HIV-positive and ART strata*

The rate of progression to asymptomatic disease is adjusted for HIV in the same way as rapid progression in Section 3.2.1.2, using the parameter $RR_{1_x}$. The prior range and references for this parameter are included in Table 10.

The rate of progression to asymptomatic disease is adjusted for ART duration in the same way as rapid progression in Section 3.2.1.2, using the values described in Table 8.

*Table 10: Prior ranges and data sources for the annual rate of progression to asymptomatic disease in HIV-negative individuals and the risk ratio associated with being HIV-positive*

| Parameter | Description | Prior range or set value | References |
|---|---|---|---|
| $x_{a\geq 15}$ | Proportion of protection against progression to asymptomatic disease due to previous infection in adults | 0.37 - 0.9 | [1, 5, 20, 21, 31, 32] |
| $x_{a=0-14}$ | Proportion of protection against progression to asymptomatic disease due to previous infection in children | 0.37 - 0.9 | [1, 5, 20, 21, 31, 32] |
| $RR_{1_x}$ | The risk ratio for protection from reinfection in HIV-positive individuals | 2.11 - 20 | [1, 5, 20, 21, 32] |

#### 3.3 Progression through disease

Progression from asymptomatic to symptomatic disease occurs at a rate $\theta$. This parameters are only dependent on HIV status and are described in Table 11.

The rate of progression from asymptomatic to symptomatic disease is adjusted for HIV in the same way as rapid progression in Section 3.2.1.2, using the parameter $RR_{1_\theta}$. The prior range and references for this parameter are included in Table 11.

There are no adjustments for ART within the progression from asymptomatic to symptomatic disease.

*Table 11: Prior ranges and data sources for the annual rate of progression from asymptomatic disease to symptomatic disease in HIV-negative individuals and the risk ratio associated with being HIV-positive*

| Parameter | Description | Prior range or set value | References |
|---|---|---|---|
| $\theta$ | Rate of progression from asymptomatic to symptomatic disease in HIV-negative individuals | 0.56 - 0.94 | [2, 3, 5] |
| $RR_{1_\theta}$ | Risk ratio for progression from asymptomatic to symptomatic disease in HIV-positive individuals | 2.11 - 20 | [1, 5, 20, 22, 27, 30] |

#### 3.4 Regression through disease

Regression from symptomatic to asymptomatic disease is described by $\epsilon_h$, a parameter are only dependent on HIV status as described in Table 12.

The rate of regression from symptomatic to asymptomatic disease in HIV-positive individuals is not adjusted in the same way. Instead it has a prior range, as shown in Table 11, with a lower bound half that of the lower bound of $\epsilon_{h=HIV-}$ and a restriction that $\epsilon_{h=HIV+} < \epsilon_{h=HIV-}$.

There are no adjustments for ART within the regression from symptomatic to asymptomatic disease.

*Table 12: Prior ranges and data sources for the annual rate of regression from symptomatic disease to asymptomatic disease in HIV-negative individuals and HIV-positive individuals*

| Parameter | Description | Prior range or set value | References |
|---|---|---|---|
| $\epsilon_{h=HIV-}$ | Rate of regression from symptomatic to asymptomatic disease in HIV-negative individuals | 0.46 - 0.73 | [2, 3, 5] |

| | | | |
|---|---|---|---|
| $\epsilon_{h=HIV+}$ | Rate of regression from symptomatic to asymptomatic disease in HIV-positive individuals | 0.23 - 0.73⁺ | [2, 3, 5] |

⁺$\epsilon_{HIV}$ must be les than $\epsilon$

#### 3.5 Self-cure

Self-cure from disease, without treatment, occurs from asymptomatic disease, and returns individuals to infection. Self-cure is described by $r_h$, a time-independent parameter, dependent on HIV status, as shown in Table 13.

*Table 13: Prior ranges and data sources for the parameters describing self-cure from disease*

| Parameter | Description | Prior range or set value | References |
|---|---|---|---|
| $r_{h=HIV-}$ | Rate of self-cure from asymptomatic disease in HIV-negative individuals | 0.1 - 0.306 | [2, 3, 5, 21, 22, 33, 34] |
| $r_{h=HIV+}$ | Rate of self-cure from asymptomatic disease in HIV-positive individuals | 0.06 - 0.16⁺ | [5, 21, 32] |

⁺$r_{h=HIV+}$ must be less than $r_{h=HIV-}$

#### 3.6 Care cascade

The care cascade is represented by three separate values: access to care, $\zeta$, an adjustment of access to care for different strata, $p_{a,d,g,k}$, and treatment completion, $\tau_{h,k,t}$.

$$R_{a,d,g,h,k,t} = \zeta \times p_{a,d,g,k} \times \tau_{h,k,t}$$

##### 3.6.1 Access to care

Access to care is informed by the treatment coverage in each country, which is a country-specific, time-dependent parameter. Data for each country is available from 2000 onwards, and before 2000 we assumed the growth of access to care based on the change in treatment coverage in the available time period from 2000 to 2022.[35] For Kenya we chose an approximately S-shaped curve to bring treatment coverage up from a low starting point, but then to stabilise between 2000 and 2015. Malawi had a more consistent change with time, so we chose a linear increase from a similarly low starting point. Nigeria had very low treatment coverage in 2000, and so we assumed a flat but low coverage for all the years prior to 2000. Uganda had consistently high treatment coverage from 2000 to 2020, with an increase in 2022, and prior to 2000 we assumed a low flat coverage from until 1990 followed by a gradually increasing increase to meet the trajectory of coverage in 2000. All these trends can be seen in Figure 4. Following 2022, for each country we have assumed an increase in-line with the expectations of Kenya's National Strategic Plan.[36]

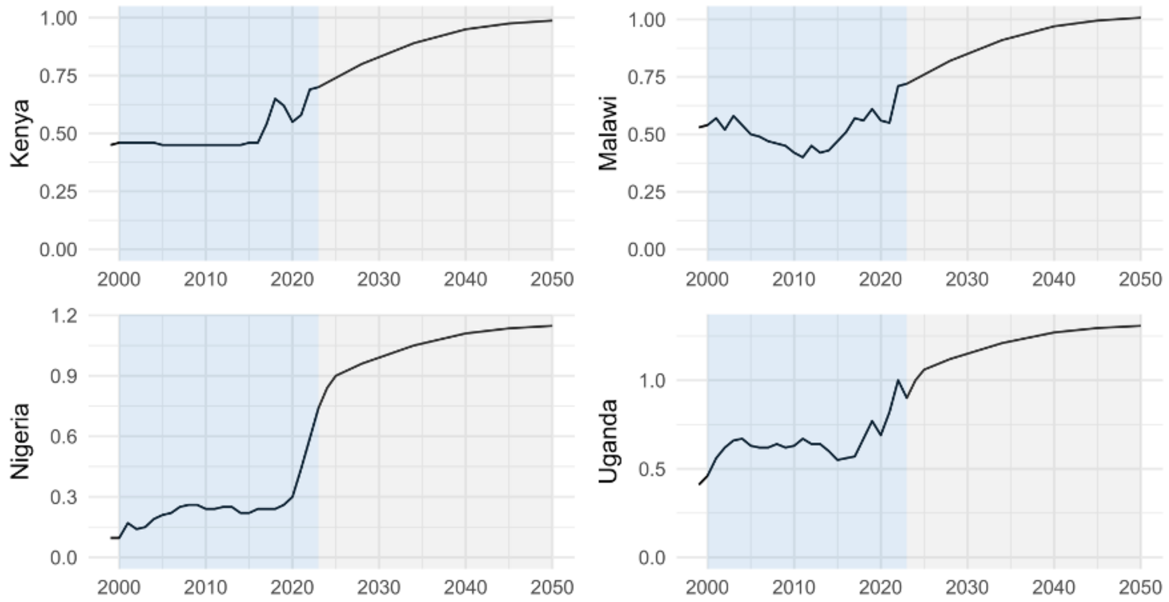

*Figure 4: Access to care over time, with assumed access in the grey area and treatment coverage data in the blue area.*

##### *3.6.1.1 Adjustments to treatment coverage*

Access to care varies across the population, and by using treatment coverage to estimate this parameter, we have introduced further uncertainty. To account for this, we introduced time-independent modifiers to the access to care value dependent on the demographic and disease state group, $p_{a,d,h,k}$. These are then multiplied with the access to care parameter to represent the overall treatment coverage at that time for the given subgroup. The values for these modifiers are in Table 14.

$$p_{a,d,h,k} = 1 + \left( \begin{cases} p_a & \text{if } a < 15 \\ p_k & \text{if on ART} \\ p_d \times p_g & \text{otherwise} \end{cases} \right)$$

*Table 14: Prior ranges for the demographic- and disease-specific modifiers to treatment coverage*

| Parameter | Description | Prior range or set value | References |
|---|---|---|---|
| $p_{d=\mathbf{sTB}}$ | Adjustment to treatment coverage for symptomatic disease in HIV-negative individuals and HIV-positive individuals who are not on ART | $0$ | Assumption |
| $p_{d=\mathbf{aTB}}$ | Adjustment to treatment coverage for asymptomatic disease in HIV-negative individuals and HIV-positive individuals who are not on ART | $-1 - 0$ | Assumption |

| | | | |
|---|---|---|---|
| $p_{k=ART}$ | Adjustment to treatment coverage for symptomatic and asymptomatic disease in HIV-positive individuals who are receiving ART and have been for any duration | $0 - 1$ | Assumption |
| $p_{k=noART}$ | Adjustment to treatment coverage for symptomatic and asymptomatic disease in HIV-positive individuals who are not receiving ART | $0$ | Assumption |
| $p_{g=M}$ | Adjustment to treatment coverage for men, regardless of disease state | –0.5 – 0.5 (–0.75 – 0.5 in Uganda)⁺ | Assumption |
| $p_{g=F}$ | Adjustment to treatment coverage for women, regardless of disease state | –0.5 – 0.5 (–0.5 – 1 in Uganda)⁺ | Assumption |
| $p_{a=0-14}$ | Adjustment to treatment coverage for children, regardless of disease state | $-1 - 0$ | Assumption |
| $p_{a\geq 15}$ | Adjustment to treatment coverage for adults, regardless of disease state | $0$ | Assumption |

⁺$p_{g=M}$ must be less than $p_{g=F}$

##### 3.6.2 CAST in Uganda

From 2022, Uganda have conducted a twice-yearly active case finding strategy that significantly increased their case-notification numbers. The first two rounds in 2022 screened over 1.25 million and over 5 million people respectively, creating an impact much larger than that of the general care cascade pathway in the model.[37] We therefore adjusted the model with further additions to the demographic-specific modifiers. As this is community-based screening, people on ART are no more likely to be screened than anyone else, unlike in classic treatment circumstances where people on ART are regularly attending clinics.[37] As this programme was only focussed on symptom screening, there was also no adjustment for asymptomatic disease.[37] The case notifications, between 2021 (before CAST was implemented) and 2022 (the first year of CAST), increased by 38% in women, 29% in men, and 3% in children.[35] To account for this, adjustments to the access to care were included for men, women, and children, from 2022 onwards with the adjustment to the care cascade equation as below, and the prior values for $f_{a,g}$ in Table 15.

$$R_{a,d,g,h,k,t} = \zeta \times (p_{a,d,g,k} + f_{a,g}) \times \tau_{h,k,t}$$

*Table 15: Prior ranges for the parameters to adjust the care cascade to represent the CAST programme in Uganda*

| Parameter | Description | Prior range or set value | References |
|---|---|---|---|
| $f_{a=0-14}$ | Adjustment to care cascade for children | 0 - 1.5 | Assumption |
| $f_{a\geq 15,g=M}$ | Adjustment to care cascade for men | 0 - 2 | Assumption |
| $f_{a\geq 15,g=F}$ | Adjustment to care cascade for women | 1 - 4 | Assumption |

#### 3.7 Treatment completion

Treatment completion is a time-dependent parameter. Data is available on treatment completion for HIV-negative individuals from 1995 to 2022, and for HIV-positive individuals it is available from 2007 in Kenya, 2015 in Malawi, 2009 in Nigeria, and 2013 in Uganda.[38] In the years before 1995, we have assumed that treatment success for HIV-negative individuals is lower and constant, with values for each country dependent on the available data during the period of 1995 to 2022. Where data is available, treatment completion in HIV-positive individuals tracks closely, albeit slightly lower, to treatment completion for HIV-negative individuals. As such, when filling in the years before data was available, we maintained a gap of minimum 4% between HIV-negative and HIV-positive before 1995, and then between 1995 and the first available data point, treatment completion for HIV-positive individuals could not be higher than the lowest value achieved for HIV-negative individuals in the future years. The resulting time trends can be seen in Figure 5.

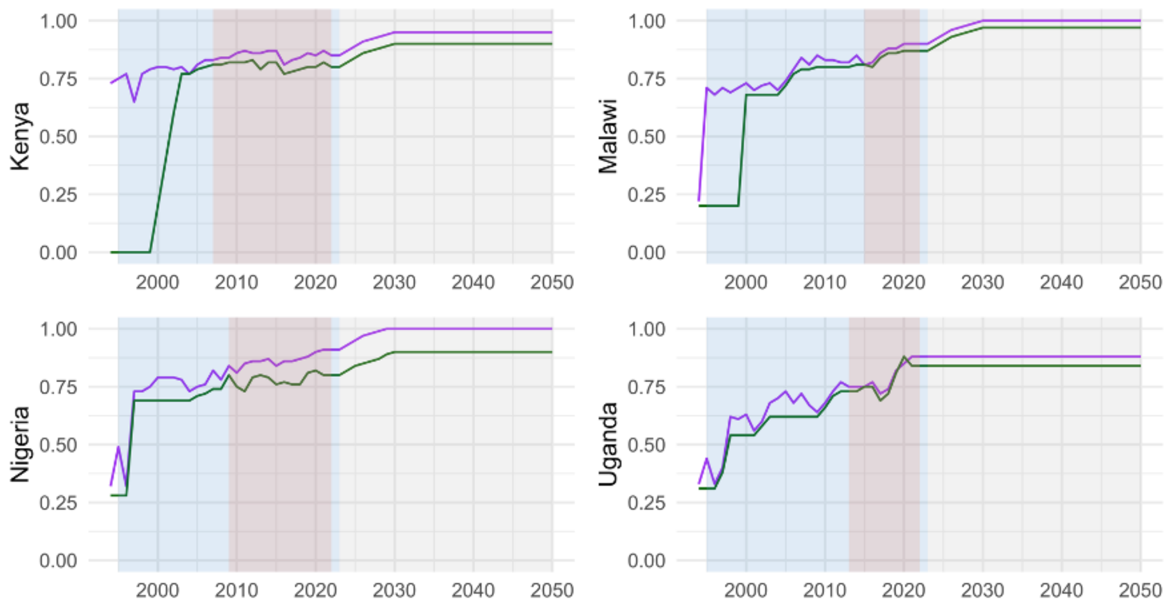

*Figure 5: The treatment completion for each of the four countries over time. The purple line is for HIV-negative treatment completion and the green line is for HIV-positive treatment completion. The blue areas is where data is available for HIV-*

*negative treatment completion, and the red area is where data is available for both HIV-negative and HIV-positive treatment completion*

#### 3.8 Mortality

Disease-specific mortality is only possible from symptomatic disease ($\mu_{\mathbf{sTB}_{a,h,k}}$), with a time-independent parameter that is specific to age, HIV status, and ART duration. The prior range for TB mortality in HIV-negative individuals is shown in Table 16.

The mortality rate for children ages 0-14 is unchanged to that in adults, but there is scope in the model to adjust it:

$$\mu_{\mathbf{sTB}_{a=0-14}} = \mu_{\mathbf{sTB}} RR_{\mu_{\mathbf{sTB}_0}}$$

where $\mu_I$ is the baseline mortality for HIV-negative adults, and $RR_{\mu_{I_0}}$ is the relative risk of symptomatic disease mortality in children. $RR_{\mu_{I_0}}$ is fixed at 1 in this model, as shown in Table 16.

*Table 16: Prior ranges for the parameters for TB mortality*

| Parameter | Description | Prior range or set value | References |
|---|---|---|---|
| $\mu_{\mathbf{sTB}}$ | Annual rate of TB mortality from symptomatic disease | 0.26 - 0.449 | [2, 3, 5, 39] |
| $RR_{\mu_{\mathbf{sTB}_0}}$ | Relative risk of TB mortality from symptomatic disease in children ages 0 - 14 | 1 | [5] |

###### *3.8.1.1 HIV-positive and ART strata*

The mortality rate in HIV-positive individuals can change compared to HIV-negative individuals:

$$\mu_{\mathbf{sTB}_{h=HIV+}} = \mu_{\mathbf{sTB}} RR_{\mu_H}$$

where $\mu_I$ is the baseline mortality for HIV-negative adults, and $RR_{\mu_H}$ is the relative risk of symptomatic disease mortality in HIV-positive individuals. The prior range for $RR_{\mu_H}$ is shown in Table 17.

The mortality rate is adjusted for ART duration in the same way as rapid progression in Section 3.2.1.2, using the values described in Table 17.

References and

*Table 17: Prior ranges and data sources for the risk ratio associated with being HIV-positive and the reduction of risk from ART*

| Parameter | Description | Prior range or set value | References |
|---|---|---|---|
| $RR_{\mu_H}$ | Risk ratio of TB mortality from symptomatic disease in HIV-positive individuals | 1.5 - 2.5 (1 - 3 in Uganda) | [5] |
| $ART_{\mu_{\mathbf{sTB}_{t<6}}}$ | Protective effect of ART treatment against TB/HIV death for treatment duration < 6 months | 0.11 - 0.28 | [1,5,28] |
| $ART_{\mu_{\mathbf{sTB}_{t=6-12}}}$ | Protective effect of ART treatment against TB/HIV death for treatment duration $\geq 12$ months | 0.51 - 0.75 | [1,5,28] |
| $ART_{\mu_{\mathbf{sTB}_{t\geq 12}}}$ | Protective effect of ART treatment against TB/HIV death for treatment duration 12 months | 0.64 - 0.95+ | [1,5,28] |

+$ART_{\mu_{\mathbf{sTB}_{t\geq 12}}}$ must be greater than $ART_{\mu_{\mathbf{sTB}_{t=6-12}}}$

### 4 Model implementation

The model is initialised with the 1970 population, with sex and age structure and 50 symptomatic, treatment-naive TB cases in the age group 30 - 44 years. The model is then run for 200 years with all parameters set at the 1970 values and no HIV.

The resulting population is then re-scaled to the 1970 population and the model is run through to 2050 with time-dependent parameter values and HIV and ART.

The model is implemented in R.[40] Model equations are implemented as ordinary differential equations with a time step of 0.5 years and are solved using the fourth order Runge-Kutte integration method.[41]

### 5 Calibration

#### 5.1 Epidemiological calibration targets

##### 5.1.1 Data sources

Each of these four countries has conducted a prevalence survey, and targets for prevalence are sourced directly from these reports for the respective year of the survey.[42–45] Calculations of the male to female ratios and the prevalence to notification ratios were similarly sourced for each of the respective prevalence survey years.[46] We included incidence and mortality estimates for the overall population from 2010 and 2020 to ensure that the model calibration followed the time trend observed in the countries, along with matching the current disease burden. [35] We also selected a single year for each of these measures for sub-populations: 2020 for incidence and mortality of TB in individuals with HIV, and 2022 for incidence and mortality in men and women (as data is only disaggregated by sex for the most recent year). [35,47–49]

We also included two time points for overall notifications, however to avoid the impact of the COVID-19 pandemic on the reporting of case notifications, we selected 2010 and 2022 for overall notifications, and then used just 2022 for men, women, children, and the HIV-positive notifications.

*Table 18: The country-level targets used to calibrate the models*

| Measure | Population | Year | Target Ranges | | | | Notes | References |
|---|---|---|---|---|---|---|---|---|
| | | | Kenya | Malawi | Nigeria | Uganda | | |
| Prevalence per 100k | Total | K: 2016<br>M: 2014<br>N: 2012<br>U: 2015 | 347-504 | 258-468 | 239-406 | 191-315 | | [42–45] |
| | Men | | 656-962 | 335-757 | 538-965 | 554-914 | | |
| | Women | | 258-460 | 246-501 | 213-505 | 109-248 | | |
| | MtoF | | 1.79-2.83 | 0.94-1.87 | 1.52-2.96 | 2.84-5.81 | | [46] |

| | | | | | | | | |
|---|---|---|---|---|---|---|---|---|
| Prevalence-to-notification ratio | Men | | 3.04-4.46 | 1.65-4.05 | 4.99-9.51 | 2.81-4.64 | | [46] |
| | Women | | 2.5-4.46 | 1.25-3.14 | 2.51-6.74 | 1.24-2.83 | | |
| Incidence per 100k (annual) | Total | 2010 | 174-1080 | 55-870 | 143-311 | 135-301 | | [35] |
| | | 2020 | 155-367 | 73-225 | 143-311 | 120-298 | | |
| | Men | 2022 | 139-769 | 10.6-466.2 | 175.3-682.3 | 102-651.5 | Section 5.1.2.1 | [47,48] |
| | Women | 2022 | 64-346 | 6.5-293.3 | 100.4-390.4 | 61.4-389.5 | | [47,49] |
| | HIV-positive | 2020 | 37-90 | 34-101 | 0.1-0.15 | 39-98 | | [35] |
| Proportion of incidence | Children (%) | 2022 | - | - | 11-24 | - | Only Nigeria | [47] |
| Mortality per 100k (annual) | Total | 2010 | 124-247 | 77-233 | 61-120 | 39-76 | | [35] |
| | | 2020 | 41-82 | 26-57 | 46-105 | 28-55 | | |
| | HIV-positive | 2020 | 13-32 | 13-38 | 6-14 | 13-29 | | |
| Notifications per 100k (annual) | Total | 2010 | 191-298 | 114-172 | 41-87 | 106-159 | Section 5.1.2.2 | [35,50–52] |
| | | 2019 | - | - | - | 123-184 | Section 5.1.2.2 Only Uganda | |
| | Men | 2019 | - | - | - | 254-382 | Section 5.1.2.3 Only Uganda | [48,50–53] |
| | Women | 2019 | - | - | - | 139-208 | Section 5.1.2.3 Only Uganda | [49–53] |
| | Children | 2019 | - | - | - | 33.1-49.7 | Section 5.1.2.3 Only Uganda | [48–50,53] |

| | | | | | | | | |
|---|---|---|---|---|---|---|---|---|
| Proportion of total notifications | HIV-positive (%) | 2019 | - | - | - | 0.313-0.47 | Section 5.1.2.4 Only Uganda | [50,53] |
| Notifications per 100k (annual) | Total | 2022 | 131-205 | 70-106 | 103-215 | 159-240 | Section 5.1.2.2 | [35,50–52] |
| | Men | 2022 | 253-395 | 144-217 | 194-406 | 308-462 | Section 5.1.2.3 | [48,50–53] |
| | Women | 2022 | 120-188 | 84-127 | 142-298 | 181-271 | Section 5.1.2.3 | [49–53] |
| | Children | 2022 | - | - | - | 57.1-85.7 | Section 5.1.2.3 Only Uganda | [48–50,53] |
| Proportion of total notifications | Children (%) | 2022 | 0.096-0.144 | 0.064-0.096 | 0.056-0.084 | - | Section 5.1.2.4 | [50,53] |
| | HIV-positive (%) | 2022 | 0.192-0.288 | 0.384-0.576 | 0.045-0.068 | 26.4-39.6 | | [50,53] |

#### 5.1.2 Calculation notes

##### *5.1.2.1 Incidence in men and women*

The WHO provides the estimated incidence of TB disease for men and women within the *TB incidence disaggregated by age group, sex, and risk factor* file, where this data is provided for the latest year available (2022).[47] These are raw numbers for the lower and upper bounds, and we transformed them into incidence per 100,000 population to use in the model. The UN World Population Prospects provides estimates for the population split by age groups and sex.[48,49] The “middle variant” estimate for future projections matches the total population estimate reported by the WHO, so we have used the “middle variant” estimate for the sex-disaggregated populations.[35]

The calculation for incidence in men and women is then:

$$\frac{\text{Incidence Estimate}}{\text{Population estimate} \times 1000} \times 100{,}000$$

using the data as presented in Table 19

*Table 19*

| Country | Sex | Lower incidence estimates | Upper incidence estimates | Population estimates | Lower estimate/100k | Upper estimate/100k |
|---|---|---|---|---|---|---|
| ***Kenya*** | *Male* | 23,000 | 127,000 | 16,524.716 | 139.2 | 768.5 |
| | *Female* | 11,000 | 59,000 | 17,074.203 | 64.4 | 345.6 |
| ***Malawi*** | *Male* | 590 | 26,000 | 5,577.175 | 10.6 | 466.2 |
| | *Female* | 400 | 18,000 | 6,136.975 | 6.5 | 293.3 |
| ***Nigeria*** | *Male* | 110,000 | 428,000 | 62,732.636 | 175.3 | 682.3 |
| | *Female* | 62,000 | 241,000 | 61,738.365 | 100.4 | 390.4 |
| ***Uganda*** | *Male* | 13,000 | 83,000 | 12,740.267 | 102.0 | 651.5 |
| | *Female* | 8,200 | 52,000 | 13,350.299 | 61.4 | 389.5 |

###### *5.1.2.2 Total notifications*

Notification data is reported as a point value of notifications reported per year, and thus do not take into account under-reporting (for example due to private healthcare diagnoses) or over-reporting (due to misdiagnoses where someone does not have TB), so there may be some uncertainty in the reported numbers.[51,52] The WHO provides notification numbers for each country from 1980.[50] We have used notifications from 2010, but to avoid any impact of COVID-19 on TB notifications, we have used 2022 as the second time-point instead of 2020. The notifications values are transformed to notifications per 100,000 by using the estimated total population for each year, provided by the WHO.[35]

To incorporate uncertainty into our targets, each country has been given an upper and lower multiplier. For countries with no, or limited, under-reporting, this is $\pm 20\%$ using multipliers of 1.2 and 0.8 respectively (Malawi and Uganda). For Kenya and Nigeria, the lower estimate is still $-20\%$ but the upper estimate takes into account the under-reporting. Kenya is reported to have $20\%$ under-reporting, and so to bring this up to 100% we have used an upper-estimate multiplier of 1.25.[51] Nigeria is reported to have 40% under-reporting, which requires an upper-estimate multiplier of 1.67.[52]

The calculation for notification estimates is:

$$\left(\frac{\text{reported notifications}}{\text{estimated population}} \times 100{,}000\right) \times \text{multiplier}$$

*Table 20*

<table>
<tr><th>Country</th><th>Year</th><th>Notifications</th><th>Population</th><th>Lower multiplier</th><th>Upper multiplier</th><th>Point value</th><th>Lower estimate</th><th>Upper estimate</th></tr>
<tr><td rowspan="2">Kenya</td><td>2010</td><td>99,272</td><td>41,517,895</td><td rowspan="2">0.8</td><td rowspan="2">1.25</td><td>239.11</td><td>191.3</td><td>298.9</td></tr>
<tr><td>2022</td><td>88,690</td><td>54,027,487</td><td>164.16</td><td>131.3</td><td>205.2</td></tr>
<tr><td rowspan="2">Malawi</td><td>2010</td><td>21,092</td><td>14,718,422</td><td rowspan="2">0.8</td><td rowspan="2">1.20</td><td>143.30</td><td>114.6</td><td>172.0</td></tr>
<tr><td>2022</td><td>18,025</td><td>20,405,317</td><td>88.33</td><td>70.7</td><td>106.0</td></tr>
<tr><td rowspan="2">Nigeria</td><td>2010</td><td>84,121</td><td>160,952,853</td><td rowspan="2">0.8</td><td rowspan="2">1.67</td><td>52.26</td><td>41.8</td><td>87.3</td></tr>
<tr><td>2022</td><td>282,184</td><td>218,541,212</td><td>129.12</td><td>103.3</td><td>215.6</td></tr>
<tr><td rowspan="3">Uganda</td><td>2010</td><td>42,885</td><td>32,341,728</td><td rowspan="2">0.8</td><td rowspan="3">1.20</td><td>132.60</td><td>106.1</td><td>159.1</td></tr>
<tr><td>2019</td><td>65,897</td><td>42,949,080</td><td>153.43</td><td>122.7</td><td>184.1</td></tr>
<tr><td>2020</td><td>94,457</td><td>47,249,585</td><td>0.8</td><td>199.91</td><td>159.9</td><td>239.9</td></tr>
</table>

###### *5.1.2.3 Notifications in men and women (and children in Uganda)*

The proportion of notifications in men and women are available from the WHO case notifications file and are reported on the WHO TB profile for each country.[50,53]

To calculate the range of notifications per 100,000 for each target, we take the total number of notifications, and the proportion of those notifications that were in men and women aged 15 and older, and multiply each to fund the number of notifications for men and women respectively. This number of notifications is then divided by the respective population size for each sex.[48,49] As with total notifications, the range is calculated using lower and upper multipliers to account for potential over- and under-reporting.[51,52]

The calculation for sex-specific notifications is:

$$\left(\frac{\text{proportion of notifications} \times \text{total reported notifications}}{\text{population of sex}} \times 100000\right) \times \text{multiplier}$$

*Table 21*

| Sex | Country | Year | Proportion | Notifications | Population | Lower multiplier | Upper multiplier | Point value | Lower estimate | Upper estimate |
|---|---|---|---|---|---|---|---|---|---|---|
| Male | ***Kenya*** | 2022 | 0.59 | 88,690 | 16,524,716 | 0.8 | 1.25 | 316.66 | 253.3 | 395.8 |
| | ***Malawi*** | 2022 | 0.56 | 18,025 | 5,577,175 | 0.8 | 1.20 | 180.99 | 144.8 | 217.2 |
| | ***Nigeria*** | 2022 | 0.54 | 282,184 | 62,732,636 | 0.8 | 1.67 | 242.90 | 194.3 | 405.6 |
| | ***Uganda*** | 2019 | 0.54 | 65,897 | 11,265,950 | 0.8 | 1.20 | 315.86 | 252.7 | 379.0 |
| | | 2022 | 0.52 | 94,457 | 12,740,268 | 0.8 | 1.20 | 385.53 | 308.4 | 462.6 |
| Female | ***Kenya*** | 2022 | 0.29 | 88,690 | 17,074,204 | 0.8 | 1.25 | 150.64 | 120.5 | 188.3 |
| | ***Malawi*** | 2022 | 0.36 | 18,025 | 6,136,975 | 0.8 | 1.20 | 105.74 | 84.6 | 126.9 |
| | ***Nigeria*** | 2022 | 0.39 | 282,184 | 61,738,365 | 0.8 | 1.67 | 178.26 | 142.6 | 297.7 |
| | ***Uganda*** | 2019 | 0.31 | 65,897 | 11,840,341 | 0.8 | 1.20 | 172.53 | 138.0 | 207.0 |
| | | 2022 | 0.32 | 94,457 | 13,350,299 | 0.8 | 1.20 | 226.41 | 181.1 | 271.7 |

| | | | | | | | | | | |
|---|---|---|---|---|---|---|---|---|---|---|
| **Children** | ***Uganda*** | 2019 | 0.12 | 65,897 | 19,842,790 | 0.8 | 1.20 | 41.41 | 33.1 | 49.7 |
| | | 2022 | 0.16 | 94,457 | 21,159,019 | 0.8 | 1.20 | 71.43 | 57.1 | 85.7 |

###### *5.1.2.4 Proportion of notifications in children and people living with HIV*

As in Section 5.1.2.3, the proportion of notifications in children and for people living with HIV are available to calculate from the WHO case notifications file and are reported on the WHO TB profile app.[50,53] These measures have been kept as a proportion of all notifications for the targets. As before, error bounds of $\pm 20\%$ have been aded to the point values available, as shown in Table 22.

*Table 22*

| | **Notifications in children** | | | **Notifications in people living with HIV** | | |
|---|---|---|---|---|---|---|
| | **Point value** | **Lower estimate** | **Upper estimate** | **Point value** | **Lower estimate** | **Upper estimate** |
| ***Kenya*** | 12 | 9.6 | 14.4 | 24.0 | 19.2 | 28.8 |
| ***Malawi*** | 8 | 6.4 | 9.6 | 48.0 | 38.4 | 57.6 |
| ***Nigeria*** | 7 | 5.6 | 8.4 | 5.7 | 4.6 | 6.8 |
| ***Uganda*** | | | | 33.0 | 26.4 | 39.6 |

#### 5.2 Calibration process

Each country was calibrated separately using the parameter ranges discussed above in section Section 3 to the unique target ranges for each country from Table 18. We calibrated each model in R using the hmer package (history matching and emulation in R).[40,54,55] For each country, we ran an initial wave of test parameters from two sets of latin hypercube sampling (LHS) across the parameter space, one set for training and one set for validation. Each set used 10 times the number of parameters for each country, giving a total of 640 parameter sets for Kenya, Malawi, and Nigeria, and 700 parameter sets for Uganda across both the training and validation sets. Using the output from the training parameter sets, we created emulators for the model. It was possible to check the accuracy of the model by comparing the output of the validation sets against the emulators from the training sets.

As outlined in Section 3, there are some parameter constraints, specified below in Equation 1.

$$\begin{cases} \alpha_{a\geq15} > \alpha_{a=0-14} & \text{rate of progression to disease is higher in children} \\ r > r_H & \text{rate of self-cure is higher in HIV-negative individuals} \\ eps > eps_H & \text{rate of disease regression is higher in HIV-negative individuals} \\ ART_{TB_{t\geq12}} > ART_{TB_{t=6-12}} & \text{reduction in disease risk greater for longer ART durations} \\ RR_{\mu_{\mathbf{sTB}_{t\geq12}}} > RR_{\mu_{\mathbf{sTB}_{t=6-12}}} & \text{reduction in mortality risk is greater for longer ART durations} \\ p_{g=F} > p_{g=M} & \text{treatment coverage for women is higher than for men} \\ \square\square_{g=M} > p_{d=\mathbf{aTB}} & \text{treatment coverage for men is higher than for asymptomatic disease} \\ p_{k=ART} > p_{g=F} & \text{treatment coverage for people on ART is higher than for women} \end{cases} \quad (1)$$

Non-implausible points were generated based on the emulators created from the training data and were checked against these parameter constraints. Again, we were looking for 20 times the number of parameters to feed into the next wave of the process. For the next, and each successive waves, those non-implausible points were run through the model and then randomly sampled to split in half for the training and validation data.

We ran this process for 21 waves. After a set number of waves (5 for Kenya and Malawi, 10 for Nigeria and Uganda), we set a further argument in the non-implausible points selection of `nth = 1` which requires that the non-implausible points selection only aims for parameter sets that hit all of the targets rather than allowing for one or two of the targets to be missed.

Once we had run all 21 waves, we collected all the parameter sets from every wave that had hit all the targets. We ran a de-duplication check and then randomly sampled 1000 parameter sets that we have used for the remainder of the analysis. The distribution of the chosen parameter sets against the priors for each country are shown in Section 5.2.1

##### 5.2.1 Successful calibrations

For each of the countries we present the wave output of the hmer calibration, showing the progress of each country's calibration over successive waves. We then present a plot comparing the priors and posteriors for each parameter and a table comparing the numerical values with the median and 95%UIs for the posterior parameters. Finally, we show the time series plots for each of the calibration targets.

###### *5.2.1.1 Kenya*

Over nine waves, the calibration for Kenya plateaued at about 340/640 of the tested parameter sets hitting all 20 targets (Figure 6). We used the emulators from wave 9 to find suitable parameter sets.

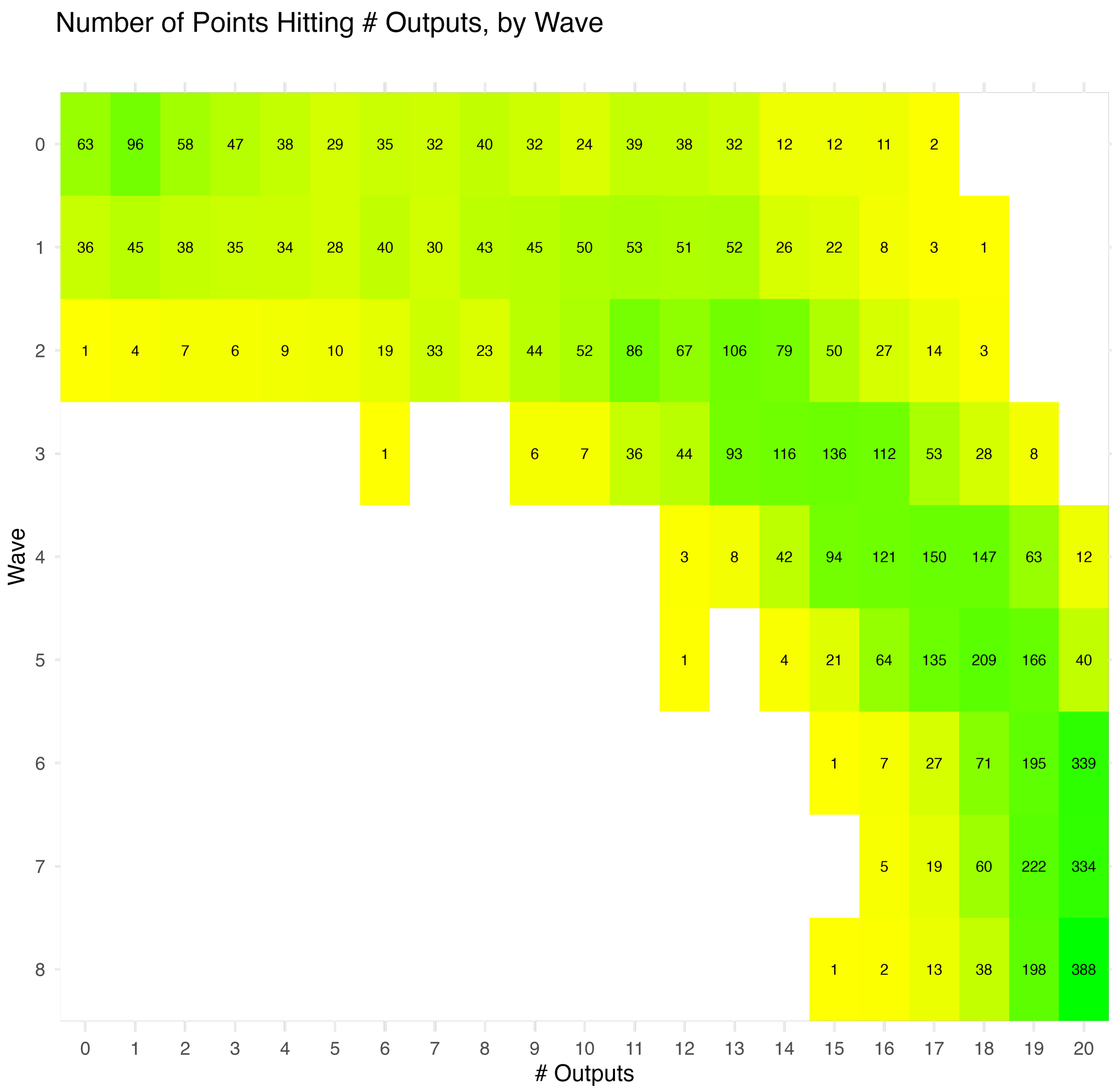

*Figure 6*

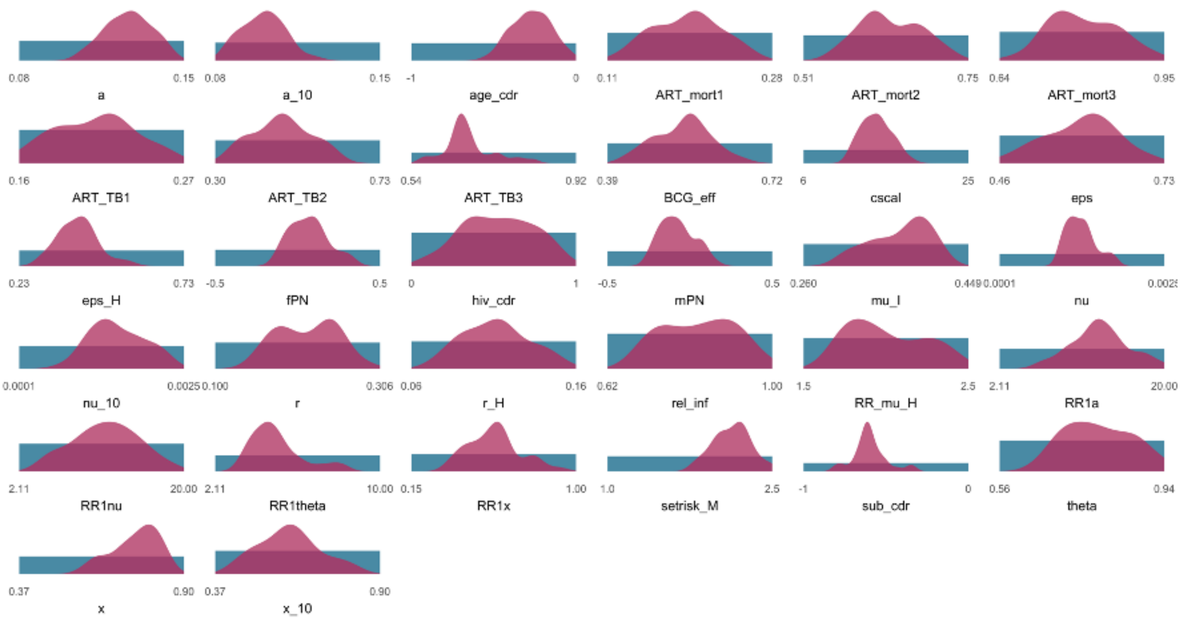

*Figure 7: The prior ranges and posterior distributions for each parameter for the 1000 selected parameter sets for Kenya*

*Table 23: The prior ranges and posterior central estimates and 95% uncertainty intervals from the 1000 calibrated parameter sets for Kenya*

| Parameters | Symbols | Description | Priors | Posteriors |
|---|---|---|---|---|
| a | $\alpha_{a\geq15}$ | Progression to disease, adults | 0.08-0.15 | 0.13 [UI 0.11-0.14] |
| a_10 | $\alpha_{a=0-14}$ | Progression to disease, children | 0.08-0.15 | 0.10 [UI 0.09-0.12] |
| age_cdr | $p_{a=0-14}$ | Treatment coverage adjustment, children | -1.00-0.00 | -0.28 [UI -0.46--0.07] |
| ART_mort1 | $ART_{\mu_{\mathbf{sTB}_{t<6}}}$ | ART mortality adjustment, 0-6 months | 0.11-0.28 | 0.20 [UI 0.14-0.25] |
| ART_mort2 | $ART_{\mu_{\mathbf{sTB}_{t=6-12}}}$ | ART mortality adjustment, 6-12 months | 0.51-0.75 | 0.62 [UI 0.56-0.72] |
| ART_mort3 | $ART_{\mu_{\mathbf{sTB}_{t\geq12}}}$ | ART mortality adjustment, 12+ months | 0.64-0.95 | 0.79 [UI 0.71-0.92] |
| ART_TB1 | $ART_{TB_{t<6}}$ | ART risk reduction, 0-6 months | 0.16-0.27 | 0.22 [UI 0.16-0.26] |
| ART_TB2 | $ART_{TB_{t=6-12}}$ | ART risk reduction, 6-12 months | 0.30-0.73 | 0.47 [UI 0.35-0.60] |
| ART_TB3 | $ART_{TB_{t\geq12}}$ | ART risk reduction, 12+ months | 0.54-0.92 | 0.66 [UI 0.58-0.80] |

| | | | | |
|---|---|---|---|---|
| BCG_eff | $BCG_{eff_{a\geq15}}$ | BCF efficacy | 0.39-0.72 | 0.55 [UI 0.44-0.64] |
| cscal | $cscal$ | Contact scaling parameter | 6.00-25.00 | 14.34 [UI 11.63-17.49] |
| eps | $\epsilon_{h=HIV-}$ | Regression through disease, HIV- | 0.46-0.73 | 0.60 [UI 0.49-0.70] |
| eps_H | $\epsilon_{h=HIV+}$ | Regression through disease, HIV+ | 0.23-0.73 | 0.41 [UI 0.31-0.54] |
| fPN | $p_{g=F}$ | Treatment coverage adjustment, women | -0.50-0.50 | 0.07 [UI -0.11-0.31] |
| hiv_cdr | $p_{k=ART}$ | Treatment coverage adjustment, ART | 0.00-1.00 | 0.52 [UI 0.13-0.87] |
| mPN | $p_{g=M}$ | Treatment coverage adjustment, men | -0.50-0.50 | -0.08 [UI -0.23-0.10] |
| mu_I | $\mu_{\mathbf{sTB}}$ | TB mortality from sympomatic disease | 0.26-0.45 | 0.38 [UI 0.31-0.41] |
| nu | $\nu_{a\geq15}$ | Progression from infection, adults | 0.00-0.00 | 0.00 [UI 0.00-0.00] |
| nu_10 | $\nu_{a=0-14}$ | Progression from infection, children | 0.00-0.00 | 0.00 [UI 0.00-0.00] |
| r | $r_{h=HIV-}$ | Self cure, HIV- | 0.10-0.31 | 0.22 [UI 0.16-0.28] |
| r_H | $r_{h=HIV+}$ | Self cure, HIV+ | 0.06-0.16 | 0.11 [UI 0.08-0.15] |
| rel_inf | $q$ | Relative infectiousness, asymptomatic disease | 0.62-1.00 | 0.83 [UI 0.70-0.97] |
| RR_mu_H | $RR_{\mu_H}$ | Relative risk of mortality, HIV+ | 1.50-2.50 | 1.95 [UI 1.71-2.41] |
| RR1a | $RR_{1_\alpha}$ | Relative risk of progression to disease, HIV+ | 2.11-20.00 | 12.89 [UI 7.29-18.70] |
| RR1nu | $RR_{1_\nu}$ | Relative risk of progression from infection, HIV+ | 2.11-20.00 | 11.64 [UI 5.40-17.79] |
| RR1theta | $RR_{1_\theta}$ | Relative risk of progression through disease, HIV+ | 2.11-10.00 | 4.73 [UI 3.46-8.03] |
| RR1x | $RR_{1_x}$ | Relative risk of reinfection, HIV+ | 0.15-1.00 | 0.58 [UI 0.37-0.82] |

| | | | | |
|---|---|---|---|---|
| setrisk_M | $RR_{sex_{g=M,a\geq15}}$ | Excess risk for men | 1.00-2.50 | 2.14 [UI 1.81-2.46] |
| sub_cdr | $p_{d=\mathbf{aTB}}$ | Treatment coverage adjustment, asymptomatic disease | -1.00-0.00 | -0.61 [UI -0.79--0.36] |
| theta | $\theta$ | Progression through disease | 0.56-0.94 | 0.78 [UI 0.67-0.93] |
| x | $x_{a\geq15}$ | Reinfection, adults | 0.37-0.90 | 0.77 [UI 0.61-0.84] |
| x_10 | $x_{a=0-14}$ | Reinfection, chilldren | 0.37-0.90 | 0.60 [UI 0.44-0.80] |

All selected parameter sets have model outputs that fit within the target ranges (Figure 8). The prevalence estimates lie on the higher end of the calibration targets, whilst incidence and mortality lie on the lower ends. P:N ratios are relatively stable over time whilst the M:F prevalence ratio increases slightly over time.

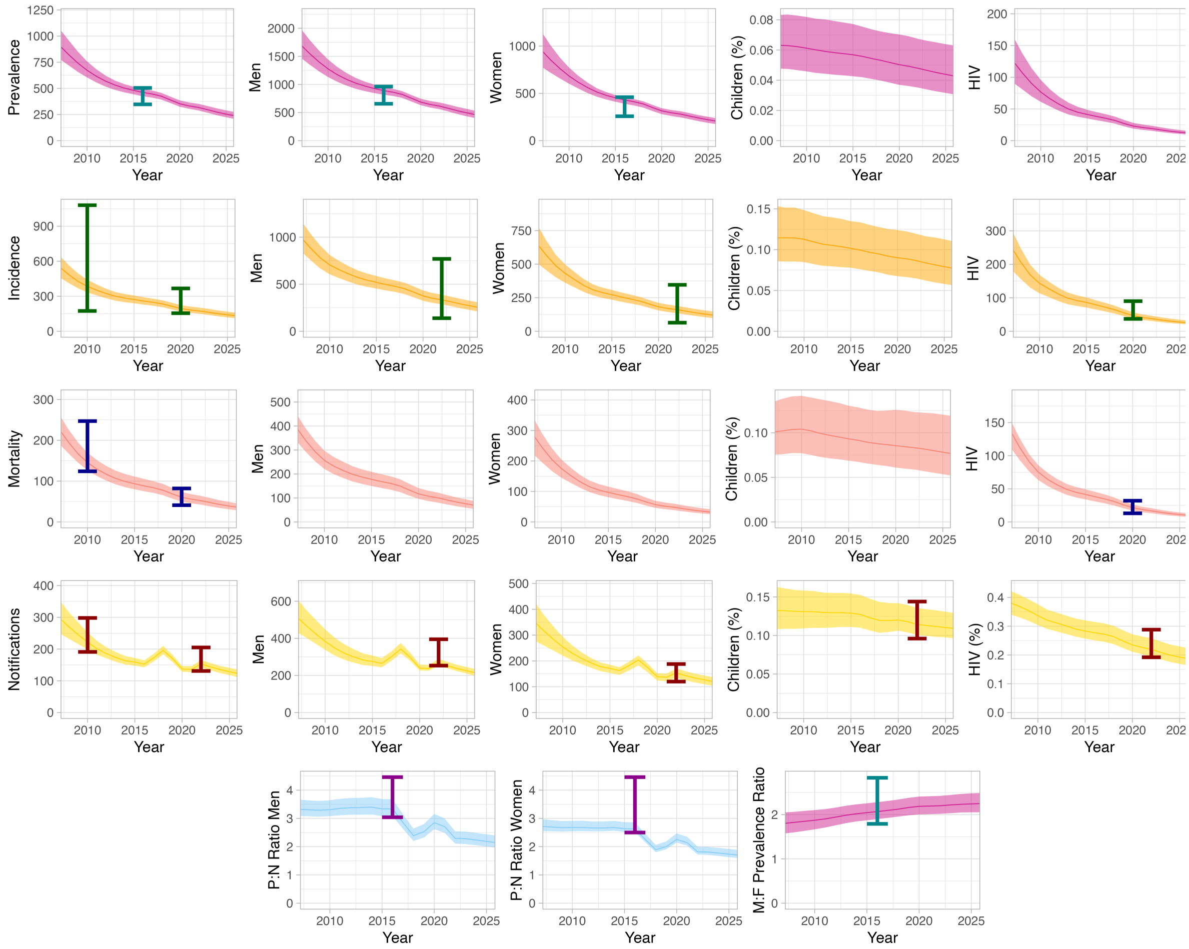

*Figure 8: Calibration plots for the Kenyan calibration targets using the 1000 selected parameters. The targets are the coloured error bars, the lines are the median estimates and the shaded regions are the 95% uncertainty intervals*

##### 5.2.1.2 Malawi

Over nine waves, the calibration for Malawi plateaued at about 350/640 of the tested parameter sets hitting all 20 targets (Figure 9). We used the emulators from wave 9 to find suitable parameter sets.

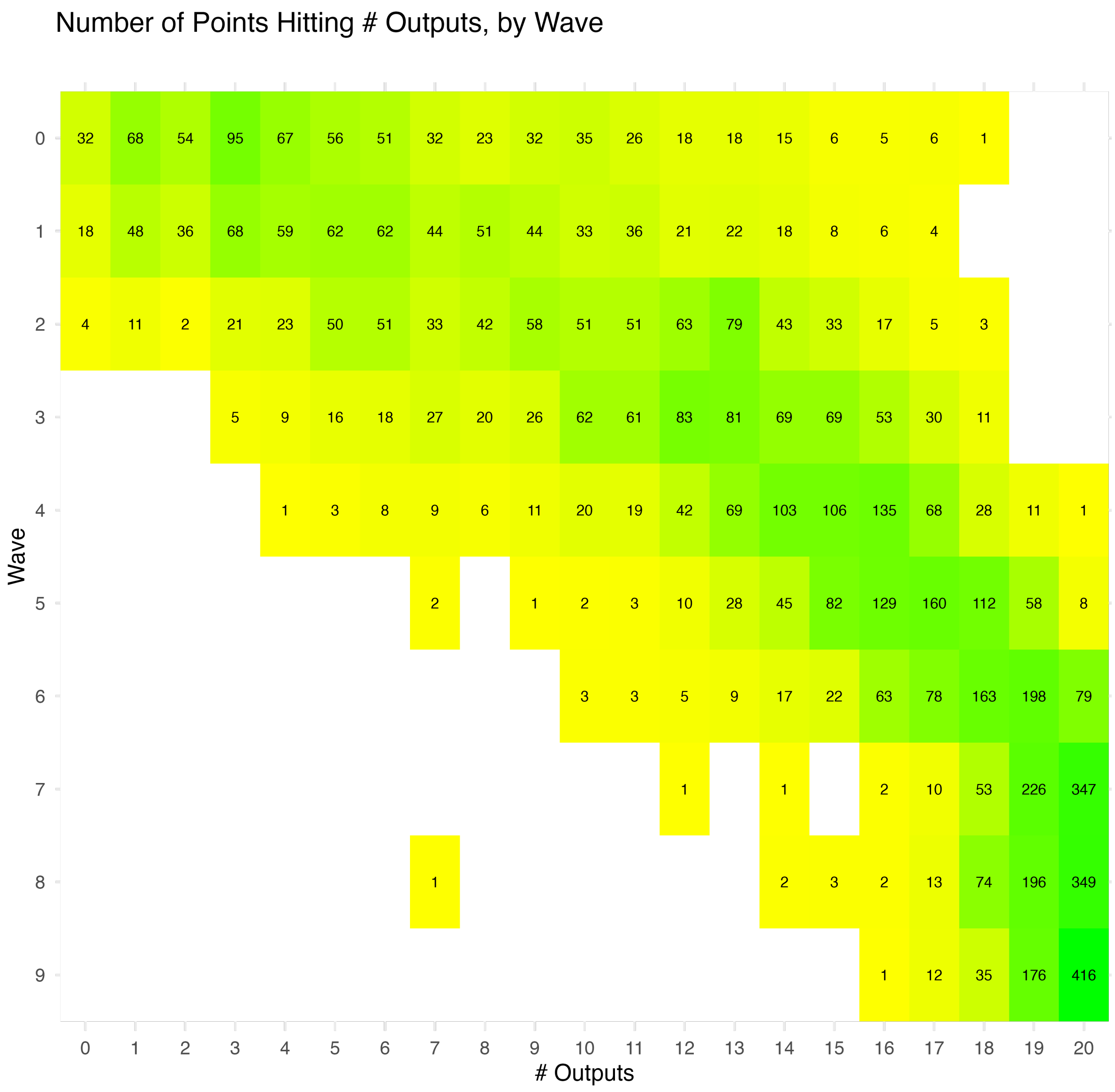

*Figure 9*

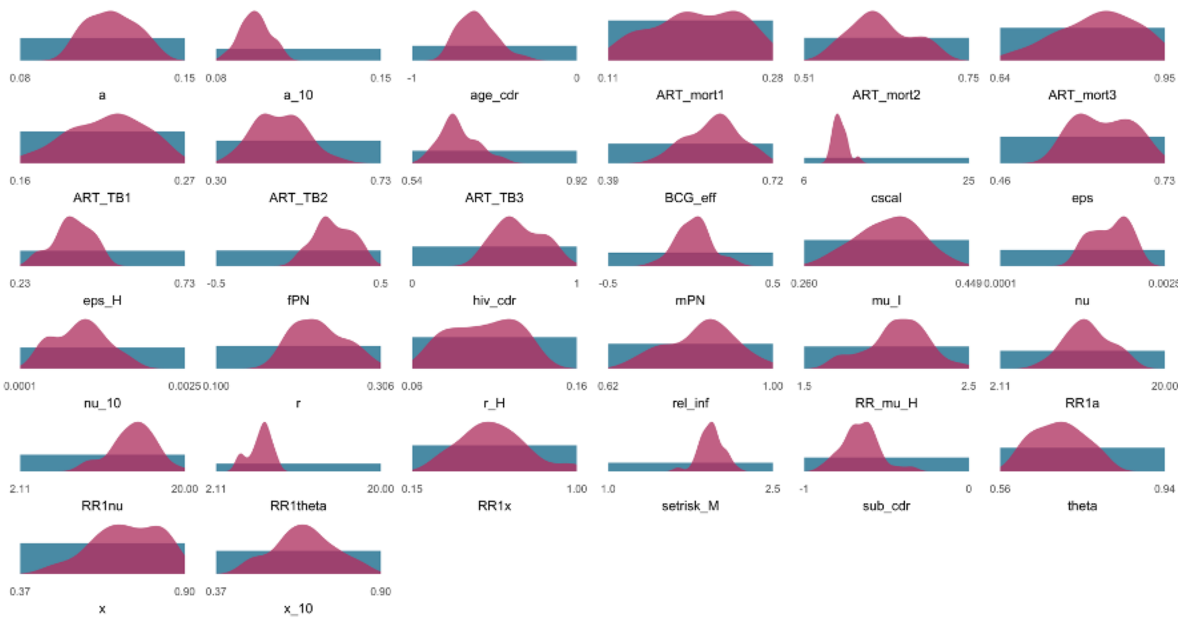

*Figure 10: The prior ranges and posterior distributions for each parameter for the 1000 selected parameter sets for Malawi*

*Table 24: The prior ranges and posterior central estimates and 95% uncertainty intervals from the 1000 calibrated parameter sets for Malawi*

| Parameters | Symbols | Description | Priors | Posteriors |
|---|---|---|---|---|
| a | $\alpha_{a\geq15}$ | Progression to disease, adults | 0.08-0.15 | 0.12 [UI 0.10-0.14] |
| a_10 | $\alpha_{a=0-14}$ | Progression to disease, children | 0.08-0.15 | 0.10 [UI 0.09-0.11] |
| age_cdr | $p_{a=0-14}$ | Treatment coverage adjustment, children | -1.00-0.00 | -0.62 [UI -0.77--0.37] |
| ART_mort1 | $ART_{\mu_{\mathbf{sTB}_{t<6}}}$ | ART mortality adjustment, 0-6 months | 0.11-0.28 | 0.20 [UI 0.13-0.27] |
| ART_mort2 | $ART_{\mu_{\mathbf{sTB}_{t=6-12}}}$ | ART mortality adjustment, 6-12 months | 0.51-0.75 | 0.61 [UI 0.55-0.70] |
| ART_mort3 | $ART_{\mu_{\mathbf{sTB}_{t\geq12}}}$ | ART mortality adjustment, 12+ months | 0.64-0.95 | 0.83 [UI 0.67-0.93] |
| ART_TB1 | $ART_{TB_{t<6}}$ | ART risk reduction, 0-6 months | 0.16-0.27 | 0.22 [UI 0.17-0.25] |
| ART_TB2 | $ART_{TB_{t=6-12}}$ | ART risk reduction, 6-12 months | 0.30-0.73 | 0.47 [UI 0.35-0.59] |

| | | | | |
|---|---|---|---|---|
| ART_TB3 | $ART_{TB_{t\geq12}}$ | ART risk reduction, 12+ months | 0.54-0.92 | 0.64 [UI 0.58-0.74] |
| BCG_eff | $BCG_{eff_{a\geq15}}$ | BCF efficacy | 0.39-0.72 | 0.61 [UI 0.51-0.69] |
| cscal | $cscal$ | Contact scaling parameter | 6.00-25.00 | 10.00 [UI 9.00-12.16] |
| eps | $\epsilon_{h=HIV-}$ | Regression through disease, HIV- | 0.46-0.73 | 0.63 [UI 0.56-0.70] |
| eps_H | $\epsilon_{h=HIV+}$ | Regression through disease, HIV+ | 0.23-0.73 | 0.39 [UI 0.27-0.48] |
| fPN | $p_{g=F}$ | Treatment coverage adjustment, women | -0.50-0.50 | 0.19 [UI 0.02-0.40] |
| hiv_cdr | $p_{k=ART}$ | Treatment coverage adjustment, ART | 0.00-1.00 | 0.62 [UI 0.43-0.88] |
| mPN | $p_{g=M}$ | Treatment coverage adjustment, men | -0.50-0.50 | 0.03 [UI -0.16-0.23] |
| mu_I | $\mu_{\mathbf{sTB}}$ | TB mortality from sympomatic disease | 0.26-0.45 | 0.36 [UI 0.30-0.42] |
| nu | $\nu_{a\geq15}$ | Progression from infection, adults | 0.00-0.00 | 0.00 [UI 0.00-0.00] |
| nu_10 | $\nu_{a=0-14}$ | Progression from infection, children | 0.00-0.00 | 0.00 [UI 0.00-0.00] |
| r | $r_{h=HIV-}$ | Self cure, HIV- | 0.10-0.31 | 0.23 [UI 0.18-0.29] |
| r_H | $r_{h=HIV+}$ | Self cure, HIV+ | 0.06-0.16 | 0.11 [UI 0.07-0.13] |
| rel_inf | $q$ | Relative infectiousness, asymptomatic disease | 0.62-1.00 | 0.84 [UI 0.68-0.96] |
| RR_mu_H | $RR_{\mu_H}$ | Relative risk of mortality, HIV+ | 1.50-2.50 | 2.09 [UI 1.70-2.37] |
| RR1a | $RR_{1_\alpha}$ | Relative risk of progression to disease, HIV+ | 2.11-20.00 | 11.35 [UI 7.31-16.06] |
| RR1nu | $RR_{1_\nu}$ | Relative risk of progression from infection, HIV+ | 2.11-20.00 | 14.50 [UI 9.50-18.12] |
| RR1theta | $RR_{1_\theta}$ | Relative risk of progression through disease, HIV+ | 2.11-20.00 | 7.21 [UI 4.55-8.53] |

| | | | | |
|---|---|---|---|---|
| RR1x | $RR_{1_x}$ | Relative risk of reinfection, HIV+ | 0.15-1.00 | 0.56 [UI 0.30-0.96] |
| setrisk_M | $RR_{sex_{g=M,a\geq15}}$ | Excess risk for men | 1.00-2.50 | 1.94 [UI 1.75-2.11] |
| sub_cdr | $p_{d=\mathbf{aTB}}$ | Treatment coverage adjustment, asymptomatic disease | -1.00-0.00 | -0.65 [UI -0.85--0.42] |
| theta | $\theta$ | Progression through disease | 0.56-0.94 | 0.70 [UI 0.60-0.80] |
| x | $x_{a\geq15}$ | Reinfection, adults | 0.37-0.90 | 0.72 [UI 0.49-0.86] |
| x_10 | $x_{a=0-14}$ | Reinfection, chilldren | 0.37-0.90 | 0.66 [UI 0.48-0.83] |

All selected parameter sets have model outputs that fit within the target ranges (Figure 11). The prevalence estimates lie in the centre of the calibration targets, whilst incidence and mortality lie on the lower ends. P:N ratios decrease over time whilst the M:F prevalence ratio is relatively stable.

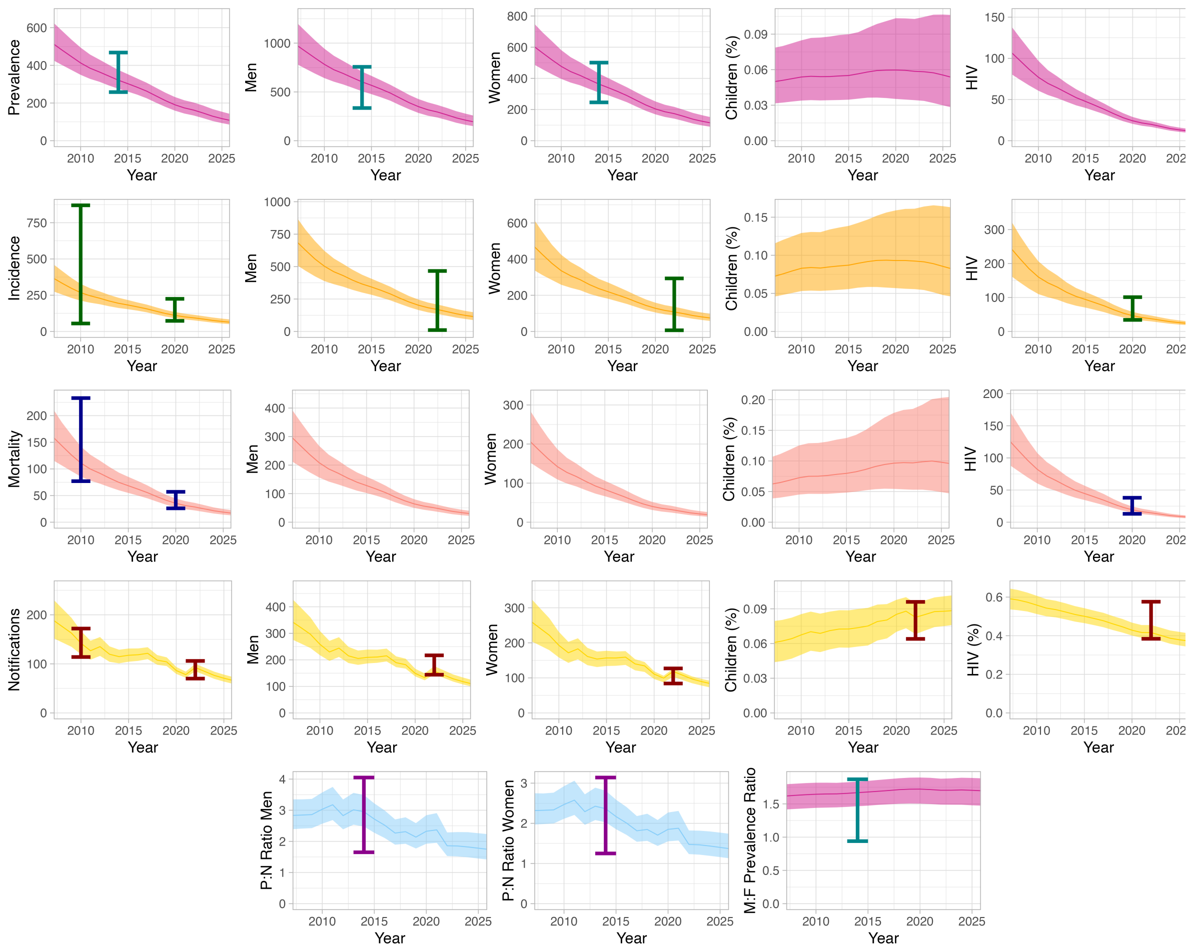

*Figure 11: Calibration plots for the Malawian calibration targets using the 1000 selected parameters. The targets are the coloured error bars, the lines are the median estimates and the shaded regions are the 95% uncertainty intervals*

##### 5.2.1.3 Nigeria

Over ten waves, the calibration for Kenya plateaued with between 30 and 80/640 of the tested parameter sets hitting all 21 targets (Figure 12). These were no longer improving and so we used the emulators from wave 9 to find suitable parameter sets.

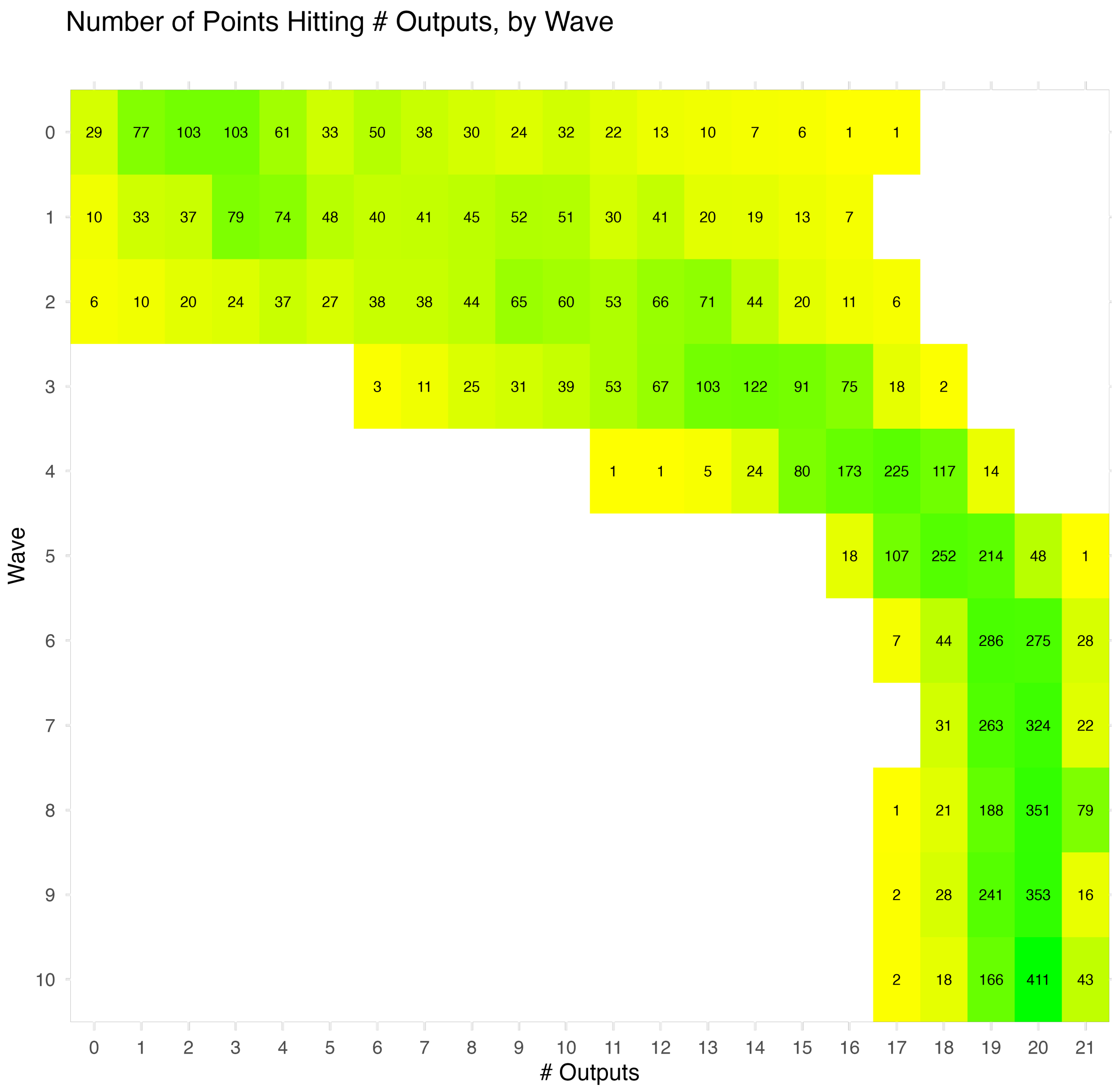

*Figure 12*

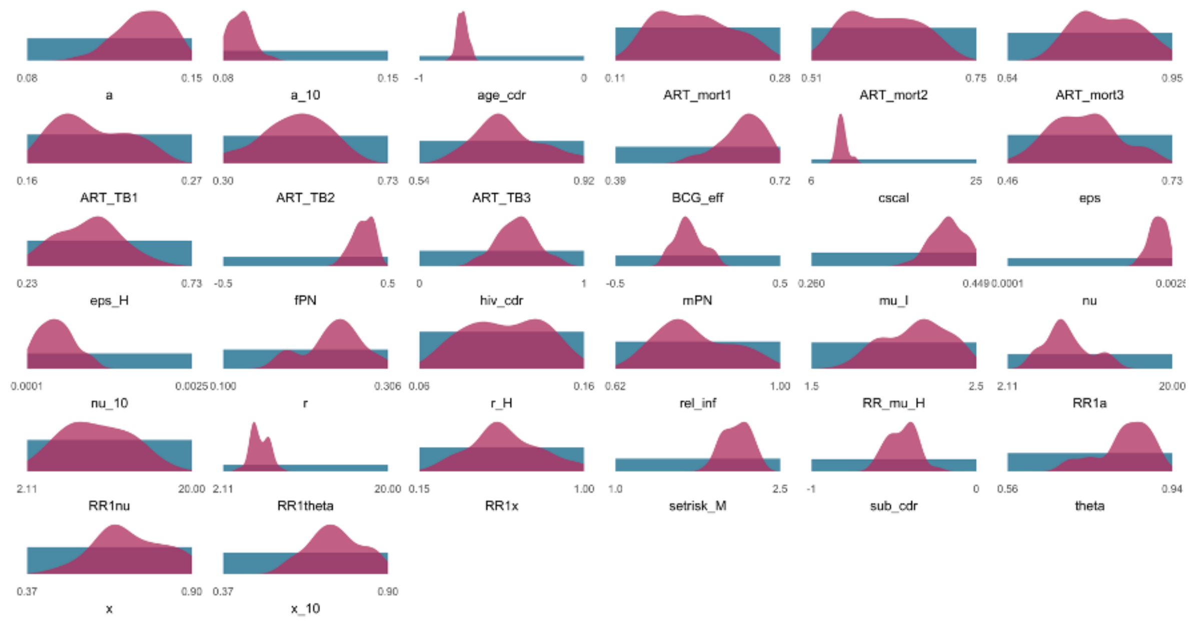

*Figure 13: The prior ranges and posterior distributions for each parameter for the 1000 selected parameter sets for Nigeria*

*Table 25: The prior ranges and posterior central estimates and 95% uncertainty intervals from the 1000 calibrated parameter sets for Nigeria*

| Parameters | Symbols | Description | Priors | Posteriors |
|---|---|---|---|---|
| a | $\alpha_{a\geq15}$ | Progression to disease, adults | 0.08-0.15 | 0.13 [UI 0.11-0.15] |
| a_10 | $\alpha_{a=0-14}$ | Progression to disease, children | 0.08-0.15 | 0.09 [UI 0.08-0.10] |
| age_cdr | $p_{a=0-14}$ | Treatment coverage adjustment, children | -1.00-0.00 | -0.74 [UI -0.78--0.68] |
| ART_mort1 | $ART_{\mu_{\mathbf{sTB}_{t<6}}}$ | ART mortality adjustment, 0-6 months | 0.11-0.28 | 0.19 [UI 0.14-0.25] |
| ART_mort2 | $ART_{\mu_{\mathbf{sTB}_{t=6-12}}}$ | ART mortality adjustment, 6-12 months | 0.51-0.75 | 0.61 [UI 0.54-0.70] |
| ART_mort3 | $ART_{\mu_{\mathbf{sTB}_{t\geq12}}}$ | ART mortality adjustment, 12+ months | 0.64-0.95 | 0.82 [UI 0.74-0.94] |
| ART_TB1 | $ART_{TB_{t<6}}$ | ART risk reduction, 0-6 months | 0.16-0.27 | 0.20 [UI 0.17-0.24] |
| ART_TB2 | $ART_{TB_{t=6-12}}$ | ART risk reduction, 6-12 months | 0.30-0.73 | 0.50 [UI 0.31-0.62] |

| | | | | |
|---|---|---|---|---|
| ART_TB3 | $ART_{TB_{t\geq12}}$ | ART risk reduction, 12+ months | 0.54-0.92 | 0.73 [UI 0.63-0.91] |
| BCG_eff | $BCG_{eff_{a\geq15}}$ | BCF efficacy | 0.39-0.72 | 0.65 [UI 0.55-0.71] |
| cscal | $cscal$ | Contact scaling parameter | 6.00-25.00 | 9.46 [UI 8.71-10.93] |
| eps | $\epsilon_{h=HIV-}$ | Regression through disease, HIV- | 0.46-0.73 | 0.59 [UI 0.50-0.69] |
| eps_H | $\epsilon_{h=HIV+}$ | Regression through disease, HIV+ | 0.23-0.73 | 0.43 [UI 0.25-0.58] |
| fPN | $p_{g=F}$ | Treatment coverage adjustment, women | -0.50-0.50 | 0.35 [UI 0.23-0.44] |
| hiv_cdr | $p_{k=ART}$ | Treatment coverage adjustment, ART | 0.00-1.00 | 0.60 [UI 0.36-0.83] |
| mPN | $p_{g=M}$ | Treatment coverage adjustment, men | -0.50-0.50 | -0.07 [UI -0.19-0.11] |
| mu_I | $\mu_{\mathbf{sTB}}$ | TB mortality from sympomatic disease | 0.26-0.45 | 0.42 [UI 0.38-0.45] |
| nu | $\nu_{a\geq15}$ | Progression from infection, adults | 0.00-0.00 | 0.00 [UI 0.00-0.00] |
| nu_10 | $\nu_{a=0-14}$ | Progression from infection, children | 0.00-0.00 | 0.00 [UI 0.00-0.00] |
| r | $r_{h=HIV-}$ | Self cure, HIV- | 0.10-0.31 | 0.24 [UI 0.17-0.30] |
| r_H | $r_{h=HIV+}$ | Self cure, HIV+ | 0.06-0.16 | 0.11 [UI 0.07-0.15] |
| rel_inf | $q$ | Relative infectiousness, asymptomatic disease | 0.62-1.00 | 0.78 [UI 0.69-0.96] |
| RR_mu_H | $RR_{\mu_H}$ | Relative risk of mortality, HIV+ | 1.50-2.50 | 2.16 [UI 1.82-2.41] |
| RR1a | $RR_{1_\alpha}$ | Relative risk of progression to disease, HIV+ | 2.11-20.00 | 8.17 [UI 5.05-13.27] |
| RR1nu | $RR_{1_\nu}$ | Relative risk of progression from infection, HIV+ | 2.11-20.00 | 9.48 [UI 3.40-15.77] |
| RR1theta | $RR_{1_\theta}$ | Relative risk of progression through disease, HIV+ | 2.11-20.00 | 5.81 [UI 4.63-7.55] |

| | | | | |
|---|---|---|---|---|
| RR1x | $RR_{1_x}$ | Relative risk of reinfection, HIV+ | 0.15-1.00 | 0.57 [UI 0.31-0.96] |
| setrisk_M | $RR_{sex_{g=M,a\geq15}}$ | Excess risk for men | 1.00-2.50 | 2.12 [UI 1.91-2.31] |
| sub_cdr | $p_{d=\mathbf{aTB}}$ | Treatment coverage adjustment, asymptomatic disease | -1.00-0.00 | -0.44 [UI -0.60--0.31] |
| theta | $\theta$ | Progression through disease | 0.56-0.94 | 0.84 [UI 0.70-0.91] |
| x | $x_{a\geq15}$ | Reinfection, adults | 0.37-0.90 | 0.67 [UI 0.51-0.89] |
| x_10 | $x_{a=0-14}$ | Reinfection, chilldren | 0.37-0.90 | 0.73 [UI 0.58-0.87] |

All selected parameter sets have model outputs that fit within the target ranges (Figure 14). Overall prevalence estimates lie on the higher end of the calibration targets, whilst prevalence estimates for men and women are more central, along with incidence and mortality estimates. P:N ratios are relatively stable over time until 2020 when active case finding strategies were implemented, at which point they reduce.[56] At the point P:N ratios decrease, the M:F prevalence ratio starts increasing slightly.

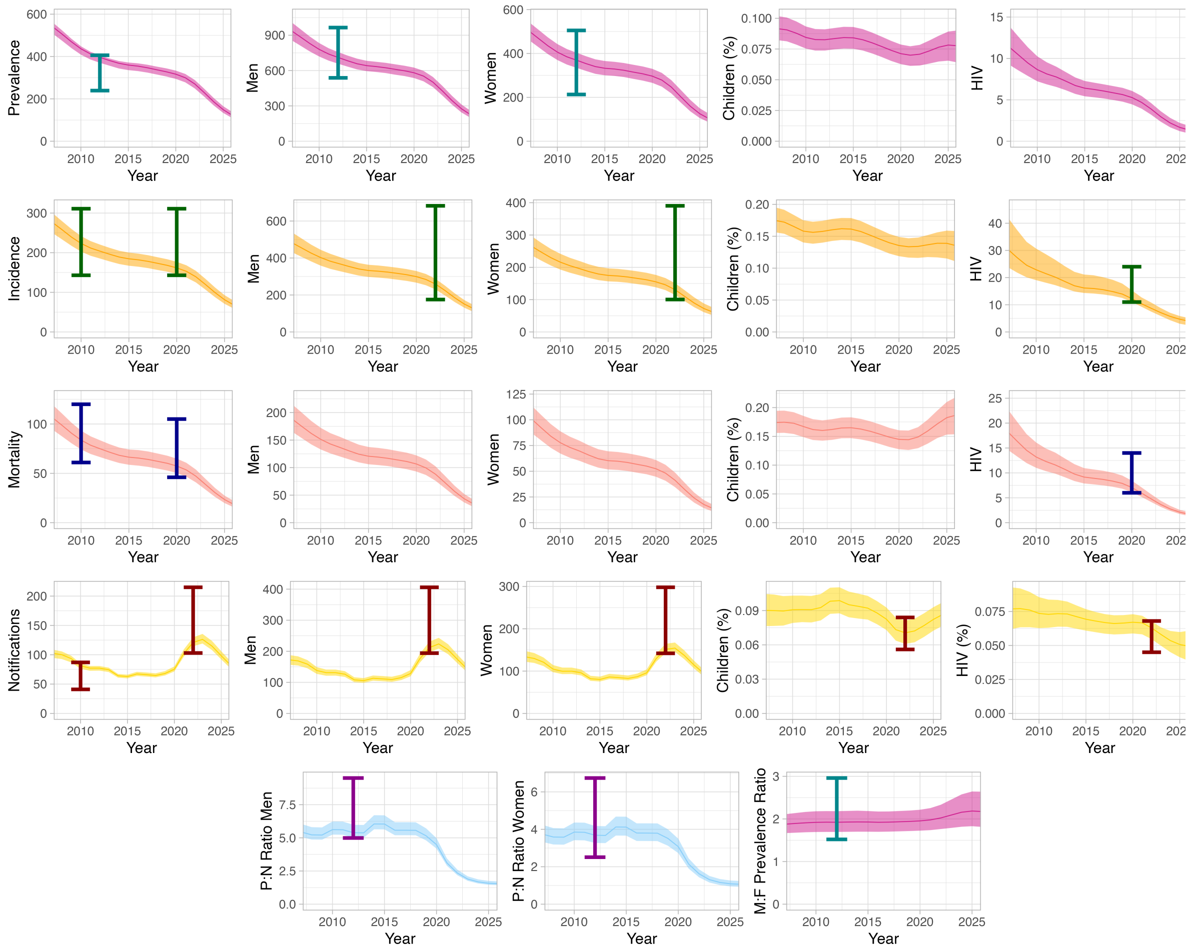

*Figure 14: Calibration plots for the Nigerian calibration targets using the 1000 selected parameters. The targets are the coloured error bars, the lines are the median estimates and the shaded regions are the 95% uncertainty intervals*

###### 5.2.1.4 Uganda

Over ten waves, the calibration for Uganda reached about 125/700 of the tested parameter sets hitting all 25 targets (Figure 15). We used the emulators from wave 10 to find suitable parameter sets.

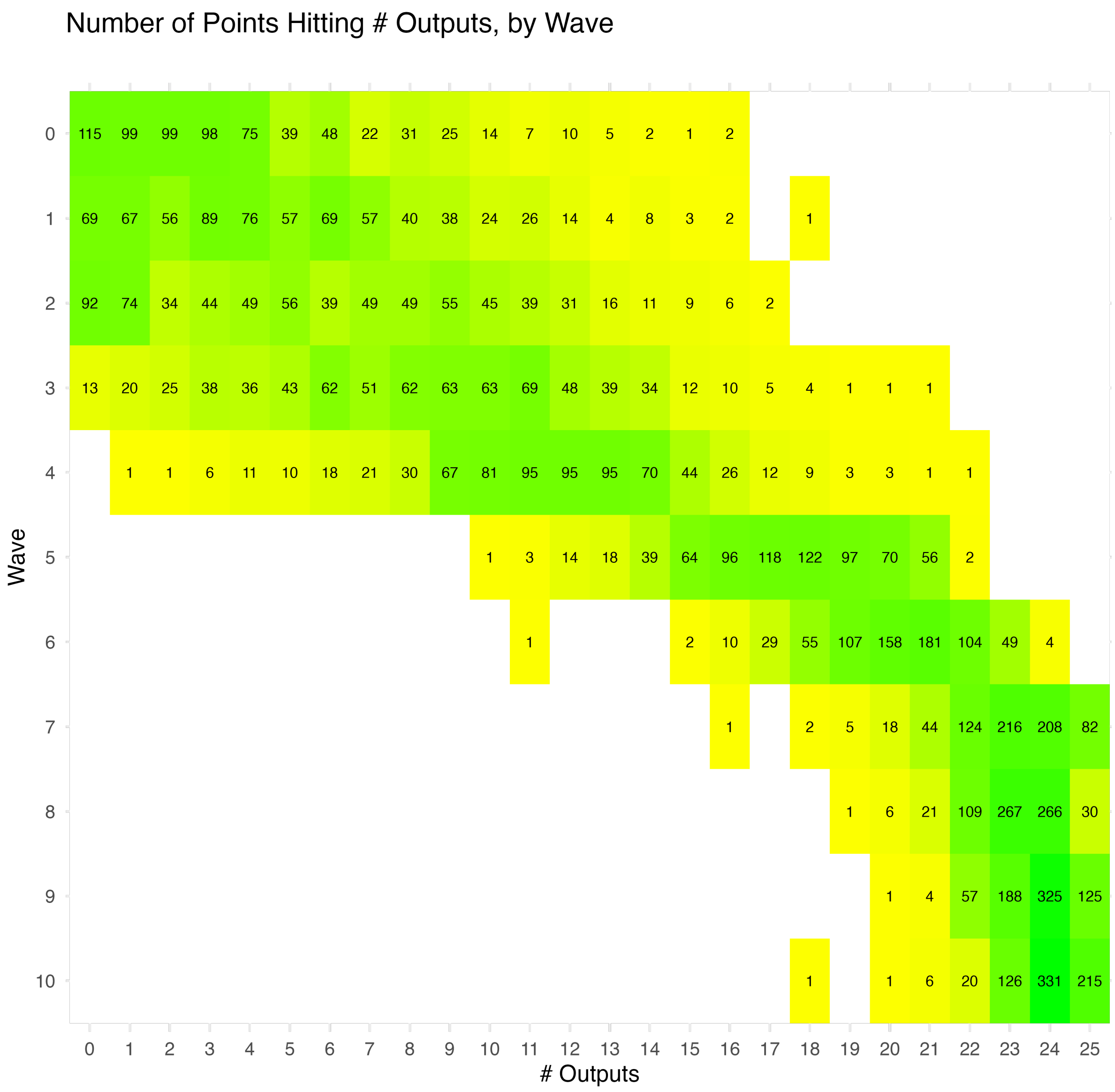
Number of Points Hitting # Outputs, by Wave
Wave
### Outputs
0 1 2 3 4 5 6 7 8 9 10 11 12 13 14 15 16 17 18 19 20 21 22 23 24 25
0 1 2 3 4 5 6 7 8 9 10
115 99 99 98 75 39 48 22 31 25 14 7 10 5 2 1 2
69 67 56 89 76 57 69 57 40 38 24 26 14 4 8 3 2 1
92 74 34 44 49 56 39 49 49 55 45 39 31 16 11 9 6 2
13 20 25 38 36 43 62 51 62 63 63 69 48 39 34 12 10 5 4 1 1 1
1 1 6 11 10 18 21 30 67 81 95 95 95 70 44 26 12 9 3 3 1 1
1 3 14 18 39 64 96 118 122 97 70 56 2
1 2 10 29 55 107 158 181 104 49 4
1 2 5 18 44 124 216 208 82
1 6 21 109 267 266 30
1 4 57 188 325 125
1 1 6 20 126 331 215

*Figure 15*

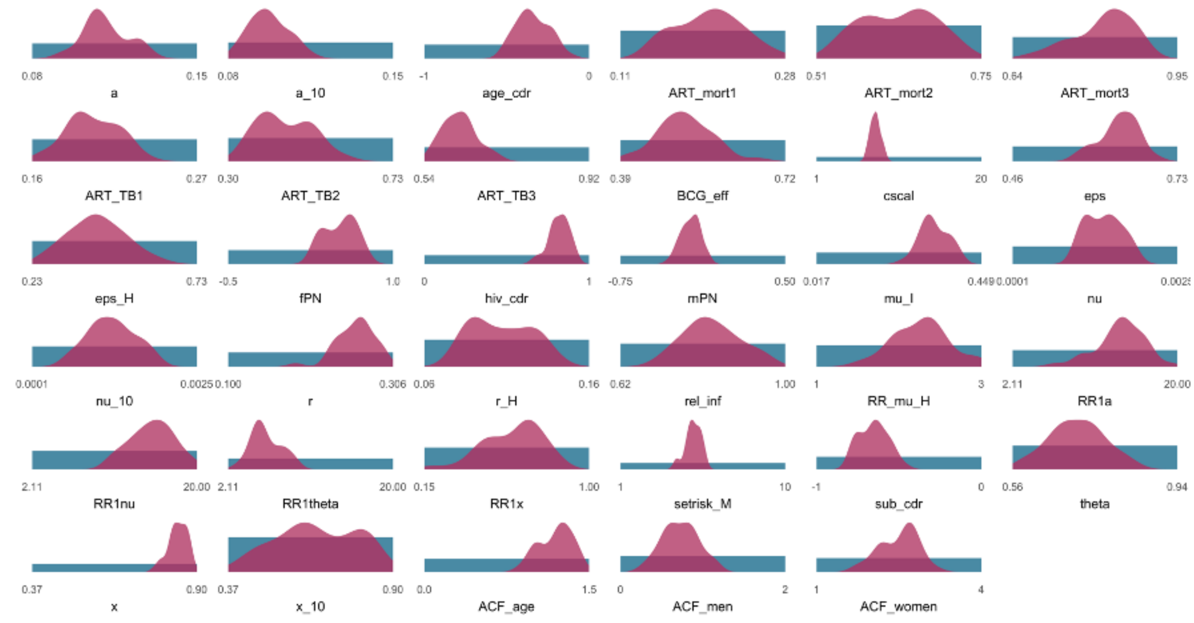

*Figure 16: The prior ranges and posterior distributions for each parameter for the 1000 selected parameter sets for Uganda*

*Table 26: The prior ranges and posterior central estimates and 95% uncertainty intervals from the 1000 calibrated parameter sets for Uganda*

| Parameters | Symbols | Description | Priors | Posteriors |
|---|---|---|---|---|
| a | $\alpha_{a\geq15}$ | Progression to disease, adults | 0.08-0.15 | 0.11 [UI 0.09-0.13] |
| a_10 | $\alpha_{a=0-14}$ | Progression to disease, children | 0.08-0.15 | 0.10 [UI 0.08-0.11] |
| age_cdr | $p_{a=0-14}$ | Treatment coverage adjustment, children | -1.00-0.00 | -0.35 [UI -0.50--0.18] |
| ART_mort1 | $ART_{\mu_{\mathbf{sTB}_{t<6}}}$ | ART mortality adjustment, 0-6 months | 0.11-0.28 | 0.20 [UI 0.15-0.26] |
| ART_mort2 | $ART_{\mu_{\mathbf{sTB}_{t=6-12}}}$ | ART mortality adjustment, 6-12 months | 0.51-0.75 | 0.64 [UI 0.54-0.71] |
| ART_mort3 | $ART_{\mu_{\mathbf{sTB}_{t\geq12}}}$ | ART mortality adjustment, 12+ months | 0.64-0.95 | 0.82 [UI 0.70-0.89] |
| ART_TB1 | $ART_{TB_{t<6}}$ | ART risk reduction, 0-6 months | 0.16-0.27 | 0.20 [UI 0.17-0.23] |
| ART_TB2 | $ART_{TB_{t=6-12}}$ | ART risk reduction, 6-12 months | 0.30-0.73 | 0.43 [UI 0.34-0.58] |

| | | | | |
|---|---|---|---|---|
| ART_TB3 | $ART_{TB_{t\geq12}}$ | ART risk reduction, 12+ months | 0.54-0.92 | 0.61 [UI 0.56-0.70] |
| BCG_eff | $BCG_{eff_{a\geq15}}$ | BCF efficacy | 0.39-0.72 | 0.52 [UI 0.41-0.61] |
| cscal | $cscal$ | Contact scaling parameter | 1.00-20.00 | 7.78 [UI 6.85-8.78] |
| eps | $\epsilon_{h=HIV-}$ | Regression through disease, HIV- | 0.46-0.73 | 0.64 [UI 0.57-0.68] |
| eps_H | $\epsilon_{h=HIV+}$ | Regression through disease, HIV+ | 0.23-0.73 | 0.42 [UI 0.27-0.58] |
| fPN | $p_{g=F}$ | Treatment coverage adjustment, women | -0.50-1.00 | 0.55 [UI 0.30-0.74] |
| hiv_cdr | $p_{k=ART}$ | Treatment coverage adjustment, ART | 0.00-1.00 | 0.83 [UI 0.69-0.91] |
| mPN | $p_{g=M}$ | Treatment coverage adjustment, men | -0.75-0.50 | -0.22 [UI -0.34--0.09] |
| mu_I | $\mu_{\mathbf{sTB}}$ | TB mortality from sympomatic disease | 0.02-0.45 | 0.32 [UI 0.26-0.39] |
| nu | $\nu_{a\geq15}$ | Progression from infection, adults | 0.00-0.00 | 0.00 [UI 0.00-0.00] |
| nu_10 | $\nu_{a=0-14}$ | Progression from infection, children | 0.00-0.00 | 0.00 [UI 0.00-0.00] |
| r | $r_{h=HIV-}$ | Self cure, HIV- | 0.10-0.31 | 0.26 [UI 0.22-0.30] |
| r_H | $r_{h=HIV+}$ | Self cure, HIV+ | 0.06-0.16 | 0.11 [UI 0.08-0.14] |
| rel_inf | $q$ | Relative infectiousness, asymptomatic disease | 0.62-1.00 | 0.82 [UI 0.73-0.95] |
| RR_mu_H | $RR_{\mu_H}$ | Relative risk of mortality, HIV+ | 1.00-3.00 | 2.28 [UI 1.63-2.94] |
| RR1a | $RR_{1_\alpha}$ | Relative risk of progression to disease, HIV+ | 2.11-20.00 | 13.93 [UI 8.85-17.47] |
| RR1nu | $RR_{1_\nu}$ | Relative risk of progression from infection, HIV+ | 2.11-20.00 | 15.29 [UI 10.68-19.30] |
| RR1theta | $RR_{1_\theta}$ | Relative risk of progression through disease, HIV+ | 2.11-20.00 | 5.80 [UI 3.57-9.36] |

| | | | | |
|---|---|---|---|---|
| RR1x | $RR_{1_x}$ | Relative risk of reinfection, HIV+ | 0.15-1.00 | 0.65 [UI 0.30-0.82] |
| setrisk_M | $RR_{sex_{g=M,a\geq15}}$ | Excess risk for men | 1.00-10.00 | 4.99 [UI 4.04-5.59] |
| sub_cdr | $p_{d=\mathbf{aTB}}$ | Treatment coverage adjustment, asymptomatic disease | -1.00-0.00 | -0.65 [UI -0.78--0.51] |
| theta | $\theta$ | Progression through disease | 0.56-0.94 | 0.71 [UI 0.59-0.82] |
| x | $x_{a\geq15}$ | Reinfection, adults | 0.37-0.90 | 0.84 [UI 0.78-0.88] |
| x_10 | $x_{a=0-14}$ | Reinfection, chilldren | 0.37-0.90 | 0.64 [UI 0.44-0.83] |
| ACF_age | $f_{a=0-14,g}$ | Impact of CAST, children | 0.00-1.50 | 1.23 [UI 0.93-1.41] |
| ACF_men | $f_{a\geq15,g=M}$ | Impact of CAST, men | 0.00-2.00 | 0.73 [UI 0.36-1.12] |
| ACF_women | $f_{a\geq15,g=F}$ | Impact of CAST, women | 1.00-4.00 | 2.58 [UI 1.79-3.02] |

All selected parameter sets have model outputs that fit within the target ranges (Figure 17). Overall prevalence estimates lie on the higher end of the calibration targets, whilst prevalence estimates for men and women are more central, along with incidence and mortality estimates. P:N ratios start to reduce from 2015 onwards to 2022 after which they become more stationary, whilst the M:F prevalence ratio increases slightly over time, until 2023 when it reduces slightly.

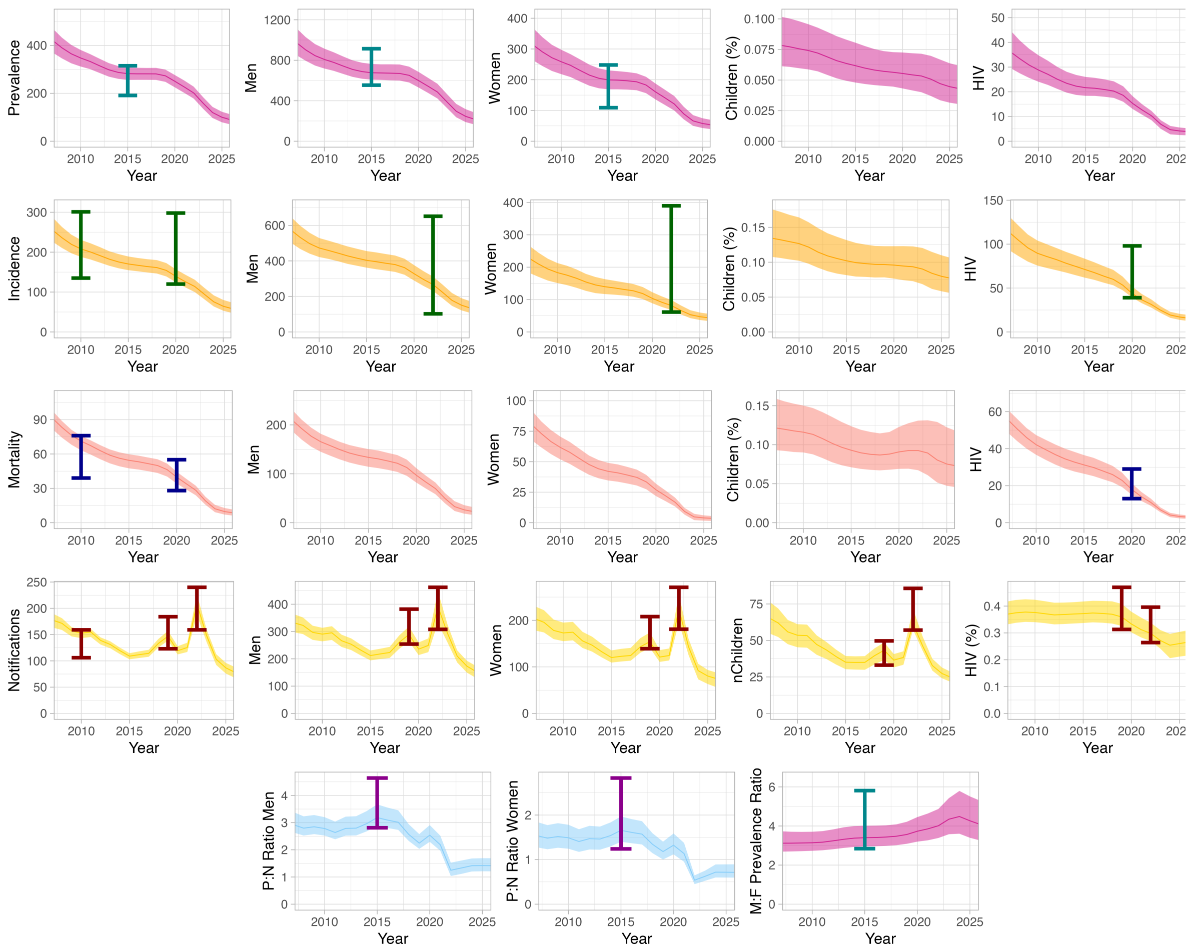

*Figure 17: Calibration plots for the Ugandan calibration targets using the 1000 selected parameters. The targets are the coloured error bars, the lines are the median estimates and the shaded regions are the 95% uncertainty intervals*

### 6 Implementation of interventions

The purpose of this model is to find the potential impact of implementing interventions specifically focussed on men. We have approached this with three different methods.

#### 6.1 Adjusting access to care

We know that men often do not access care at the same rate as women, and have included this in our assumptions around access to care in Section 3.6.1.1. We have assumed that it is possible to increase men's access to care to the same level achieved in women. Within the model, this is represented as increasing $p_{g=M}$ to equal $p_{g=F}$. Acknowledging that this change would not be possible overnight, we introduced a time dependent parameter to drive the change, $\delta_p$, that varies from 0 before 2025 to 1 after 2030. Over 5 years, from 2025 to 2030, the difference between the two parameters is decreased with an approximately S-shaped curve, as seen in Figure 18. The curve is derived from the equation $3x^2 - 2x^3$ with the values for each year taken from the evaluation of the curve at $\{0.2, 0.4, 0.6, 0.8\}$. Mathematically, this changes the formula for access to care to be

$$p_{a,d,h,k} = \left( \begin{cases} 1 + p_a & \text{if } a < 15 \\ 1 + p_k & \text{if on ART} \\ (1 + p_d) \times (1 + p_{g=F}) & \text{for women} \\ (1 + p_d) \times (1 + p_{g=M}) & \text{for men if } t < 2025 \\ (1 + p_d) \times \left(1 + \left(p_{g=M} + (p_{g=F} - p_{g=M}) \times \delta_p\right)\right) & \text{for men if } t \geq 2025 \end{cases} \right)$$

Beyond 2030, the formula for men and women (not on ART) is identical ($(1 + p_d) \times (1 + p_{g=F})$) as when $\delta_p = 1$, the expression $p_{g=M} + (p_{g=F} - p_{g=M}) \times \delta_p$ simplifies to $p_{g=F}$.

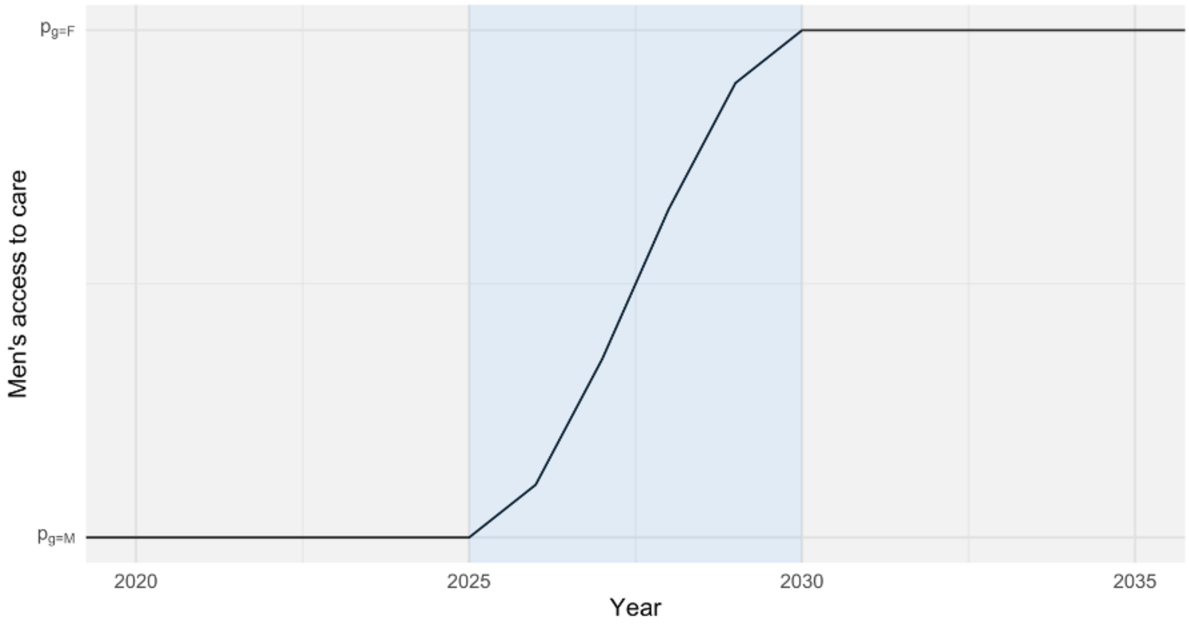

*Figure 18: The improvement of men's access to care over time, compared to women's access to care*

#### 6.2 Reducing excess risk

Men also have an excess risk of infection, ascribed to social and structural determinants, increased contact with men compared to women and children, and underlying biological factors. It will not be possible to eliminate the excess risk in men, however, it may be possible to reduce it through interventions focussed on the social and structural determinants that particularly differentially impact men. These could include smoking and drinking cessation interventions, social protection measures, or nutritional support. We are not stating which intervention, or combination of interventions, is being implemented, but instead looking at the broader statement of what is the impact if these interventions can reduce the risk by a given percentage. As with improving the access to care in Section 6.1, reducing the excess risk is achieved by introducing another time-dependent variable, $\delta_{RR_{sex}}$, which also uses an approximate S-shaped curve. The same equation for the curve is used as in Section 6.1, adapted to decrease from 1 to 0 and incorporate fractional decreases rather, $1-(3x^2-2x^3)\times w$, where $w$ is the proportion of risk reduction, with the values for each year taken from the evaluation of the curve at $\{0.2,0.4,0.6,0.8\}$. With this implementation, $RR_{sex_{g=M,a\geq 15}}$ becomes a time-dependent parameter calculated as:

$$RR_{sex_{g=M,a\geq 15,t}} = 1 + \left(RR_{sex_{g=M,a\geq 15}} - 1\right) \times \delta_{RR_{sex}}$$

Our main analyses have been conducted assuming a 50% reduction in risk is achievable, but sensitivity analyses have been conducted with 25% and 75% reductions.

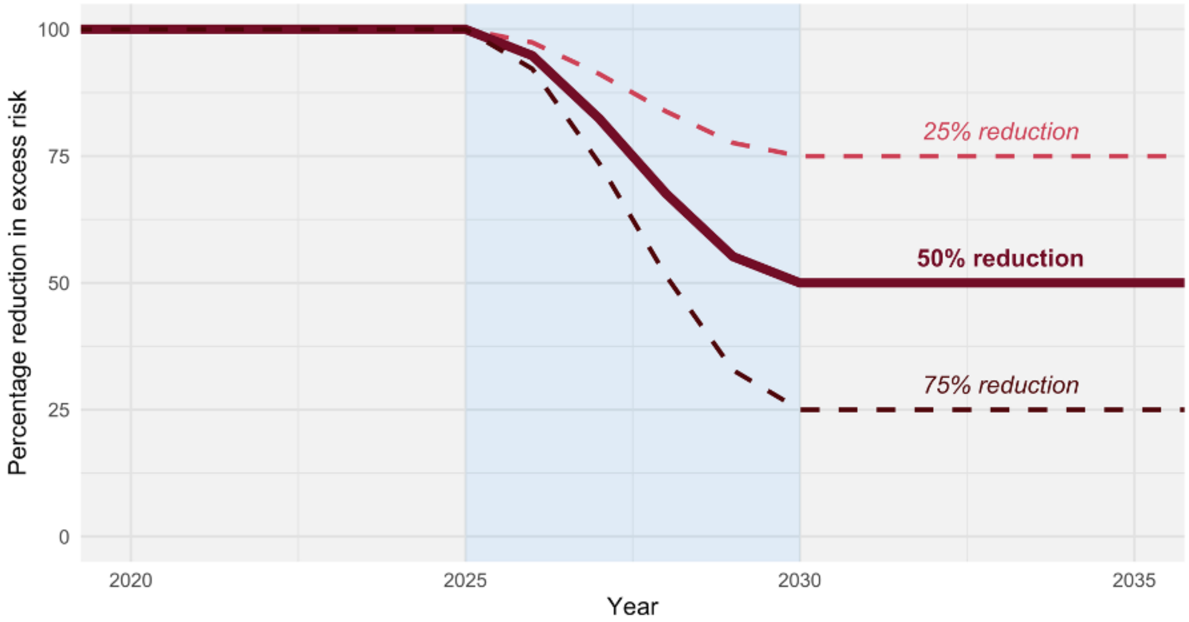

*Figure 19: The reduction in men's excess risk over time*

### 7 Supplementary results

#### 7.1 Country profiles

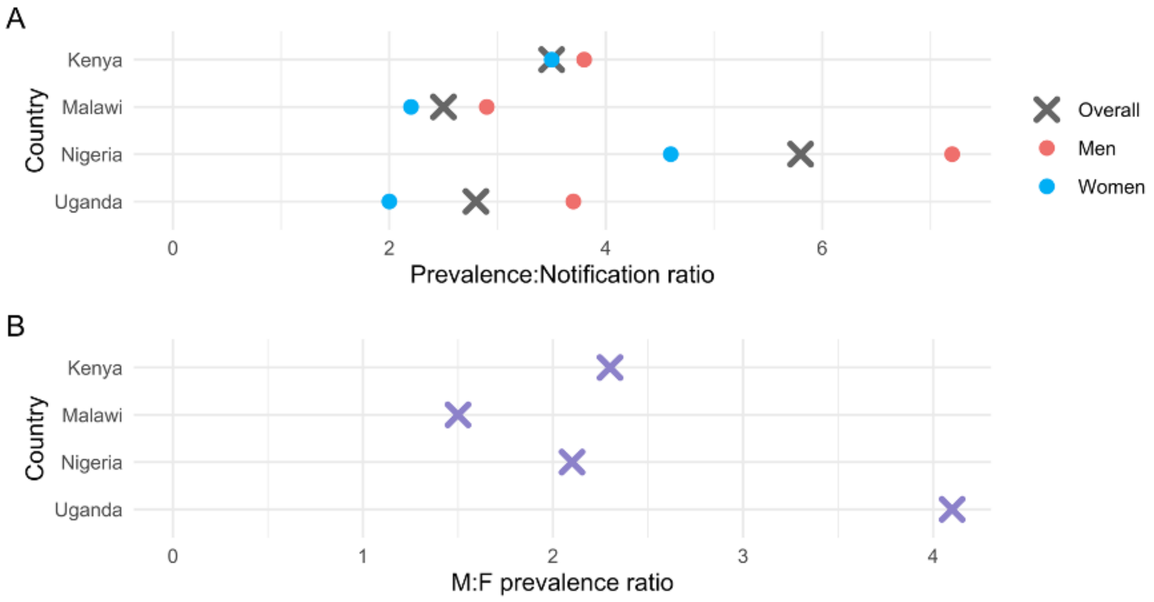

*Figure 20: Measures of sex-disparity in TB burden and care across the four countries through A) the ratio between TB prevalence and notifications overall and split by sex and B) the ratio between male and female bacteriologically positive TB*

#### 7.2 Impact of interventions on mortality and prevalence compared to incidence

Each strategy generally has a slightly larger impact on population-level mortality than it does on either prevalence or incidence, which are largely the same. However, the differences are not statistically significant as can be seen in Figure 21 and.

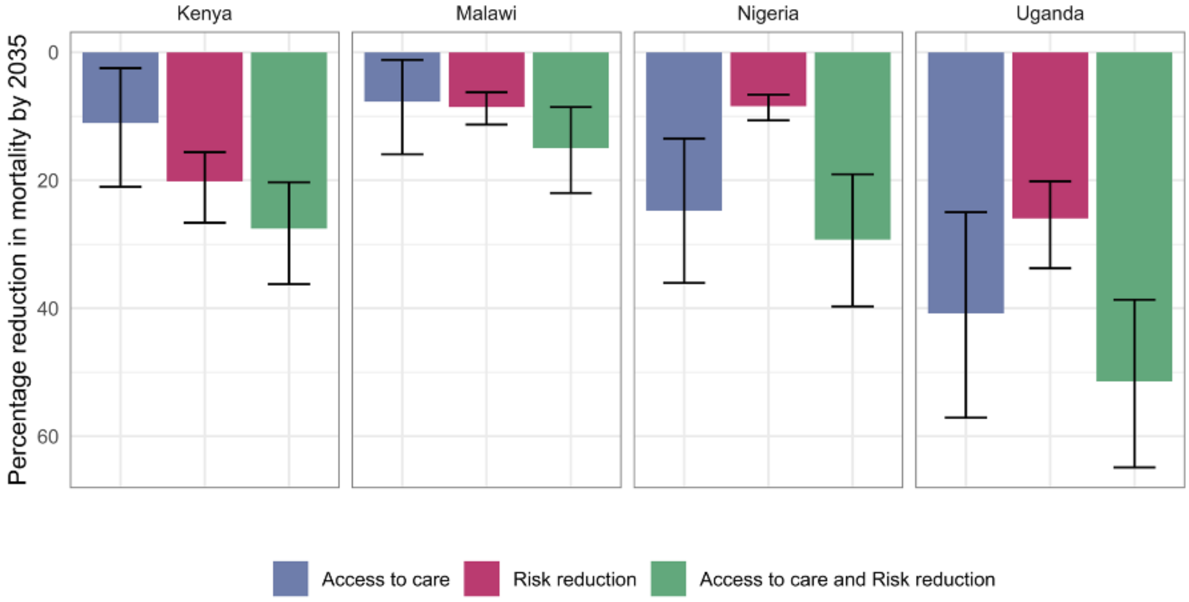

*Figure 21: The impact of each strategy on mortality at the population level*

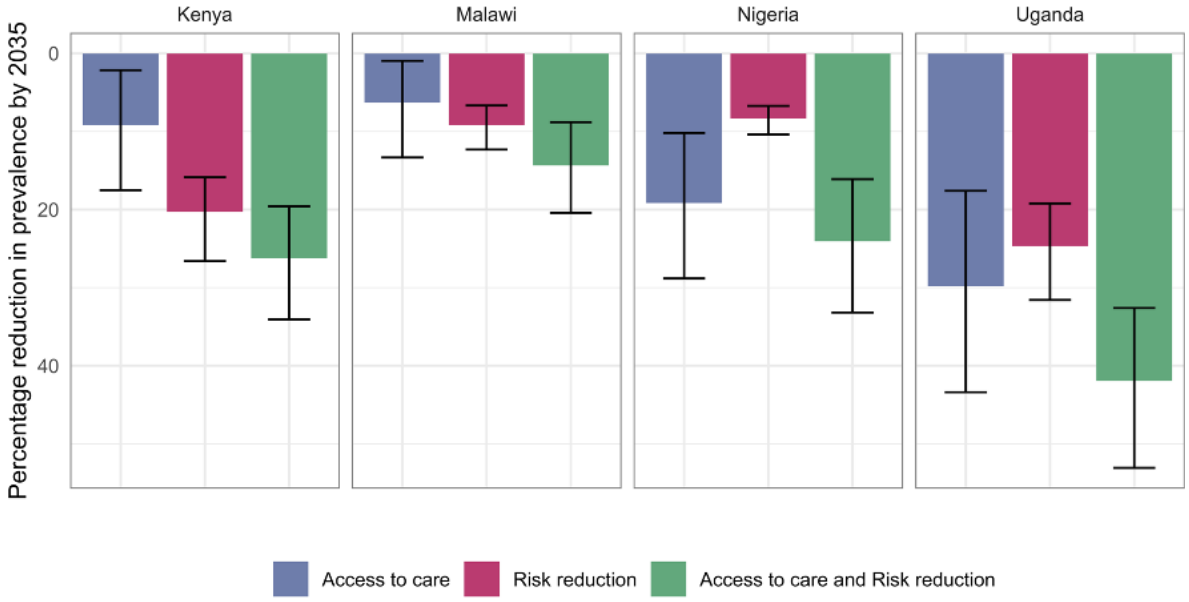

*Figure 22: The impact of each strategy on prevalence at the population level*

#### 7.3 Impact of interventions for varying levels of risk reduction

##### 7.3.1 Incidence

Increasing the levels of risk reduction increases the benefits achieved by strategies reducing risk, but brings less of an improvement to a combined strategy than it does to the single strategy as can be seen in Figure 23

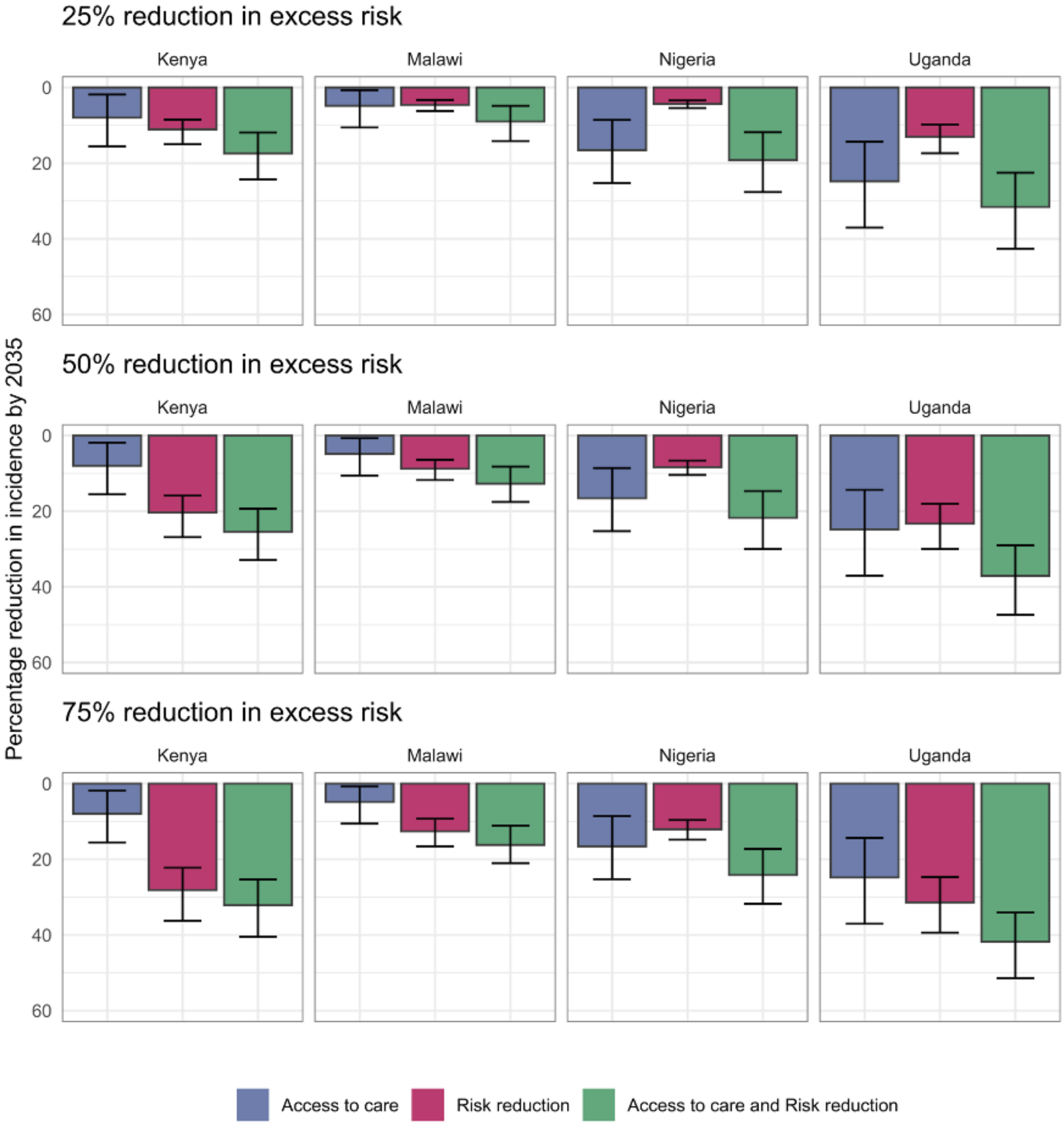

*Figure 23: The reduction in population-level incidence across the three strategies and across three different risk reduction levels*

##### 7.3.2 Notifications

The higher the level of risk reduction, the lower the peak of excess notifications in the combined strategy, and the closer the risk reduction and combined strategies are in change in notifications by 2035, as can be seen in Figure 24.

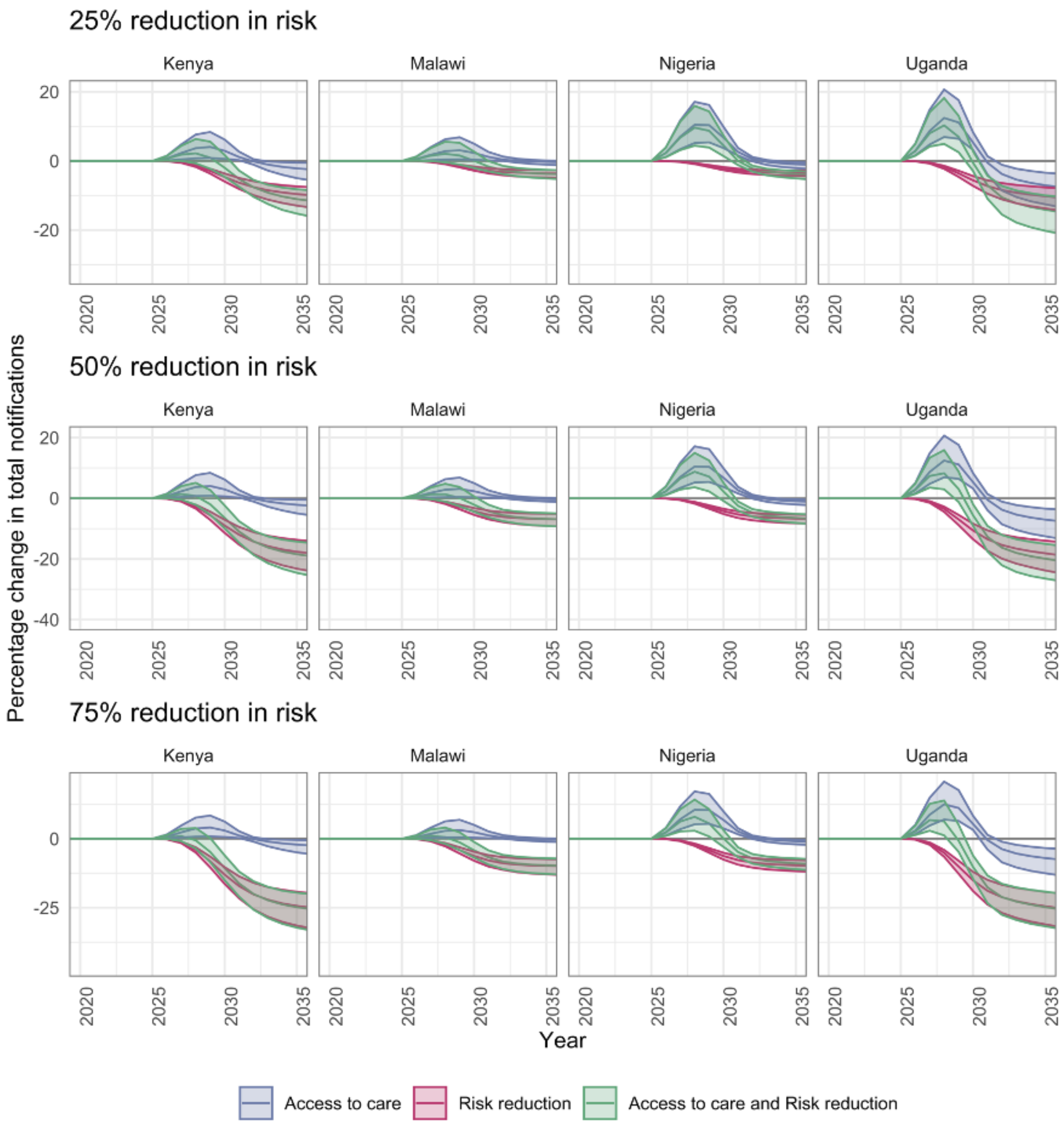

*Figure 24: The change in notifications as risk reduction increases*

#### 7.4 Impact out to 2050

##### 7.4.1 Incidence

Impact of all strategies increases by 2050 as compared to 2035, with the impact of reduction in risk increasing more than the impact of access to care as can be seen in Figure 25, likely because reduction in risk takes a longer time to take effect.

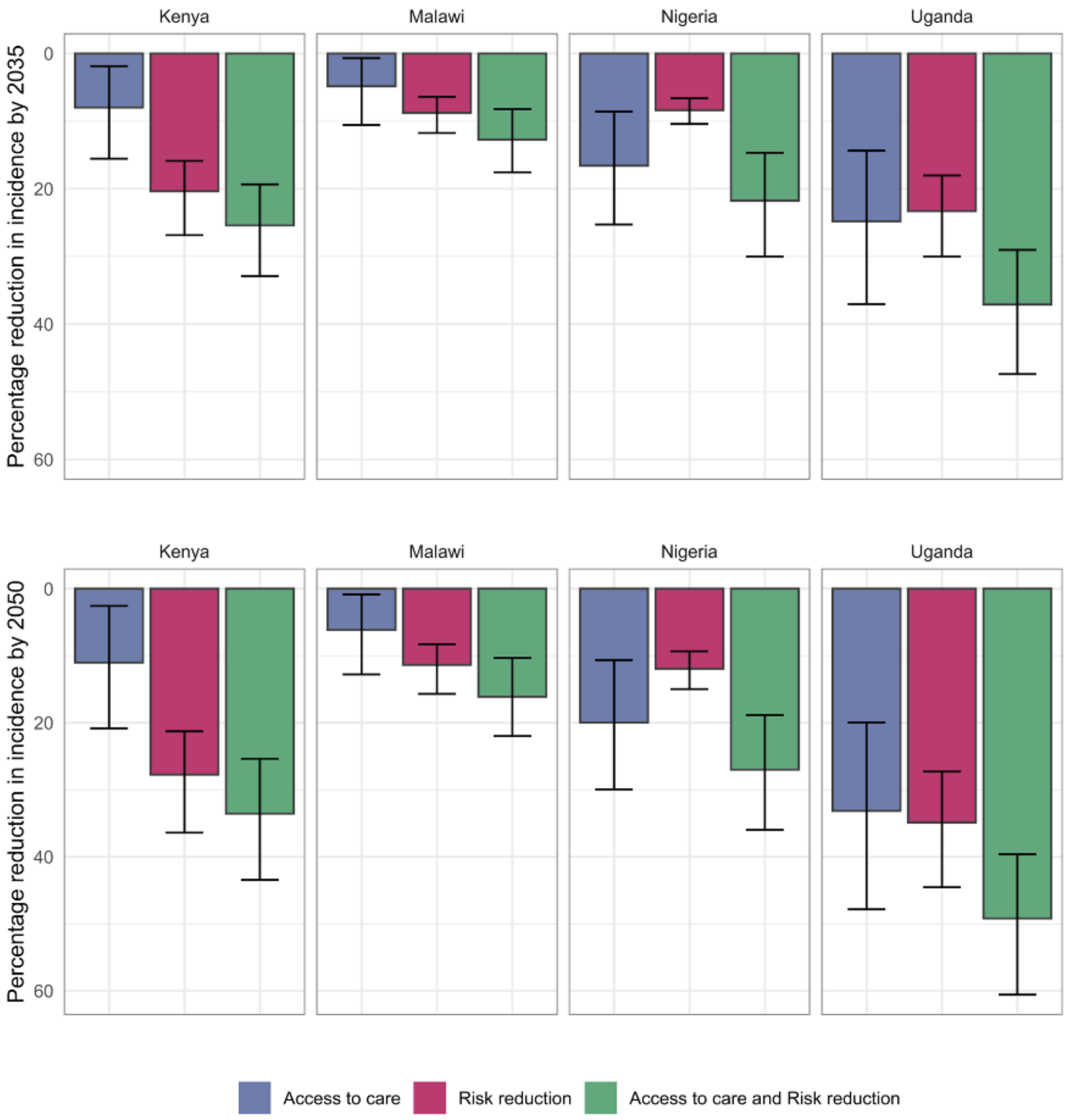

*Figure 25: The reduction in population-level incidence across the three strategies and across three different risk reduction levels*

##### 7.4.2 Notifications

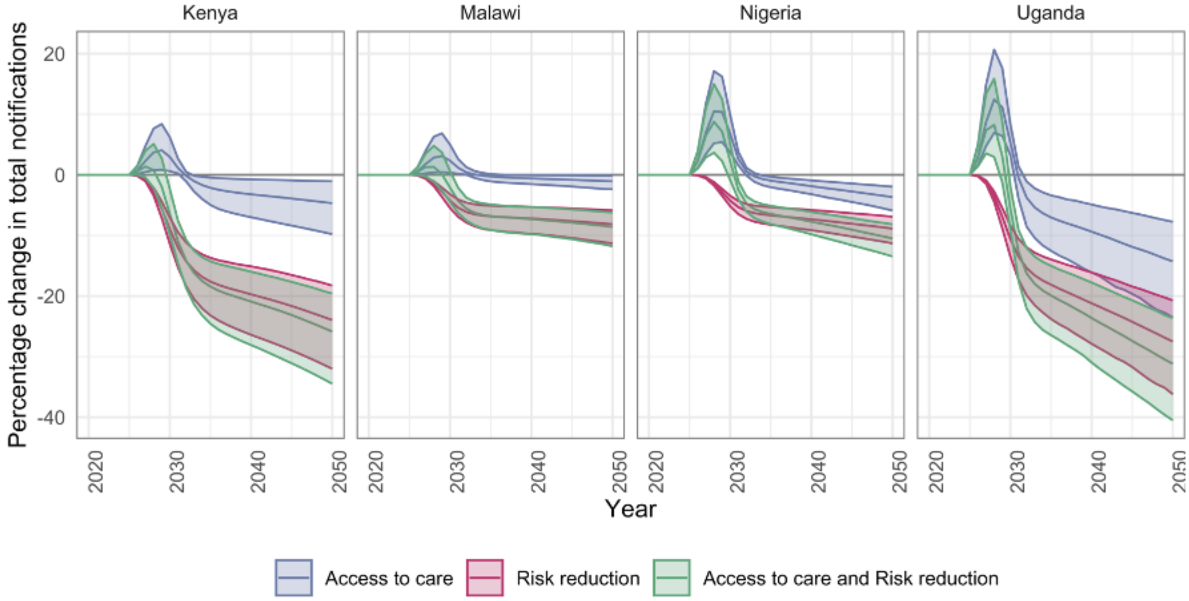

*Figure 26: The change in notifications for each scenario from 2020 to 2050 after the introduction of the three strategies between 2025 and 2030*

### 8 References

1. Horton KC, White RG, Hoa NB, Nguyen HV, Bakker R, Sumner T, et al. Population benefits of addressing programmatic and social determinants of gender disparities in tuberculosis in viet nam: A modelling study. PLOS Global Public Health. 2022;2: e0000784. doi:10.1371/journal.pgph.0000784

2. Richards AS, Sossen B, Emery JC, Horton KC, Heinsohn T, Frascella B, et al. Quantifying progression and regression across the spectrum of pulmonary tuberculosis: A data synthesis study. The Lancet Global Health. 2023;11: e684–e692. doi:10.1016/S2214-109X(23)00082-7

3. Horton KC, Richards AS, Emery JC, Esmail H, Houben RMGJ. Reevaluating progression and pathways following mycobacterium tuberculosis infection within the spectrum of tuberculosis. Proceedings of the National Academy of Sciences. 2023;120: e2221186120. doi:10.1073/pnas.2221186120

4. World Health Organization. Global tuberculosis report 2024. World Health Organization; 2024. Available: https://www.who.int/teams/global-tuberculosis-programme/tb-reports/global-tuberculosis-report-2024

5. Houben RMGJ, Lalli M, Sumner T, Hamilton M, Pedrazzoli D, Bonsu F, et al. TIME impact  a new user-friendly tuberculosis (TB) model to inform TB policy decisions. BMC Medicine. 2016;14: 56. doi:10.1186/s12916-016-0608-4

6. Stover J, McKinnon R, Winfrey B. Spectrum: A model platform for linking maternal and child survival interventions with AIDS, family planning and demographic projections. International Journal of Epidemiology. 2010;39: i7–i10. doi:10.1093/ije/dyq016

7. United Nations. World population prospects - population division - united nations. https://population.un.org/wpp/downloads; 2024.

8. Emery JC, Dodd PJ, Banu S, Frascella B, Garden FL, Horton KC, et al. Estimating the contribution of subclinical tuberculosis disease to transmission: An individual patient data analysis from prevalence surveys. Kana BD, editor. eLife. 2023;12: e82469. doi:10.7554/eLife.82469

9. Dye C, Bassili A, Bierrenbach A, Broekmans J, Chadha V, Glaziou P, et al. Measuring tuberculosis burden, trends, and the impact of control programmes. The Lancet Infectious Diseases. 2008;8: 233–243. doi:10.1016/S1473-3099(07)70291-8

10. Abu-Raddad LJ, Sabatelli L, Achterberg JT, Sugimoto JD, Longini IM, Dye C, et al. Epidemiological benefits of more-effective tuberculosis vaccines, drugs, and diagnostics. Proceedings of the National Academy of Sciences. 2009;106: 13980–13985. doi:10.1073/pnas.0901720106

11. Knight GM, Griffiths UK, Sumner T, Laurence YV, Gheorghe A, Vassall A, et al. Impact and cost-effectiveness of new tuberculosis vaccines in low- and middle-income

countries. Proceedings of the National Academy of Sciences. 2014;111: 15520–15525. doi:10.1073/pnas.1404386111

12. Baguelin M, Flasche S, Camacho A, Demiris N, Miller E, Edmunds WJ. Assessing optimal target populations for influenza vaccination programmes: An evidence synthesis and modelling study. PLOS Medicine. 2013;10: e1001527. doi:10.1371/journal.pmed.1001527

13. Del Fava E, Adema I, Kiti MC, Poletti Pp, Merler S, Nokes DJ, et al. Individual's daily behaviour and intergenerational mixing in different social contexts of kenya | scientific reports. Scientific Reports. 2021;11: 21589. doi:10.1038/s41598-021-00799-1

14. Thindwa D, Jambo KC, Ojal J, MacPherson P, Dennis Phiri M, Pinsent A, et al. Social mixing patterns relevant to infectious diseases spread by close contact in urban Blantyre, Malawi. Epidemics. 2022;40: 100590. doi:10.1016/j.epidem.2022.100590

15. Kleynhans J, Tempia S, McMorrow ML, Gottberg A von, Martinson NA, Kahn K, et al. A cross-sectional study measuring contact patterns using diaries in an urban and a rural community in South Africa, 2018. BMC public health. 2021;21: 1055. doi:10.1186/s12889-021-11136-6

16. Miller PB, Zalwango S, Galiwango R, Kakaire R, Sekandi J, Steinbaum L, et al. Association between tuberculosis in men and social network structure in Kampala, Uganda. BMC infectious diseases. 2021;21: 1023. doi:10.1186/s12879-021-06475-z

17. Potter GE, Wong J, Sugimoto J, Diallo A, Victor JC, Neuzil K, et al. Networks of face-to-face social contacts in Niakhar, Senegal. PloS One. 2019;14: e0220443. doi:10.1371/journal.pone.0220443

18. Marquez C, Chen Y, Atukunda M, Chamie G, Balzer LB, Kironde J, et al. The association between social network characteristics and tuberculosis infection among adults in 9 rural ugandan communities. Clinical Infectious Diseases. 2023;76: e902–e909. doi:10.1093/cid/ciac669

19. Horton KC, MacPherson P, Houben RMGJ, White RG, Corbett EL. Sex differences in tuberculosis burden and notifications in low- and middle-income countries: A systematic review and meta-analysis. PLOS Medicine. 2016;13: e1002119. doi:10.1371/journal.pmed.1002119

20. Dowdy DW, Chaisson RE. The persistence of tuberculosis in the age of DOTS: Reassessing the effect of case detection. Bulletin of the World Health Organization. 2009;87: 296–304. doi:10.2471/BLT.08.054510

21. Menzies NA, Cohen T, Lin H-H, Murray M, Salomon JA. Population health impact and cost-effectiveness of tuberculosis diagnosis with xpert MTB/RIF: A dynamic simulation and economic evaluation. PLOS Medicine. 2012;9: e1001347. doi:10.1371/journal.pmed.1001347

22. Dye C, Garnett GP, Sleeman K, Williams BG. Prospects for worldwide tuberculosis control under the WHO DOTS strategy. The Lancet. 1998;352: 1886–1891. doi:10.1016/S0140-6736(98)03199-7

23. Eamranond P, Jaramillo E. Tuberculosis in children: Reassessing the need for improved diagnosis in global control strategies. The International Journal of Tuberculosis and Lung Disease. 2001;5: 594–603.

24. World Health Orgaanization. Bacillus calmette-guérin (BCG) vaccination coverage. https://immunizationdata.who.int/pages/coverage/BCG.html?CODE=Global+KEN+NGA+UGA+MWI&YEAR=; 2024.

25. Dutta T. Sex-disaggregated immunization coverage data: Input from key stakeholders. PATH; 2010 Oct.

26. Dodd PJ, Gardiner E, Coghlan R, Seddon JA. Burden of childhood tuberculosis in 22 high-burden countries: A mathematical modelling study. The Lancet Global Health. 2014;2: e453–e459. doi:10.1016/S2214-109X(14)70245-1

27. Sonnenberg P, Glynn JR, Fielding K, Murray J, Godfrey-Faussett P, Shearer S. How soon after infection with HIV does the risk of tuberculosis start to increase? A retrospective cohort study in south african gold miners. The Journal of Infectious Diseases. 2005;191: 150–158. Available: https://www.jstor.org/stable/30077675

28. Lawn SD, Kranzer K, Wood R. Antiretroviral therapy for control of the HIV-associated tuberculosis epidemic in resource-limited settings. Clinics in Chest Medicine. 2009;30: 685–699. doi:10.1016/j.ccm.2009.08.010

29. Ferebee SH. Controlled chemoprophylaxis trials in tuberculosis. A general review. Bibl Tuberc. 1970;26: 28–106. Available: http://www.ncbi.nlm.nih.gov/pubmed/4903501

30. Gilks CF, Godfrey-Faussett P, Batchelor BIF, Ojoo JC, Ojoo SJ, Brindle RJ, et al. Recent transmission of tuberculosis in a cohort of HIV-1-infected female sex workers in nairobi, kenya. AIDS. 1997;11: 911. Available: https://journals.lww.com/aidsonline/abstract/1997/07000/recent_transmission_of_tuberculosis_in_a_cohort_of.11.aspx

31. Sutherland I, Svandova E, Radhakrishna S. The development of clinical tuberculosis following infection with tubercle bacilli. Tubercle. 1982;63: 255–68. Available: **Error! Hyperlink reference not valid.**

32. Cohen T, Lipsitch M, Walensky RP, Murray M. Beneficial and perverse effects of isoniazid preventive therapy for latent tuberculosis infection in HIVtuberculosis coinfected populations. Proceedings of the National Academy of Sciences. 2006;103: 7042–7047. doi:10.1073/pnas.0600349103

33. Dye C, Williams BG. Criteria for the control of drug-resistant tuberculosis. Proceedings of the National Academy of Sciences of the United States of America. 2000;97: 8180–8185. doi:10.1073/pnas.140102797

34. Horton KC, Sumner T, Houben RMGJ, Corbett EL, White RG. A Bayesian approach to understanding gender differences in tuberculosis disease burden. American Journal of Epidemiology. 2018;187: 2431--2438. doi:10.1093/aje/kwy131

35. World Health Organization Global Tuberculosis programme. WHO TB burden estimates. https://www.who.int/teams/global-tuberculosis-programme/data; 2024.

36. Kenya Ministry of Health, National Tuberculosis, Leprosy and Lung Disease Programme. Kenya national strategic plan for 2023/24 to 2027/28. 2023. Available: https://nltp.co.ke/national-strategic-plan-2019-2023-3/

37. Turyahabwe S, Bamuloba M, Mugenyi L, Amanya G, Byaruhanga R, Fry Imoko J, et al. Community tuberculosis screening, testing and care, uganda. Bulletin of the World Health Organization. 2024;102: 400–409. doi:10.2471/BLT.23.290641

38. World Health Organization Global Tuberculosis programme. Treatment success rate: HIV-positive TB cases. https://www.who.int/data/gho/data/indicators/indicator-details/GHO/treatment-success-rate-hiv-positive-tb-cases; 2023.

39. Ragonnet R, Flegg JA, Brilleman SL, Tiemersma EW, Melsew YA, McBryde ES, et al. Revisiting the natural history of pulmonary tuberculosis: A bayesian estimation of natural recovery and mortality rates. Clinical Infectious Diseases. 2021;73: e88–e96. doi:10.1093/cid/ciaa602

40. R Core Team. R: A language and environment for statistical computing. Vienna, Austria: R Foundation for Statistical Computing; 2024. Available: https://www.R-project.org/

41. Press WH, Teukolsky SA, Vetterling WT, Flannery BP. Numerical recipes 3rd edition: The art of scientific computing. 3rd ed. Hardcover; Cambridge University Press; 2007. Available: http://www.amazon.com/exec/obidos/redirect?tag=citeulike07-20\&path=ASIN/0521880688

42. National Tuberculosis L, Lung Disease Program R of K Ministry of Health. Kenya tuberculosis prevalence survey 2016. Nairobi, Kenya; 2018. Available: https://www.nltp.co.ke/survey-reports-2/

43. Ministry of Health NTCP. Technical report: Malawi tuberculosis prevalence survey (2013–2014). 2016.

44. Department of Public Health FR of N. Report of the first national TB prevalence survey 2012, nigeria. 2014.

45. Public Health MUS of. Report on the population-based survey of prevalence of tuberculosis disease in uganda 2014–15. Kampala, Uganda; 2016. Available: https://library.health.go.ug/index.php/communicable-disease/tuberculosis/uganda-national-tuberculosis-prevalence-survey-2014-2015-survey

46. Law I, Floyd K, Group the ATPS. National tuberculosis prevalence surveys in africa, 20082016: An overview of results and lessons learned. Tropical Medicine & International Health. 2020;25: 1308–1327. doi:10.1111/tmi.13485

47. WHO Global Tuberculosis programme. WHO TB incidence estimates disaggregated by age group, sex, and risk factor. https://www.who.int/teams/global-tuberculosis-programme/data; 2024.

48. UN World Population Prospects. Population by select age groups - male. https://population.un.org/wpp/Download/Standard/Population/; 2024.

49. UN World Population Prospects. Population by select age groups - female. https://population.un.org/wpp/Download/Standard/Population/; 2024.

50. WHO Global Tuberculosis programme. Case notifications. https://www.who.int/teams/global-tuberculosis-programme/data; 2024.

51. Tollefson D, Ngari F, Mwakala M, Gethi D, Kipruto H, Cain K, et al. Under-reporting of sputum smear-positive tuberculosis cases in kenya. The International Journal of Tuberculosis and Lung Disease. 2016;20: 1334–1341. doi:10.5588/ijtld.16.0156

52. Gidado M, Mitchell EMH, Adejumo AO, Levy J, Emperor O, Lawson A, et al. Assessment of TB underreporting by level of reporting system in lagos, nigeria. Public Health Action. 2022;12: 115–120. doi:10.5588/pha.22.0008

53. WHO Global Tuberculosis programme. WHO TB profiles. https://worldhealthorg.shinyapps.io/tb_profiles/; 2024.

54. Iskauskas A, Vernon I, Goldstein M, Scarponi D, McCreesh N, McKinley TJ, et al. Emulation and history matching using the hmer package. Journal of Statistical Software. 2024;109: 1–48. doi:10.18637/jss.v109.i10

55. RStudio Team. RStudio: Integrated development environment for r. Boston, MA: RStudio, PBC.; 2020. Available: http://www.rstudio.com/

56. Ogoamaka C, Bethrand O, Lotanna U, Chidubem O, Sani U, Nkiru N, et al. The TB surge intervention: An optimized approach to TB case-finding in nigeria. Public Health Action. 2023;13: 136–141. doi:10.5588/pha.23.0039
